# Unmasking the Current Scenario of Indian Biomedical Devices Industry: Ventilators being the Heart of the Discussion

**DOI:** 10.1101/2021.02.26.21251720

**Authors:** Sheersha Pramanik, Sucheta Karmakar, Shreyas Mukherjee, Indraneel Dhavale, Rohan Shrestha

**Affiliations:** Dept. of Biotechnology, Bhupat and Jyoti Mehta School of Biosciences, Indian Institute of Technology Madras, Chennai – 600036, India; Dr. B.C. Roy College of Pharmacy and Allied Health Sciences, Affiliated to MAKAUT, Meghnad Saha Sarani, Bidhannagar, Durgapur, West Bengal-713206, India; Jamia Hamdard, Mehrauli-Badarpur Rd, Near Batra Hospital, Block D, Hamdard Nagar, New Delhi, Delhi-110062, India; Dept. of Pharmaceutical Sciences and Technology, Institute of Chemical Technology, Nathalal Parekh Marg, Near Khalsa College, Matunga, Mumbai, Maharashtra-400019, India; Amity Institute of Pharmacy, Amity University, Amity Road, Sector 125, Noida, Uttar Pradesh-201301, India

**Keywords:** Medical devices industry, Medical devices, SARS-CoV-2, Coronavirus, Ventilators, Ventilator-associated Pneumonia, Barotrauma, Microaspiration

## Abstract

**Background:** The Indian Biomedical Device Industry has been growing at an unprecedented rate, but several hindrances need to be acknowledged in offering access to quality, budget-friendly medical devices in India. This article explores the current loopholes of the Indian biomedical device industry along with the proposal of various innovative solutions, with emphasis on ventilators.

**Methods:** An online survey with the help of Google forms was conducted from 1^st^ November to 25^th^ December 2020, addressing the problems of the Indian medical device industry (MDI) along with probable solutions. The survey also provides a glimpse into the complications aroused from the frequent use of ventilators during COVID-19 outburst, along with possible measures.

**Results:** According to the survey, 51.6%, 46.5%, 55.7%, and 47.5% of respondents have agreed to ‘Stringent laws and implementation,’ ‘Safety testing and strict regulations,’ ‘Unfavorable duty structure,’ and ‘Reducing tax rate on domestic manufacturers,’ as a possible solution to advance the implementation of ISO 13485 and ISO 10651, implementation of ISO 14971, the probable cause of limitations of Indian MDI, possible measure to overcome limitations of Indian MDI, respectively. 46.7%, 47.5%, 46.2%, and 44.6% of respondents have agreed to ‘Surgical decompression,’ ‘Nebulized and broad-spectrum antibiotics,’ ‘PEEP,’ and ‘Adopting lung protective ventilation strategies’ as possible solutions to treat various ventilator complications, respectively.

**Conclusion:** The study briefs about the people’s perception of the Indian MDI as well as on the ventilator complications. The results complied with our hypothesis as the majority of the respondents have agreed with almost all the probable solutions in both the sections given by us as options.

## **1.** Introduction

In the wake of COVID 19, we have had many implications, some positive and some negative. As they say, necessity is the mother of invention, and only upon seeing the impact of the pandemic have we as a nation began to understand the importance of Pharmacy and Biomedical Sciences in minimizing the negative impacts and creating long-lasting positive changes for generations to come. The “Make in India” initiative started way back in 2014 by Prime Minister Narendra Modi, paved the way for the “Self Reliant Movement” (Aatmanirbhar Bharat Abhiyaan) in 2020 amidst the coronavirus pandemic [1]. We now see the importance and the need to be free, a type of second independence, to be independent as a nation in our production of biomedical supplies and devices. At present, around 75-80% is India’s overall import dependency, which is quite surprising when considering India to be among the top-20 markets for medical devices globally. The current scenario in terms of the market size in India is estimated at 11 billion dollars. This figure is expected to go up to 50 billion dollars by 2025. According to the statistics of the year 2018-19, imports have been valued at 6.2 billion dollars, whereas the exports have been a meager $2.1 billion. India’s expected export of medical devices will reach an estimated $10 billion by 2025 [2].

Upon inspection, it is seen that the biomedical sector in India is far from perfect. There are numerous limitations we need to face and work on. India has a long way too, but just as India is a rapidly developing nation, its biomedical sector is rapidly improving and evolving. As we look into the limitations, we find various factors responsible primarily in the industry’s macro-environment [3]. The sector of the biomedical device is relatively capital intensive, and in the present scenario, the cost of financing is in the range of 14-18% and thus, smaller companies face immense financial burden when setting up manufacturing and production facilities in the country of India [4].

This is only the tip of the iceberg as there are various factors pertaining to numerous shortcomings in the biomedical sector as we look into the industry, such as unfavorable duty structure, weak implementation, and lack of comprehensive laws primarily for the intellectual property of medical devices, absence of reputable quality certification authority. Due to the sharp contrast of the unfavorable duty structure on production and manufacturing concerning importing biomedical devices from foreign countries, it is often seen as favorable in monetary terms to simply import the required biomedical devices rather than focus and improve upon the indigenous facilities in India [5]. Quality Certification of products in India is not at par with foreign companies, as India lacks a standard Quality Certification authority such as the FDA or EU. The absence of a reputable certification authority that licenses the quality of the products reduces the country’s opportunities to export its product and expand its market. Indigenous manufacturers are still required to get an FDA/CE certification to be available in India and the rest of the world. Another problem with India’s certification process is that the approval process can take a considerable amount of time, negatively impacting the sector, prolonging the time for the product to reach the market [6].

A large part of the solutions is to analyze and critically evaluate the biomedical industry’s limitations and achieve sustainable goals. As we have looked into the shortcomings, we can channel our efforts towards bettering India’s biomedical device industry. One of the most important aspects of improvement is the development of better ecosystem support. The biomedical device sector should be holistic in its functioning and with various complementary industries that contribute to the functioning and supporting the niche of the industry. It is essential to establish an adequate ecosystem in India to provide a sense of safety and assurance for domestic companies, both big and small, while at the same time attracting foreign companies to invest and expand on our soil, thus further benefiting the sector and our economy [7]. Similarly, it is essential to emphasize the importance of indigenous development of biomedical devices and the industry. As COVID 19 brought the world to a standstill, nations were made to improve, adapt, and work towards a brighter future. This brought attention to the medical and pharmacy sector like never before as the importance of the industry in the country’s development, and the well-being of its citizens was realized. As the market has been dependent majorly upon imports, indigenous innovation and research have been limited. With the exclusion of a few players, most have looked at India as one of many export markets, with a primary focus on extensive sales and distribution. As trades with other nations dwindle and the remarkable effect of COVID 19, India will be forced to break free of its chains and become independent in its production and development of biomedical devices. With the newfound awareness and importance brought to this essential sector, the government, along with the respective authorities, have taken various initiatives in the right direction and implemented laws to better the industry. Medical devices are now classified as drugs and have levels of risk and stringent regulations, and have incentives and benefits depending on the type of branch of healthcare the biomedical device works on. Such initiatives are exactly what we need to bring change and improve the sector [8].

The SARS-CoV-2 pandemic and its unparalleled worldwide societal and economic obstreperous effect has pronounced the third zoonotic initiation of an extremely infective coronavirus into the human community. The SARS-CoV-2 virus causes inflammation in the airways and lungs, making it difficult for the patient to breathe due to low oxygen levels. The ventilator is used to force the air with increased oxygen levels into the lungs to encounter this problem. According to the ISO 19223:2019, ‘Ventilator’ is defined as a medical device or medical electrical equipment intended to provide artificial ventilation (cyclical movement of respirable gas into and out of the lungs)[9]. The ventilator uses positive pressure to provide oxygen to the lungs. A humidifier is also included in the ventilator for adding moisture and heat to the oxygen to match the patient’s body temperature [10]. The ventilator doesn’t cure COVID-19 but improves the patient’s survival chance and helps until the lungs perform breathing independently.

India used to import the ventilators or parts for the manufacture from foreign countries from the starting phase. So, there was a lack of a separate regulatory standard for the ventilators. As it was stopped during the pandemic, India was in dire need of ventilators for COVID-19 patients. To overcome this situation, India focused on ramping up the production of low-cost ventilators ignoring safety and performance testing of the devices. To satisfy the need for raw materials for manufacture, automobile, and IT companies have joined with ventilator manufacturing companies, e.g., the partnership between Mahindra & Mahindra and Skanray Technologies under the Make In India initiative. The engineering institutes have also been joined together to develop low-cost and simplified versions of a ventilator, e.g., NOCCA V310 ICU Ventilator by Nocca Robotics under IIT Kanpur incubation [11]. In this surge of Covid 19, the use of ventilators has increased to a great extent [11]. Overuse of ventilators welcomes several complications like Ventilator-Associated pneumonia and microaspiration [12], barotrauma [13], Acute respiratory distress syndrome (ARDS) [14].

This research survey aims to pinpoint the stumbling blocks prevailing in the Indian biomedical industry, identify the patient’s problem caused due to frequent use of mechanical ventilators, and correlate the proposed groundwork solutions for both of the above significant issues with the perspective of the respondents.

## 2. Materials and Methods

### 2.1. Study Design and Sample

Survey research is a way to congregate data about a sample drawn from a precisely defined population of persons or organizations. It offers researchers an opportunity to gather useful data, although substitute tools such as remarks, panels, focus communities, and interviews might furnish, at times, excellent information. This study utilized a cross-sectional research questionnaire survey of bio-researchers and medical device industrialists of India. The population (n=315) included the students from the department of pharmacy and biomedical engineering. The respondents holding various degrees participated in the aforementioned research survey. A convenient sampling design was prepared for this study because of the time constraints imposed on the same.

The present survey comprises three sections: (1) demographics and personal information, (2) knowledge assessment on the Indian biomedical industry and ventilators, and (3) perceptions evaluation (5-point Likert scale questions).

A 5-point Likert-type scale questionnaire (SDA – Strongly Disagree, DA – Disagree, N - Neutral, A – Agree, SA – Strongly Agree) was used to collect data for different research problems. The left-over questions were based on selecting the most suitable option/options by using a tick mark (√) and closed questions (Yes/No/Maybe).

For better understanding, the readers are referred to check the survey instrument (see Appendix 1).

### 2.2. Data collection

The data collection tool was first pilot-tested with selected pharma/biomedical students (5-10 people) for better understanding. The final google form questionnaire was then disseminated via different social handles and emails to the targeted communities. The survey started on 1^st^ Nov 2020. The targeted communities included biomedical engineering, pharmacy students/researchers/industrialists who had a basic knowledge of the research topic. It was suggested that every community of people pass the questionnaire survey to their respected colleagues for a better sample score. A URL of the google form was provided to our targeted population with a specific deadline to complete the survey electronically. In addition to the primary emails, the survey conductors made follow-up emails and messages to complete their filling of the survey. The respondents were provided sufficient time to fill the survey, which closed on 25^th^ December 2020. All the data provided by the respondents were confidential and were not exposed publically. The people who pre-or pilot-tested the questionnaire survey were ruled out from study contribution.

### 2.3. Data analysis

IBM SPSS Statistics Version 26.0 for Windows (IBM Corp., Armonk, NY, U.S.) was utilized to execute the analyses to address the research-based questions. The Cronbach alpha value for every research-based question was calculated to understand the reliability of the data. The data which were reliable were analyzed further to calculate mean, median, and standard deviations. Descriptive statistics were also provided for each of the questions of the survey. The categorical data were outlined using frequencies and percentages. To interpret the mean of Likert scale-based questions, the standard range was considered. (Strongly agree-Mean range: 4.20-5.00; Agree-Mean range: 3.40-4.19; Neutral-Mean range: 2.60-4.3.39; Disagree-Mean range:1.80-2.59; Strongly disagree-Mean range: 1.00-1.79).

## 3. Results and Discussion

### 3.1. Indian Biomedical Industry

#### 3.1.1. Demographic characteristics

The questionnaire was distributed between all pharmacy/biomedical engineering college students and pharma biomedical industrialists to understand the respondents’ perception of our proposed solutions for each stumbling problem faced by the Indian biomedical industry.

According to **Chart 1**, 64.13% of respondents fell in the age group of 21-30, followed by 21.59% of respondents in the age group of 31-40. **Table 1** depicts the frequency characteristics of the same.

**Chart 1.**
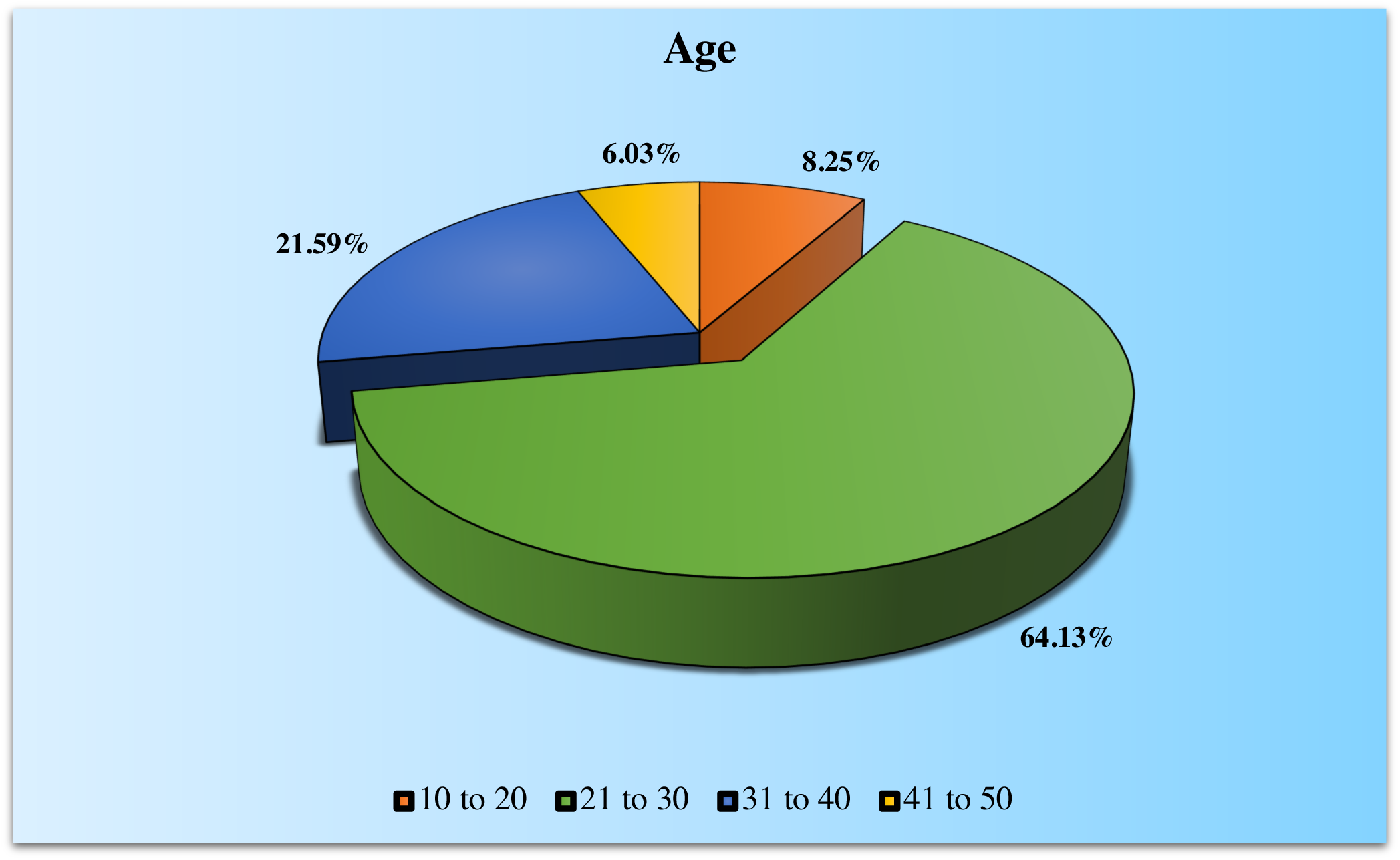
The age distribution of the respondents of the questionnaire survey.

**Table 1.** The frequency characteristics of the collected data from the respondents.

| Age |  |  |  |  |  |
| --- | --- | --- | --- | --- | --- |
|  |  | Frequency | Percent | Valid Percent | Cumulative Percent |
| Valid | 10-20 | 26 | 8.2 | 8.3 | 8.3 |
|  | 21-30 | 202 | 63.9 | 64.1 | 72.4 |
|  | 31-40 | 68 | 21.5 | 21.6 | 94.0 |
|  | 41-50 | 19 | 6.0 | 6.0 | 100.0 |
|  | Total | 315 | 99.7 | 100.0 |  |
| Missing | System | 1 | .3 |  |  |
| Total |  | 316 | 100.0 |  |  |

From **Chart 2** **and** **Table 2**, we observed that most of the respondents had Ph. D. (45.40%) as their educational qualification, followed by 29.21% of respondents who were post-graduated. Thus, most of the collected data is based on highly qualified respondents’ perceptions, leading to more reliability.

**Chart 2.**
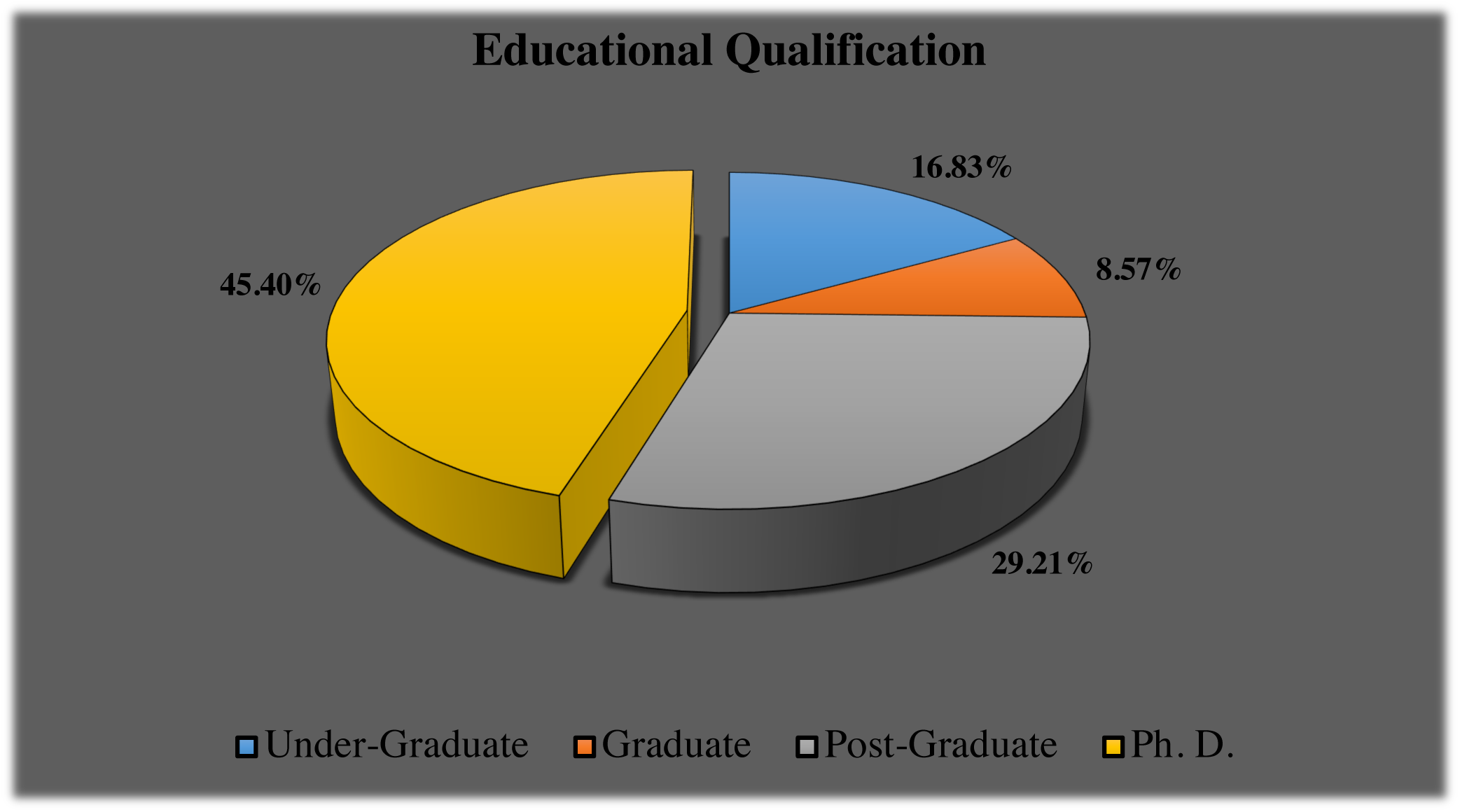
The educational qualification statistics of the respondents of the questionnaire survey.

**Table 2.** The frequency characteristics of the collected data from the respondents.

| Educational Qualification |  |  |  |  |  |
| --- | --- | --- | --- | --- | --- |
|  |  | Frequency | Percent | Valid Percent | Cumulative Percent |
| Valid | Under-Graduate | 53 | 16.8 | 16.8 | 16.8 |
|  | Graduate | 27 | 8.5 | 8.6 | 25.4 |
|  | Post-Graduate | 92 | 29.1 | 29.2 | 54.6 |
|  | Ph. D. | 143 | 45.3 | 45.4 | 100.0 |
|  | Total | 315 | 99.7 | 100.0 |  |
| Missing | System | 1 | .3 |  |  |
| Total |  | 316 | 100.0 |  |  |

#### 3.1.2. Understanding the knowledge of medical devices

Medical device means ‘any instrument, apparatus, implement, machine, appliance, implant, reagent for in vitro use, software, material or other similar or related article, intended by the manufacturer to be used, alone or in combination, for human beings, for one or more of the specific medical purpose(s) of:

- diagnosis, prevention, monitoring, treatment, or alleviation of disease,
- diagnosis, monitoring, treatment, alleviation of or compensation for an injury,
- the investigation, replacement, modification, or support of the anatomy or of a physiological process,
- supporting or sustaining life,
- control of conception,
- disinfection of medical devices
- providing information by means of in vitro examination of specimens derived from the human body; and does not achieve its primary intended action by pharmacological, immunological or metabolic means, in or on the human body, but which may be assisted in its intended function by such means [15].

This question was intended to determine whether the respondents were aware of the medical devices and their sectors.

From the observation (**Chart 3**), it is clearly understood that the targeted community for the questionnaire survey had the basic knowledge about medical devices (96.12%) and could proceed with other sections of the questionnaire. 1.29% of the respondents were unaware, and 2.59% of respondents were confused about the same.

**Chart 3.**
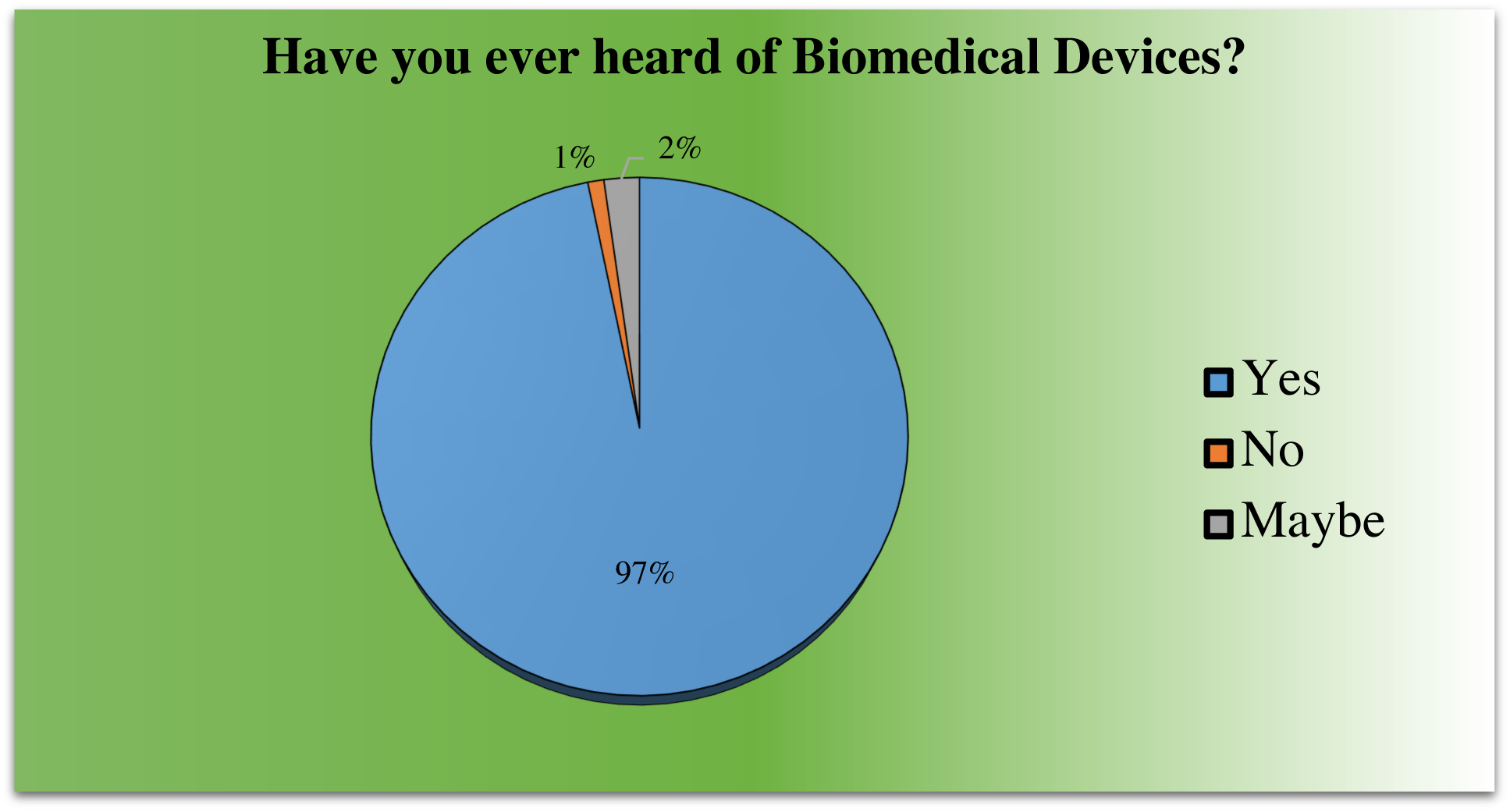
The percentage of respondents having knowledge on medical devices.

**Table 3** denotes the frequency characteristics of the same question calculated from the collected data of the respondents.

**Table 3.**
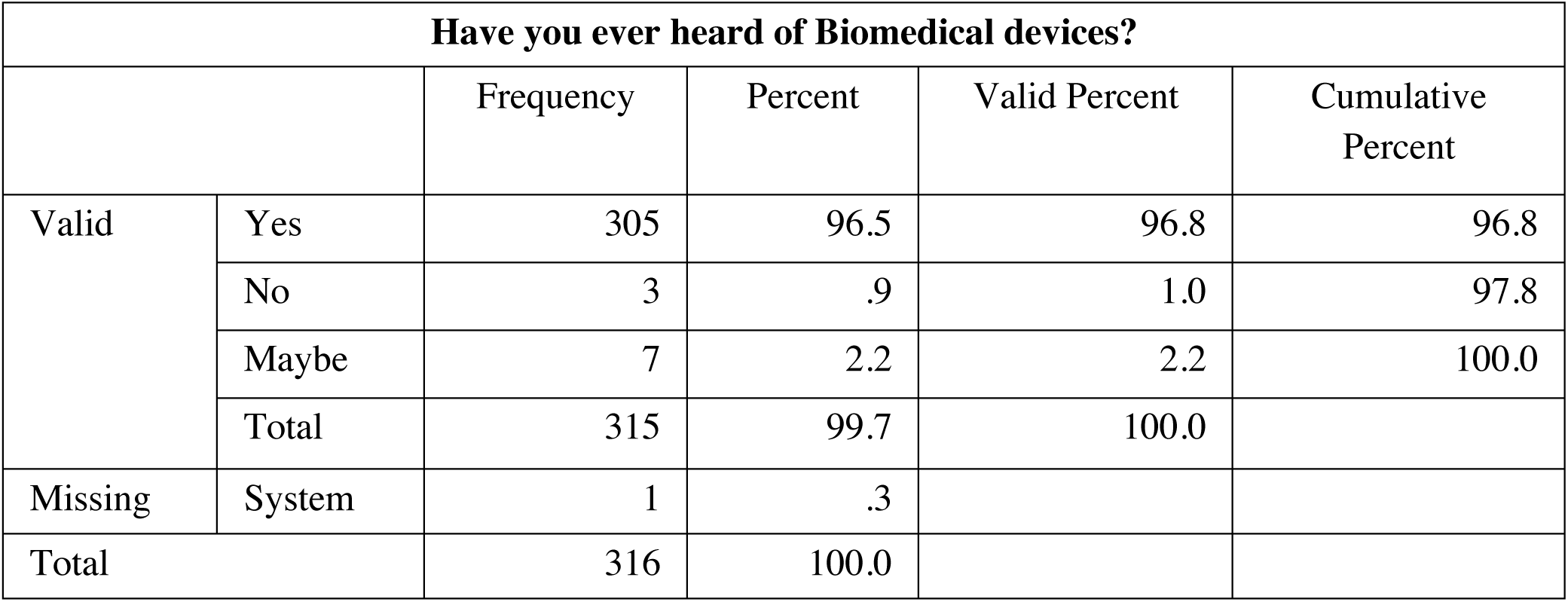
The frequency characteristics of the respondents’ data.

| Have you ever heard of Biomedical devices? |  |  |  |  |  |
| --- | --- | --- | --- | --- | --- |
|  |  | Frequency | Percent | Valid Percent | Cumulative Percent |
| Valid | Yes | 305 | 96.5 | 96.8 | 96.8 |
|  | No | 3 | .9 | 1.0 | 97.8 |
|  | Maybe | 7 | 2.2 | 2.2 | 100.0 |
|  | Total | 315 | 99.7 | 100.0 |  |
| Missing | System | 1 | .3 |  |  |
| Total |  | 316 | 100.0 |  |  |

We also tried to gain an in-depth understanding knowledge of the respondents on common standard medical devices used in the healthcare sector as well as daily life.

**Chart 4** depicted 71.2% of respondents were mostly aware of CT Scanner as one of the medical devices followed by Hot Air Oven (68.7%), Infusion Pump (64.6%), Weighing machines (58.2%), and Xray Scanners (54.1%). Thus, except for one respondent, everyone was aware of multiple standard biomedical devices. For other devices, too, more than half of the respondents were aware.

**Chart 4.**
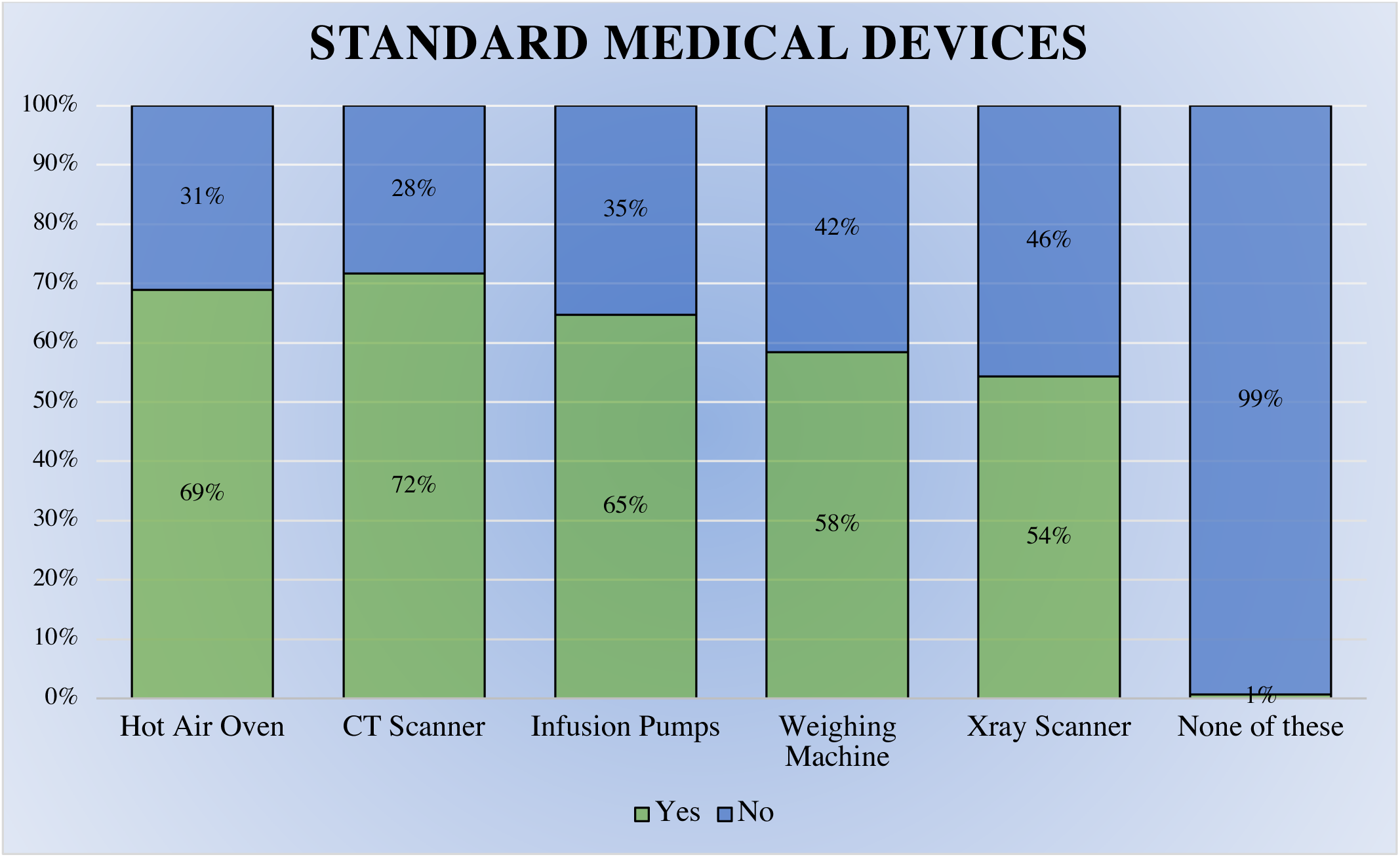
The percentage of responses recorded for each option from the respondents.

The frequency characteristics for the same question are depicted in **Fig 1**.

**Fig 1.**
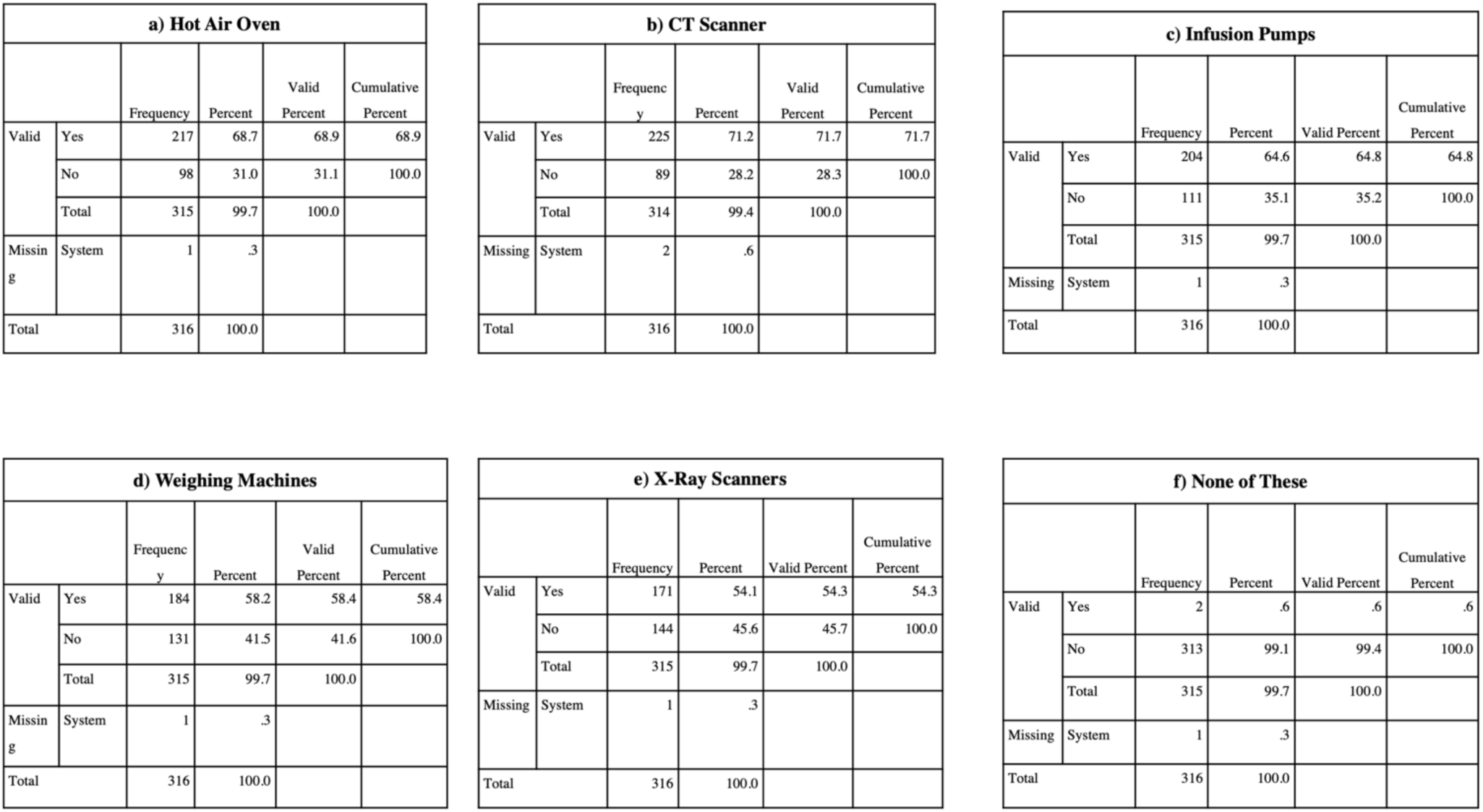
The frequency characteristics of a) Hot air oven b) CT scanner c) Infusion pumps d) Weighing machines e) X-Ray scanners f) None of these calculated from the responses collected from the questionnaire survey.

#### 3.1.3. Awareness of the respondents concerning the Indian medical industry

The Indian medical devices industry has a probability of leapfrogging novelty combining physical devices and incorporating digital infrastructure for long-term innovation. Medical devices in India are now deemed to perform an essential role in delivering quality health care to the people. The initiation of MDR 2017 was a substantial breakthrough in the establishment of a powerful platform. The awareness around the need to have a robust medical devices ecosystem in the country is gaining traction resulting in higher growth rates for India as compared to the global industry.

The objective was to determine if respondents, mostly from the biomedical fraternity, know any top Indian medical devices manufacturing company.

According to **Chart 5**, 83.2 % of respondents are mostly aware of Indian companies manufacturing medical devices, and 8% of respondents are unsure about the 1same.

**Chart 5.**
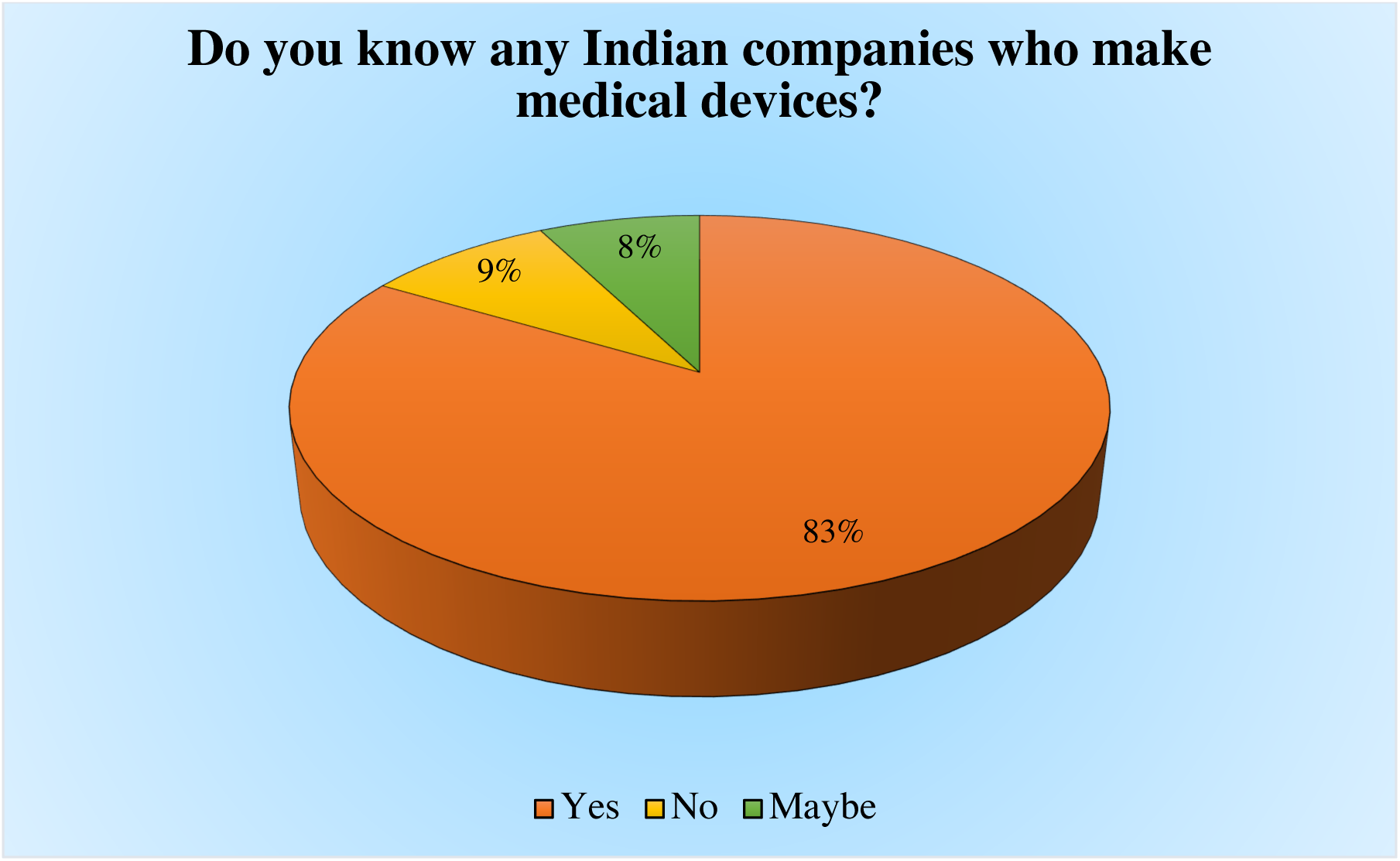
The percentage of responses recorded concerning the awareness of respondents on Indian medical devices manufacturing companies.

**Table 4** depicts the frequency characteristics of the same data collected from the respondents.

**Table 4.** The frequency characteristics were calculated from the respondents’ data.

| Do you know any Indian companies that make medical devices? |  |  |  |  |  |
| --- | --- | --- | --- | --- | --- |
|  |  | Frequency | Percent | Valid Percent | Cumulative Percent |
| Valid | Yes | 263 | 83.2 | 83.5 | 83.5 |
|  | No | 28 | 8.9 | 8.9 | 92.4 |
|  | Maybe | 24 | 7.6 | 7.6 | 100.0 |
|  | Total | 315 | 99.7 | 100.0 |  |
| Missing | System | 1 | .3 |  |  |
| Total |  | 316 | 100.0 |  |  |

Respondents were mostly aware of the names of the companies manufacturing medical devices in India. This question doesn’t confirm whether the companies they think to be Indian are Indian companies or not. The next question throws light on that.

We also tried to investigate the respondents’ awareness about the companies manufacturing medical devices in India. Some multinational companies that made their products popular in India may be considered Indian companies in public perception.

Based on the data acquired, depicted in **Chart 6** and **Fig 2**, an average of 62.10% of respondents correctly identified the two Indian companies (Centenial Surgical Suture Ltd. and CDR Medical Industries Ltd), and about 50.08% of respondents wrongly identified two foreign companies, Medtronic and Phillips Healthcare, as Indian companies. For the other foreign company, Johnson & Johnson, people are more aware as 77% of respondents identified it as a foreign company.

**Chart 6.**
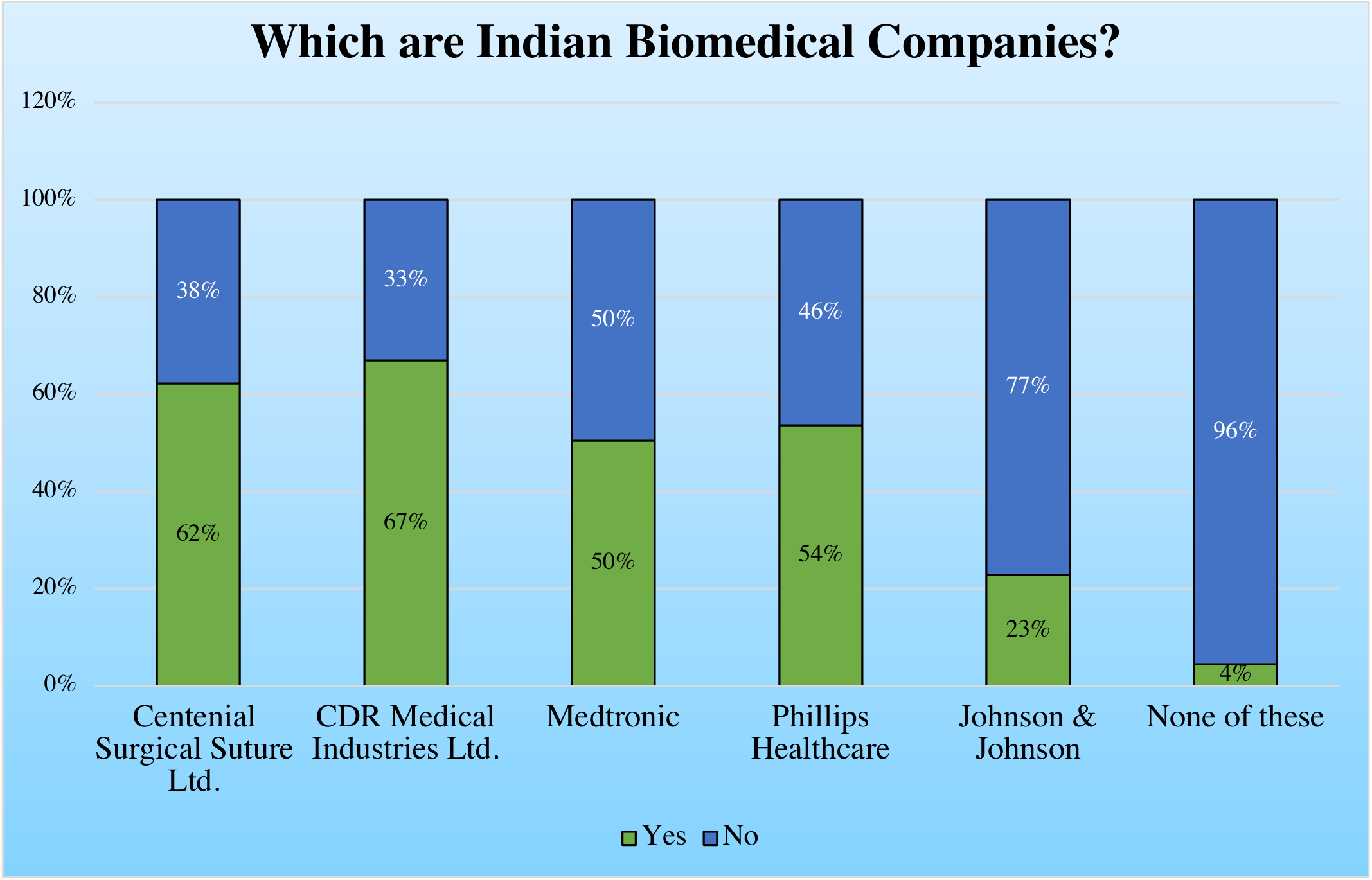
The percentage of responses estimated for each of the standard Indian medical device companies.

**Fig 2.**
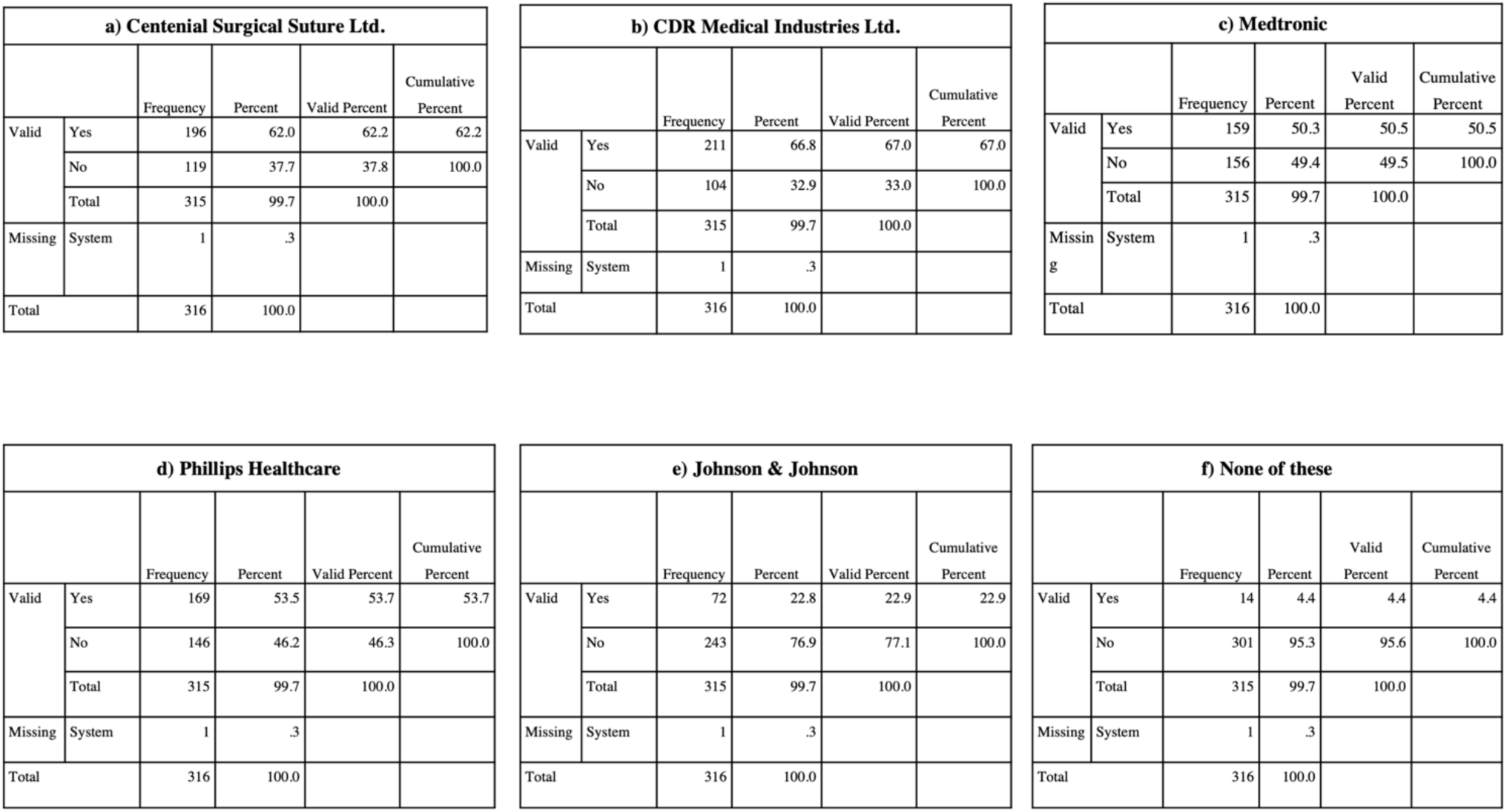
The frequency characteristics of a) Centenial Surgical Suture Ltd. b) CDR Medical Industries Ltd. c) Medtronic d) Philips Healthcare e) Johnson and Johnson f) None of these estimated from the data acquired from the respondents.

Thus, we can conclude that the Indians are less aware of Indian medical device manufacturing companies and are more inclined towards the wrong perception of considering foreign companies as Indian ones.

#### 3.1.4. Perception of the steps taken by the Indian government to enhance the Indian medical industry

In 2020, the Government of India brought a few new regulations/schemes for the benefit of the medical devices sector in India. With the revolutionary consciousness and significance brought to this fundamental sector, the government, along with the respective officials, have taken various measures in the right direction and implemented laws for the amelioration of the industry. Medical devices are now categorized as drugs and have levels of risk and stringent regulation, which also have incentives and benefits depending on the type of branch of healthcare the biomedical device works on. Such initiatives are exactly what we need to bring change and enhance the sector.

On this positive note, we look into the several steps the Government of India has taken to ensure the development of a vibrant ecosystem of medical device manufacturing in India over the past few years, such as the ones mentioned previously. These include acts such as The Medical Devices Amendment Rules of 2020, bring all medical devices in India under regulation as drugs. Prior to this amendment, only 37 classes of medical devices were strictly regulated by the regulatory bodies, and changes in the Indian law caused amends to include all medical devices, and the Indian law that governs quality and safety of medical devices will now apply to all medical devices, effective April 1, 2020 [4]. Under the ‘Productions Linked Incentives Scheme for Medical Devices, 2020’ scheme, the government provides incentive at the rate of 5% of incremental sales on the initial year 2019-20, which will be applicable to the segments of medical devices identified under the categories such as radiotherapy and cancer care products, Anaesthetics & Cardio-Respiratory medical devices and Imaging medical devices (both ionizing & non-ionizing radiation products) and Nuclear Imaging Devices. Aimed at reducing the manufacturing cost of medical devices and equipment produced in the country, there was funding for ‘Medical Devices Parks’ in the country, 2020. They work by financing infrastructure facilities in 4 medical device parks with financial aid and grants of Rs. 400 crores, with the maximum aid being Rs.100 crore to each located intricately across the country in major central hubs [4].

This question intends to investigate the awareness about these initiatives among the people related to this sector.

It has been observed in **Chart 7** and **Fig 3**, the majority of the respondents are unaware of the initiatives taken by the Government of India in 2020. On average, only about 9-15% of respondents are aware of each of these initiatives, and 40% of respondents know about all the government initiatives taken to boost up the medical device industry.

**Chart 7.**
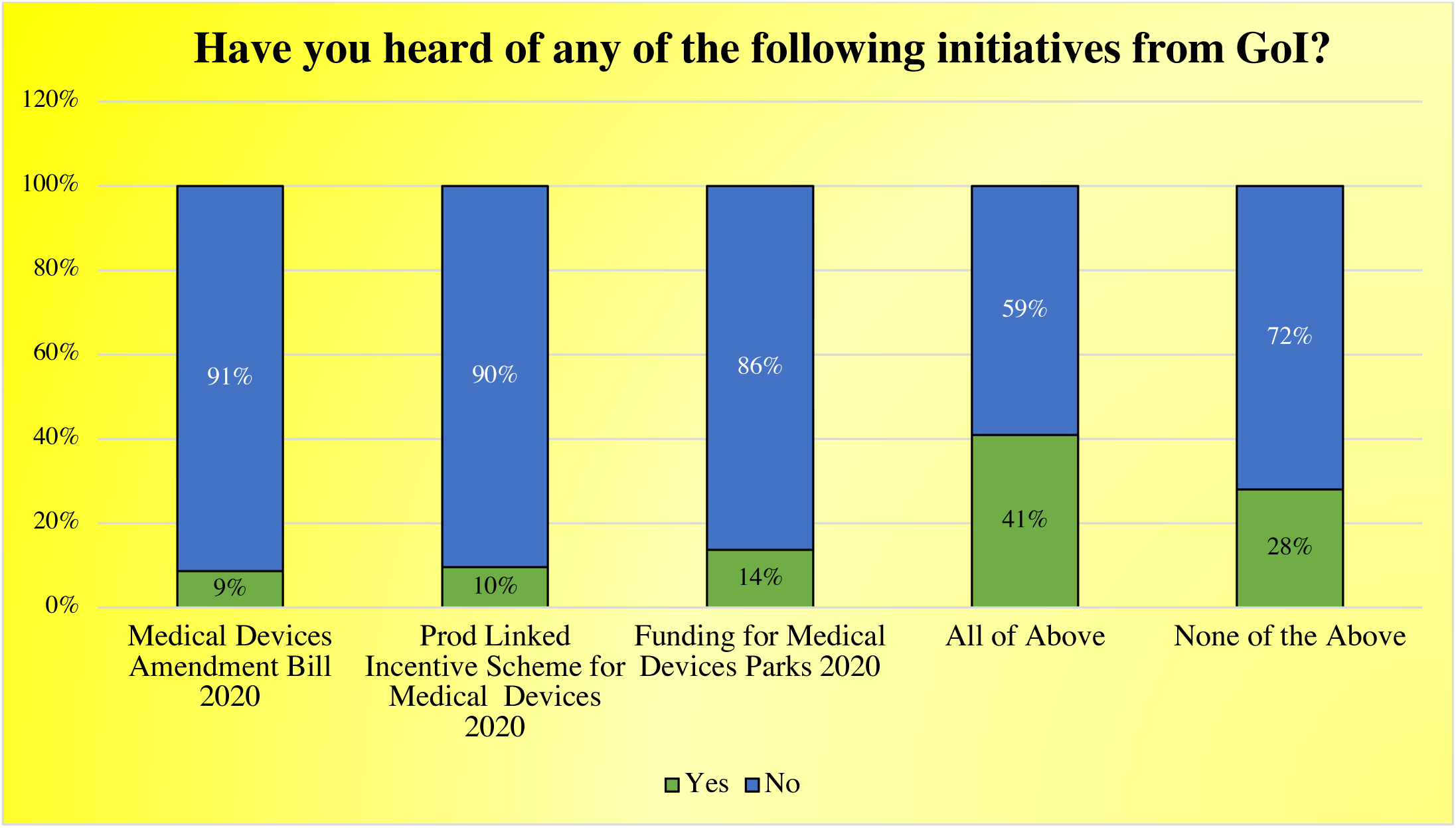
The percentage of responses recorded for each initiative taken by the government based on the data of respondents.

**Fig 3.**
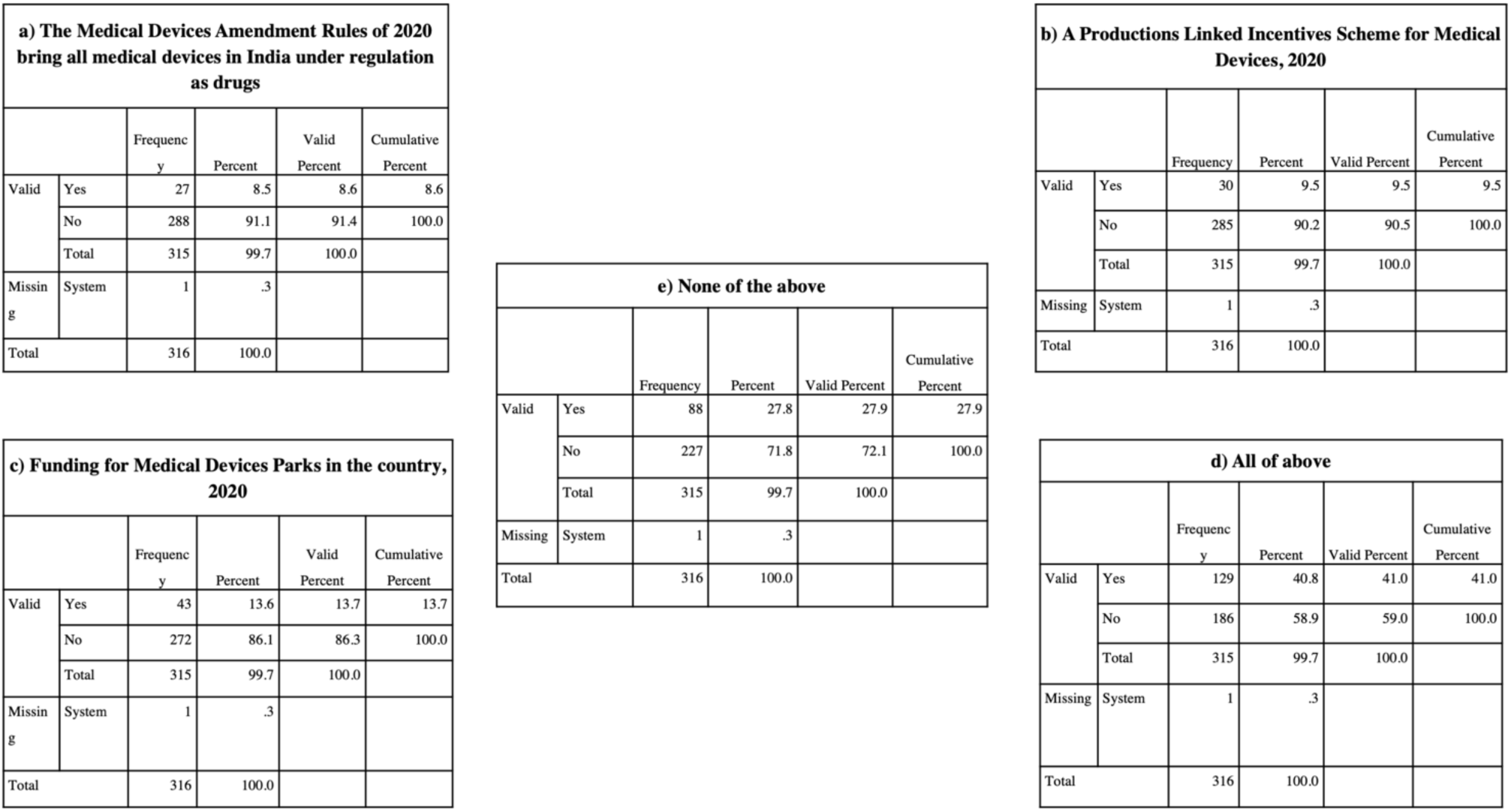
The frequency characteristics of a) The medical devices amendment rules of 2020 that bring all medical devices in India under regulation as drugs b) A production linked incentives scheme for medical devices, 2020 c) Funding for medical devices parks in the country, 2020 d) All of the above e) None of the above estimated from the respondents’ data.

2020 was the year of COVID-19 when a lot of small indigenous biomedical device manufacturers had risen to the occasion and helped in handling the pandemic. To support them, the government has put them under regulations (like drugs) and, on the other hand, gave this industry some benefits through some initiatives. But there was not enough dissemination of knowledge/awareness about these initiatives. Unless the awareness increases, it will not give enough boost to the medical devices manufacturing sector.

#### 3.1.5. Regulatory standards of Indian medical devices

The question was intended to get the respondents’ view (who are aware of the Biomedical Devices manufacturing industry in India) about the need for a Licensing Authority to regulate this industry. 75-80% of all supplies for Biomedical Devices are imported from foreign countries. To boost the indigenous manufacturers in India, it is imperative to have a regulatory authority like the Food and Drugs Administration (FDA) of the USA or the European Medicines Agency (EMA) of the EU.

From **Chart 8** and **Table 5**, we observe that a whopping 94% (majority) of respondents believe that such a regulatory authority is needed to control the Indian medical devices manufacturing industry.

**Chart 8.**
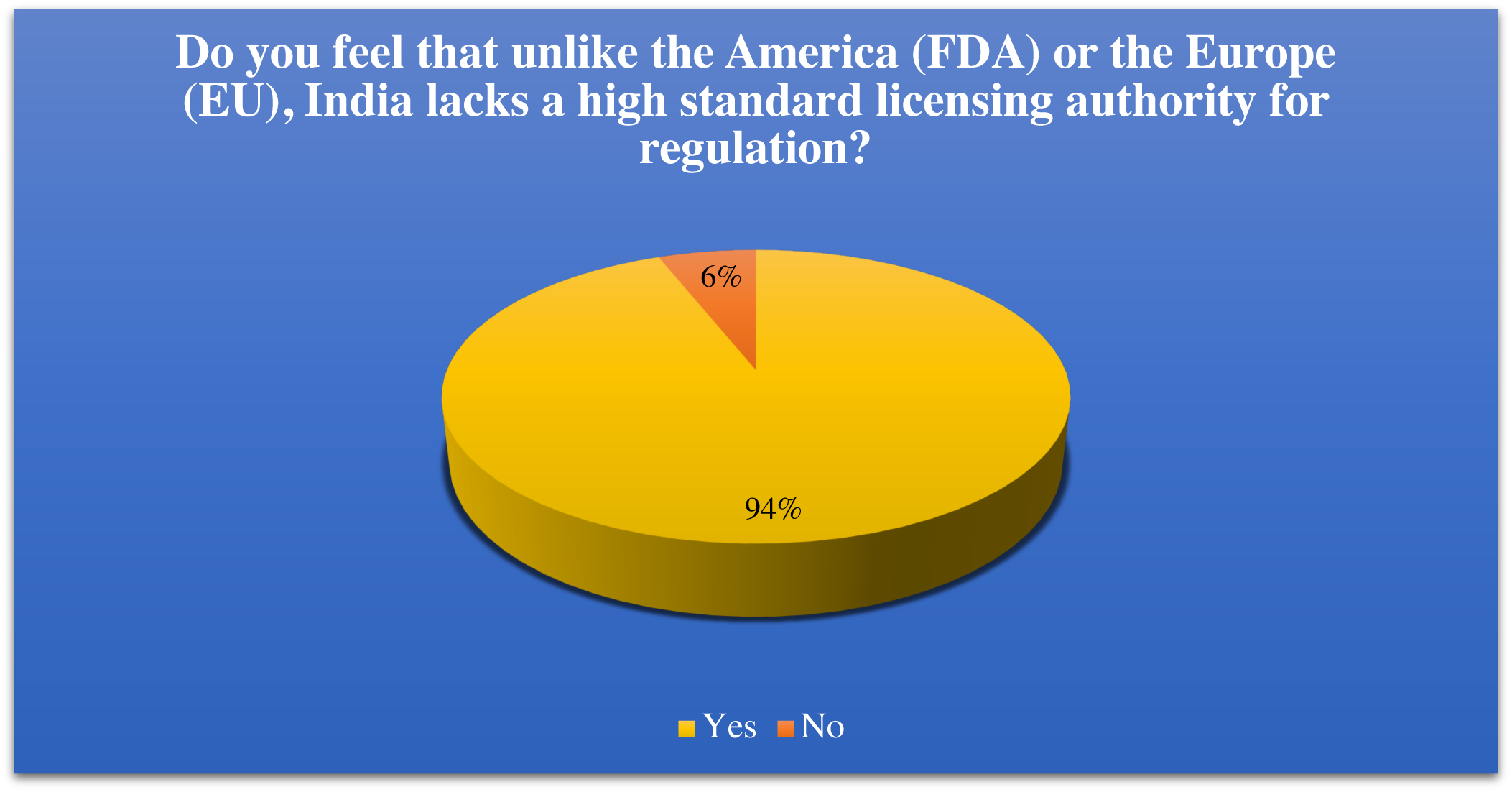
The percentage of responses agreeing and disagreeing on the fact that Indian lacks standard licensing authority in contrast to the US and Europe.

**Table 5.** The frequency characteristics of the data collected from the respondents.

| **Do you feel that unlike America (FDA) or Europe (EU), India lacks a high standard licensing authority for regulation?** |  |  |  |  |  |
| --- | --- | --- | --- | --- | --- |
|  |  | Frequency | Percent | Valid Percent | Cumulative Percent |
| Valid | Yes | 296 | 93.7 | 94.0 | 94.0 |
|  | No | 19 | 6.0 | 6.0 | 100.0 |
|  | Total | 315 | 99.7 | 100.0 |  |
| Missing | System | 1 | .3 |  |  |
| Total |  | 316 | 100.0 |  |  |

This is a positive sign that many respondents are aware of our current regulatory bodies’ shortcomings and limitations and the strict implementation of their standards. Thus, people see a reason for bringing about change and implementing a higher standard licensing authority as the Food and Drugs Administration (FDA) or European Medicines Agency (EMA).

We also tried to investigate the ways to improve the quality management (ISO 13485) and the endurance testing (ISO 10651) implementation on medical devices in the Indian biomedical sector. The main intention was to understand the perception of the respondents on the proposed solutions to enhance the quality management system and the endurance testing of the medical devices manufactured in India.

ISO 13485 is a standard related to the quality management system (QMS). The current version of this guideline is ISO 13485:2016 specifies requirements for QMS where an organization needs to showcase its ability to offer medical devices and related services that consistently meet client and applicable regulatory conditions. ISO 13485 is based on the ISO 9001 Process model concepts of Plan, Do, Check, Act intended for regulatory compliance. Participation of such organizations can be at any phase of the life cycle, which comprises production, installation, design, and development or servicing of a medical device and design development or provision of related activities (for example, technical support). This standard is also usable by suppliers or external parties that provide products, including QMS related services to such organizations [16] [17].

ISO 13485 certification mainly helps to improve overall performance, increase market reach & exclude unreliability. It helps to achieve the following objectives:

- Increase efficiency cut monitor performance and cost
- Outline how to review the processes
- Meet regulatory conditions and customers’ expectations
- Demonstrate that you produce safer medical devices

The main classic feature of this standard is its applicability to organizations, irrespective of their type and size, except where clearly stated. If applicable regulatory requirements permit leaving out of designing & development protocols, which can be used to explain their exclusion from QMS. These can be served as alternative tactics that can be addressed to QMS. It is the important responsibility of the organization to make sure that claims of conformity to this standard reflect any exclusion of design and development controls [16] [17].

ISO 10651 consists of six parts, under the general title Lung ventilators for medical use — Particular requirements for basic safety and essential performance. These parts of ISO 10651 specify the basic safety and essential performance requirements for gas-powered emergency resuscitators planned for use with humans by first responders. This equipment is intended for emergency field use and is planned to be continuously operator attended in everyday use [18].

**Table 6** depicts the value of central tendency calculated from the respondents’ data. All the respondents have agreed to the proposed solutions for enhancing the QMS and the endurance testing of Indian medical devices.

**Table 6.** The measures of central tendency.

| Statistics |  |  |  |  |  |
| --- | --- | --- | --- | --- | --- |
|  |  | Stringent laws and implementation | Formation of strong certification authority | Having a knowledgeable and skilled workforce | Proper funding from government along with adequate testing facilities |
| N | Valid | 315 | 315 | 315 | 315 |
|  | Missing | 1 | 1 | 1 | 1 |
| Mean |  | 3.9143 | 3.9079 | 4.1714 | 4.0921 |
| Median |  | 4.0000 | 4.0000 | 4.0000 | 4.0000 |
| Std. Deviation |  | .87914 | .97473 | .90776 | .93129 |

To determine the reliability of data, we performed the Cronbach Alpha test, which resulted in a coefficient of alpha of 0.802, providing us the way for further data analysis (alpha value-0.6-1 is reliable).

The frequency characteristics of each possible solution to improve ISO 13485 and ISO 10651 are described in **Fig 4** and **Fig 5**.

**Fig 4.**
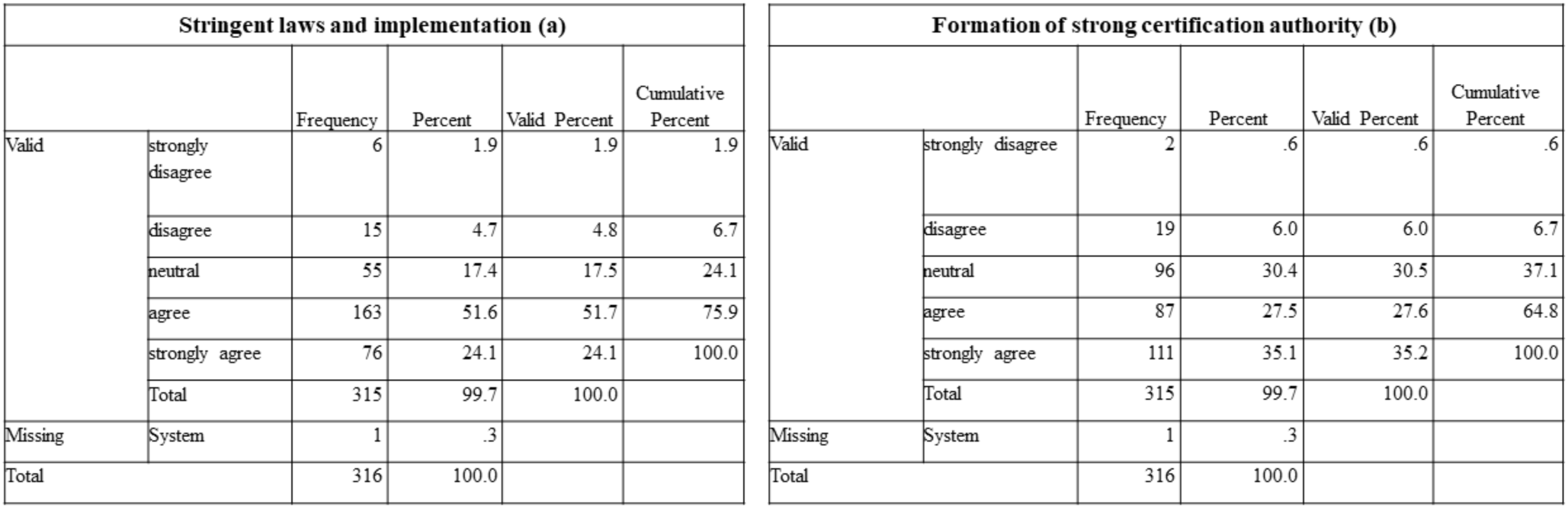
The frequency characteristics of a) Stringent laws and implementation b) Formation of strong certification authority calculated from the respondents’ data.

**Fig 5.**
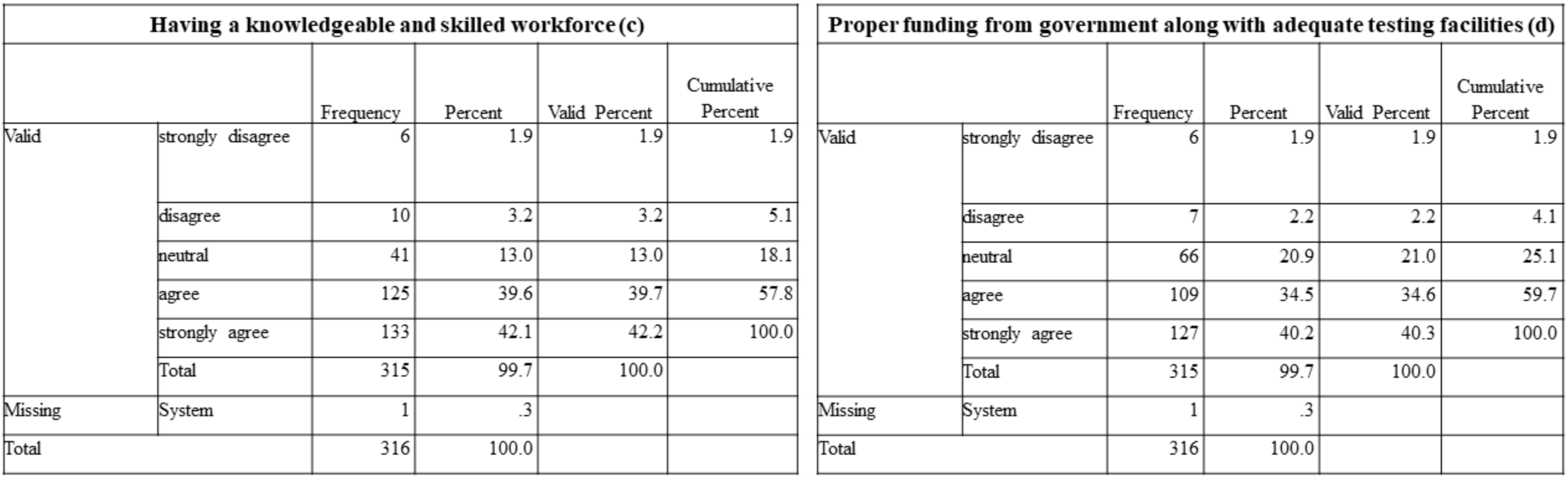
The frequency characteristics of c) Having a knowledgeable and skilled workforce d) Proper funding from the government along with adequate testing facilities estimated from the respondents’ data.

As already discussed, stringent laws and their implementation lack from the government’s side, which results in poor manufacturing of medical devices. 51.6% of respondents agreed that the implementation of strong and strict laws might improve quality management and endurance testing, making it the superior solution from all the other possible solutions.

Also, a skilled workforce (by taking into account poor literacy rate and less availability of biomedical device industry personnel) is essential in every case, whether to improve the manufacturing or to enhance the quality testing in the Indian medical devices industry. For the same, 42.1% of respondents strongly agreed that skilled personnel recruitment might improve the quality management and the endurance testing of the medical devices manufactured in India.

Also, 40.2% of respondents strongly agreed that the government should uplift the indigenous medical device sector by offering fundings and appropriate testing facilities (considered based on the availability of money and the damaged ecosystem infrastructure, for raising money on their own for qualitative checking of devices providing the facility for the same).

A strong certification body (by taking into account the ‘Aatmanirbhar’ movement of India and thinking of setting standards for our own country which can be accepted worldwide. This can increase both ease of implementation and market opportunities for native organizations) should be present to avoid the production of ineffective, hazardous medical devices, to improve consumer protection, eliminating trading of below standard devices, etc. Thus, the establishment of the same might improve the quality and the endurance testing of Indian medical devices, as strongly agreed by 35.1% of respondents.

Overall results (**Chart 9**) demonstrated a need to improve the quality management and endurance testing for the implementations in the medical device sector, and most of the respondents agreed with the proposed solutions.

**Chart 9.**
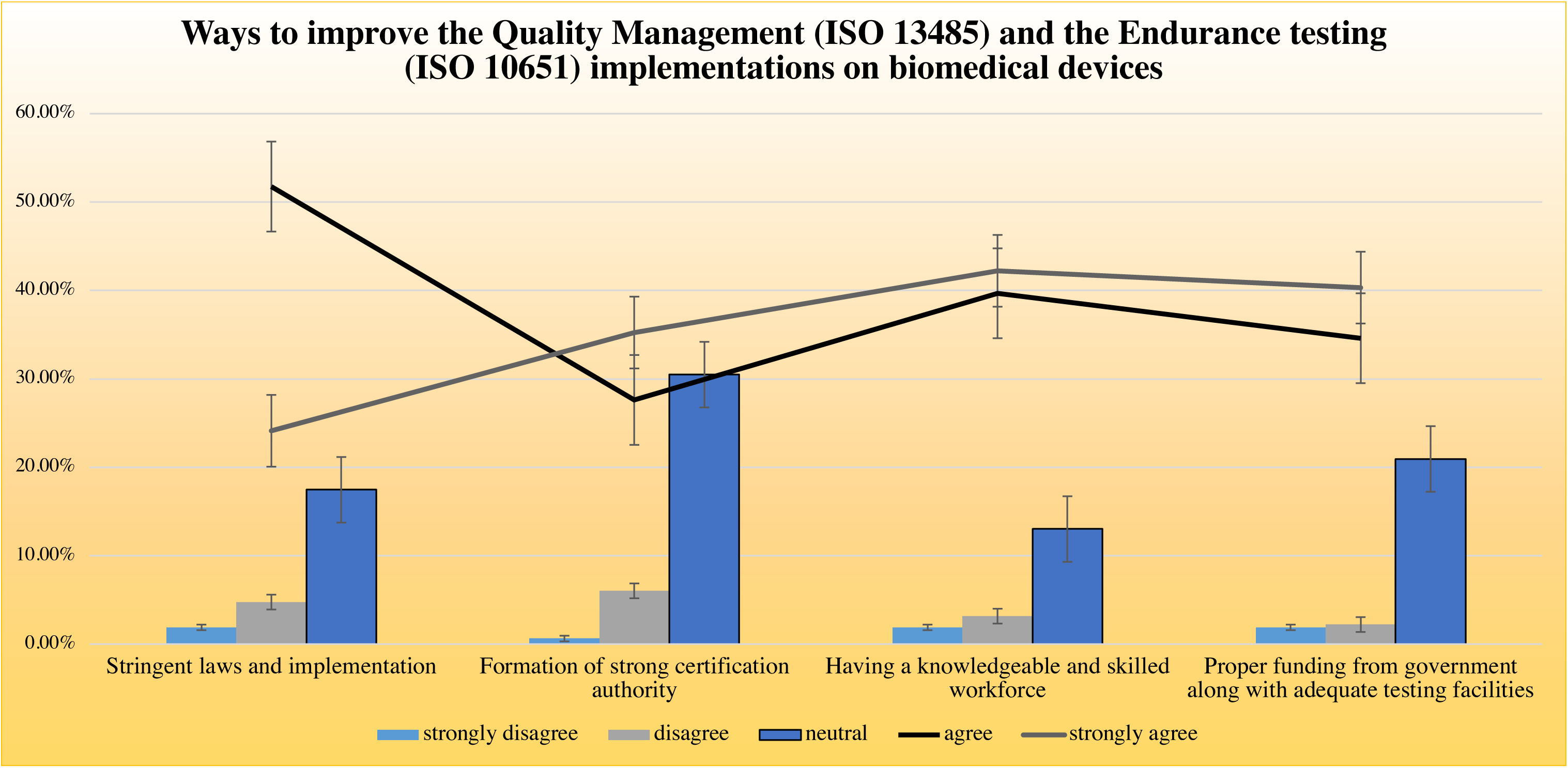
The percentage of responses recorded for each possible solution to advance the implementation of ISO 13485 and ISO 10651 on medical devices.

ISO 14971-The risk management system aims to help manufacturers of medical devices recognize the hazards of the medical device, estimate and evaluate the related risks, and monitor the controls’ effectiveness.

‘ISO 14971:2019 Medical devices-Applications of risk management to medical devices guidelines specify terminologies, principles, and a process of risk management of medical devices comprising software as a medical device and in vitro diagnostic medical devices. The most important annexes in this guideline as follows:

(1) Annex A: Identifications of Hazards and characteristics of safety
(2) Annex B: Risk analysis techniques
(3) Annex C: Risk acceptability conditions
(4) Annex D: Information on safety and information on residual risk
(5) Annex E: Role of international safety standards in the risk management
(6) Annex F: Guidance on risk related to cyber and data security
(7) Annex G: Components and devices not designed using ISO 14971 (new)
(8) Annex H: Guidance on in-vitro diagnostic medical devices (new) [19]

The requirements of this standard are applicable to all phases of the life cycle of medical devices. The process explained in this standard appropriate to risks associated with a medical device, for example, risks linked with biocompatibility, radiation, electricity, data & systems security, moving parts, and usability. This process also can be applied to products, not essentially medical devices in some conditions, and can also be used by others concerned with the overall medical device life cycle [19].

The question intends to define the possible ways to improve the risk management implementation of biomedical devices.

To determine the reliability of data, we performed the Cronbach Alpha test, which resulted in a coefficient of alpha of 0.789, providing us the way for further data analysis (alpha value-0.6-1 is reliable).

**Table 7** describes the values of the central tendency calculated from the respondents’ data, and all the respondents have agreed with the probable solutions to improve the implementation of ISO 13485 on medical devices.

**Table 7.** The measures of central tendency.

| Statistics |  |  |  |  |  |
| --- | --- | --- | --- | --- | --- |
|  |  | Safety testing and strict regulations | Development of an effective risk management plan for each device | Post production analysis of marketed devices | Recruitment of expertees in design of clinical studies for validation |
| N | Valid | 315 | 315 | 315 | 315 |
|  | Missing | 1 | 1 | 1 | 1 |
| Mean |  | 3.8603 | 3.8540 | 4.0984 | 4.1079 |
| Median |  | 4.0000 | 4.0000 | 4.0000 | 4.0000 |
| Std. Deviation |  | .88125 | .95983 | .94084 | .93470 |

The frequency characteristics of each of the probable solutions to strengthen the quality management system and the endurance testing are described in **Fig 6** and **Fig 7**.

**Fig 6.**
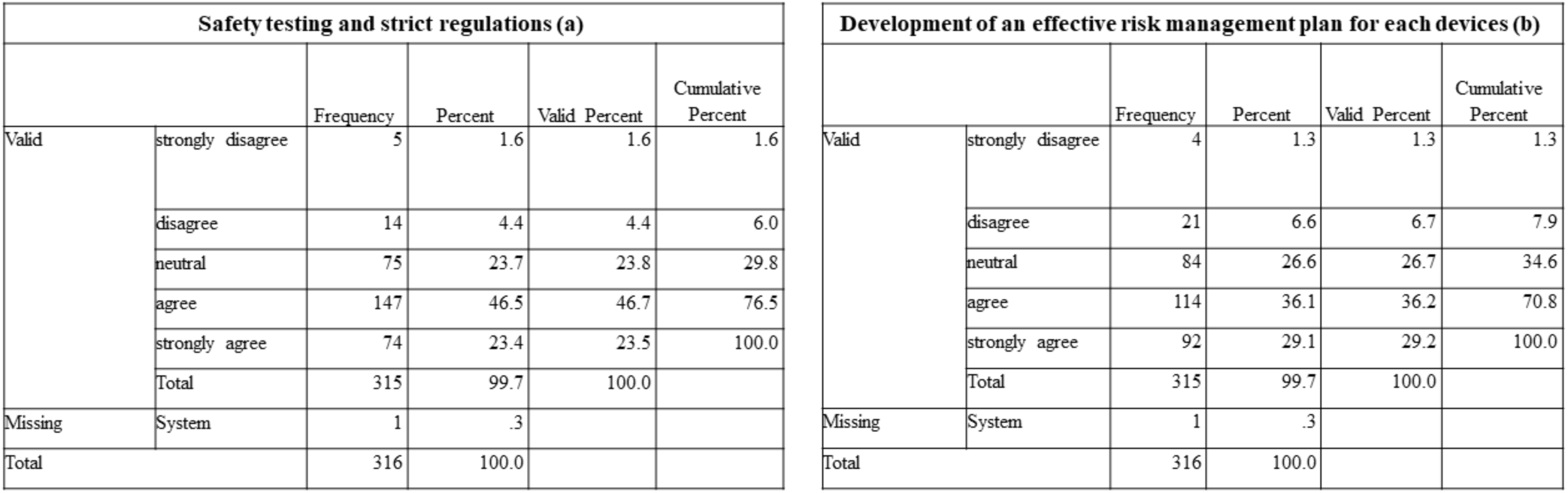
The frequency statistics of a) Safety testing and strict regulations b) Development of an effective risk management plan for each device was calculated from the respondents’ data.

**Fig 7.**
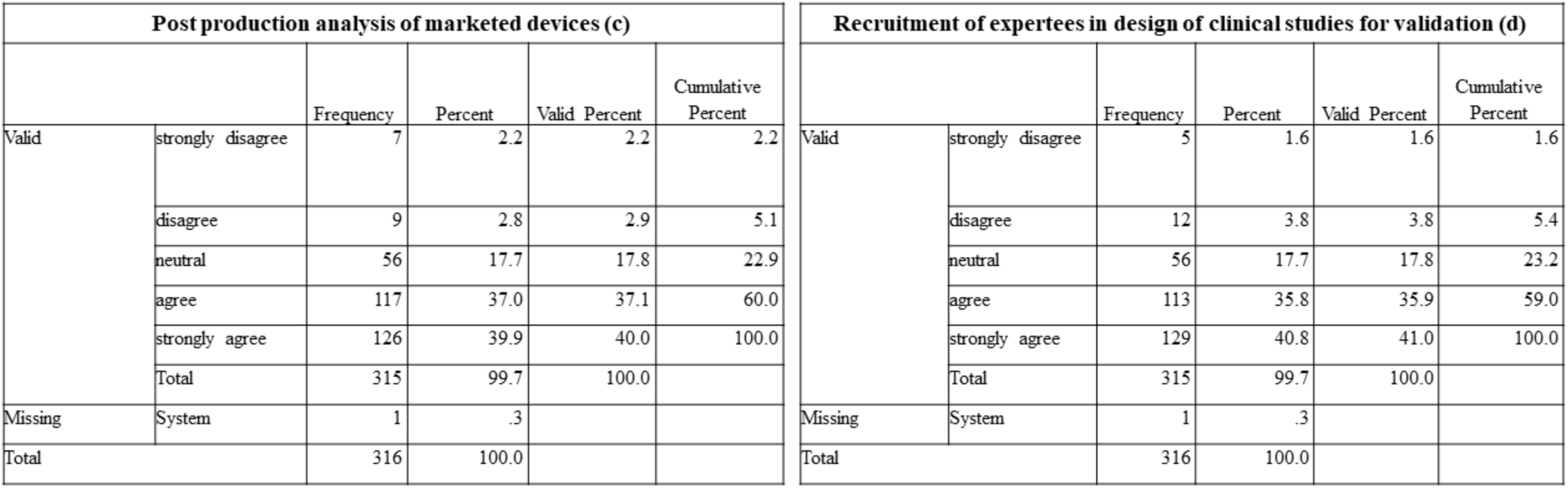
The frequency statistics of c) Post-production analysis of marketed devices d) Recruitment of expertees in the design of clinical studies for validation estimated from the data of respondents.

46.5% of respondents agreed that safety testing and implementation of strict regulations might improve the risk management system of medical devices manufactured in India.

The post-production analysis consists of four sections: a) to set up a system to assemble and review appropriate production and post-production data b) to gather relevant data for the medical device under examination c) review if the information is appropriate to the medical device safety d) if any of the aforementioned situations take place, the manufacturer needs to take measures [20]. 39.9% of respondents strongly agreed that post-production analysis is obligatory to enhance the risk management system.

A risk management plan is a record that a project manager develops to anticipate risks, evaluate impacts, and determine responses to risks. The plan consists of identifying risk, assessing risk, a potential treatment of risk, creating strategy, implementing, reviewing, and evaluating the plan [21]. 36.1% of respondents agreed that developing a proper risk management plan might avoid manufacturing faulty and sub-standard medical devices.

Designing clinical studies for medical device validation requires expertees for better productive results. Thus, lack of a skilled workforce and deficiency of adequate knowledge are among the major causes for the failure of the risk management system. Therefore, 40.8% of respondents strongly agreed that the recruitment of professional, knowledgeable expertees might improve the risk management system leading to the manufacturing of useful medical devices.

Overall results (**Chart 10**) proved that risk management should be improved, and the respondents also agreed on the possible solutions that can be implemented to improve this risk management on medical devices.

**Chart 10.**
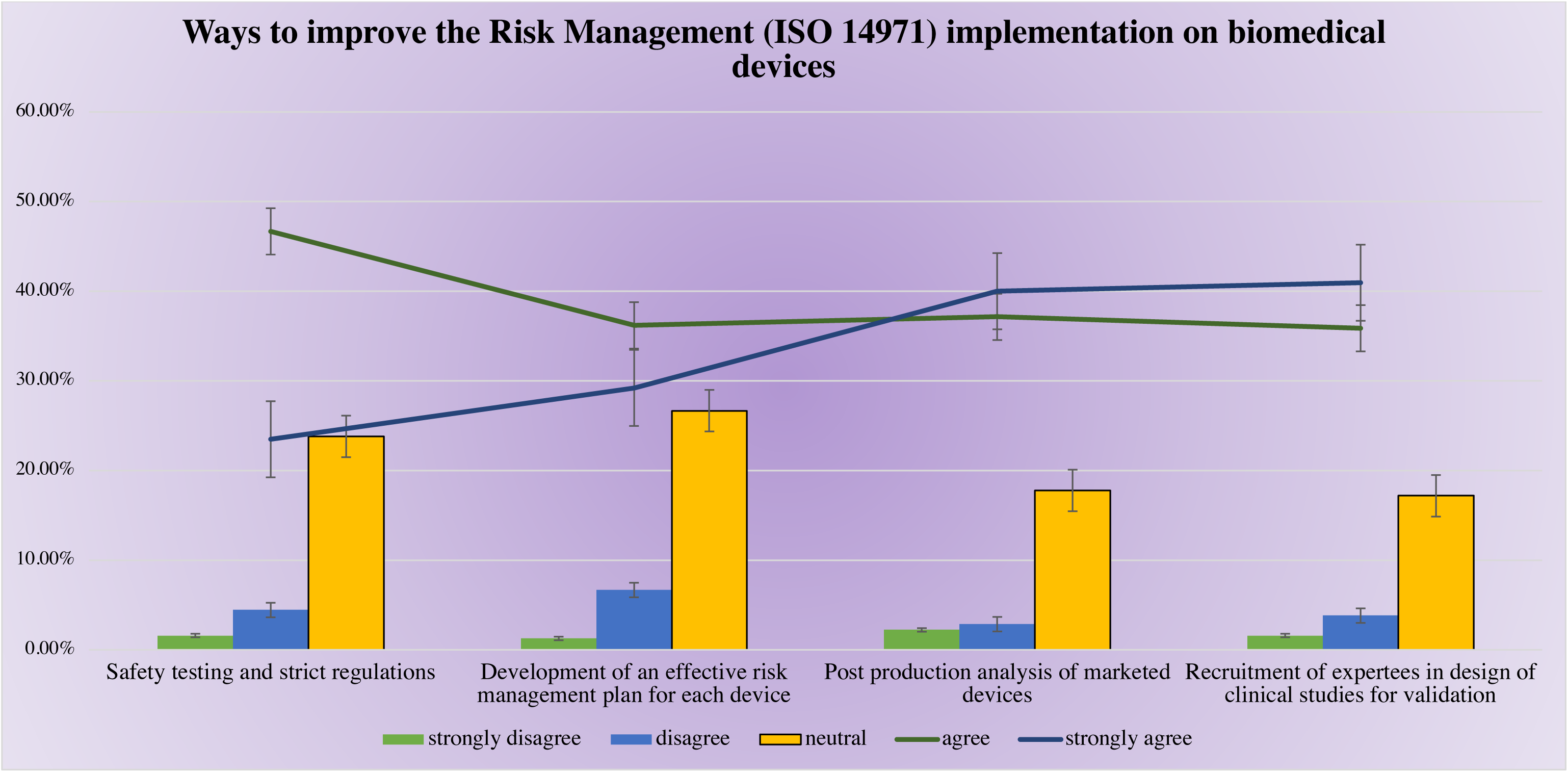
The percentage of responses calculated for each possible solution to implement ISO 14971 effectively on Indian medical devices.

#### 3.1.6. Current limitations in the Indian medical devices industry

There are some difficulties that the Indian medical device industry is confronting, like regulatory/legal issues, unfavorable duty structure for both imports and exports, establishing affordable products, rising labor productivity skills, elevated capital cost, and consent delays [22]. Compared to others, like the US and the EU, the concerns in India vary explicitly in areas such as design systematization/localization of devices, regulatory standards to meet the original equipment manufacturer brand, in-country production, and dissemination.

The question was intended to find out the perception of respondents on the major cause of limitations faced by the medical devices sector.

To determine the reliability of data, we performed the Cronbach Alpha test, which resulted in a coefficient of alpha of 0.746, providing us the way for further data analysis (alpha value-0.6-1 is reliable).

**Table 8** discusses the measures of central tendency estimated from the data of the respondents. The participants have agreed to most of the possible reasons causing a setback for the Indian medical devices industry. The respondents remained neutral for ‘Limited domestic demand’ revealed after the interpretation of mean.

**Table 8.** The measures of central tendency.

| Statistics |  |  |  |  |  |  |
| --- | --- | --- | --- | --- | --- | --- |
|  |  | Unfavorable duty structure | Limited domestic demand | Lack of Laws and regulations | Deficiency of skilled personnel | Lack of collaboration among govt. and industry |
| N | Valid | 315 | 315 | 315 | 315 | 315 |
|  | Missing | 1 | 1 | 1 | 1 | 1 |
| Mean |  | 3.6921 | 3.4984 | 3.9429 | 3.8063 | 3.9619 |
| Median |  | 4.0000 | 3.0000 | 4.0000 | 4.0000 | 4.0000 |
| Std. Deviation |  | .82387 | 1.07483 | 1.02045 | .99871 | 1.01821 |

**Fig 8** and **Fig 9** depict the frequency characteristics of all the possible causes of limitations calculated from the respondents’ data.

**Fig 8.**
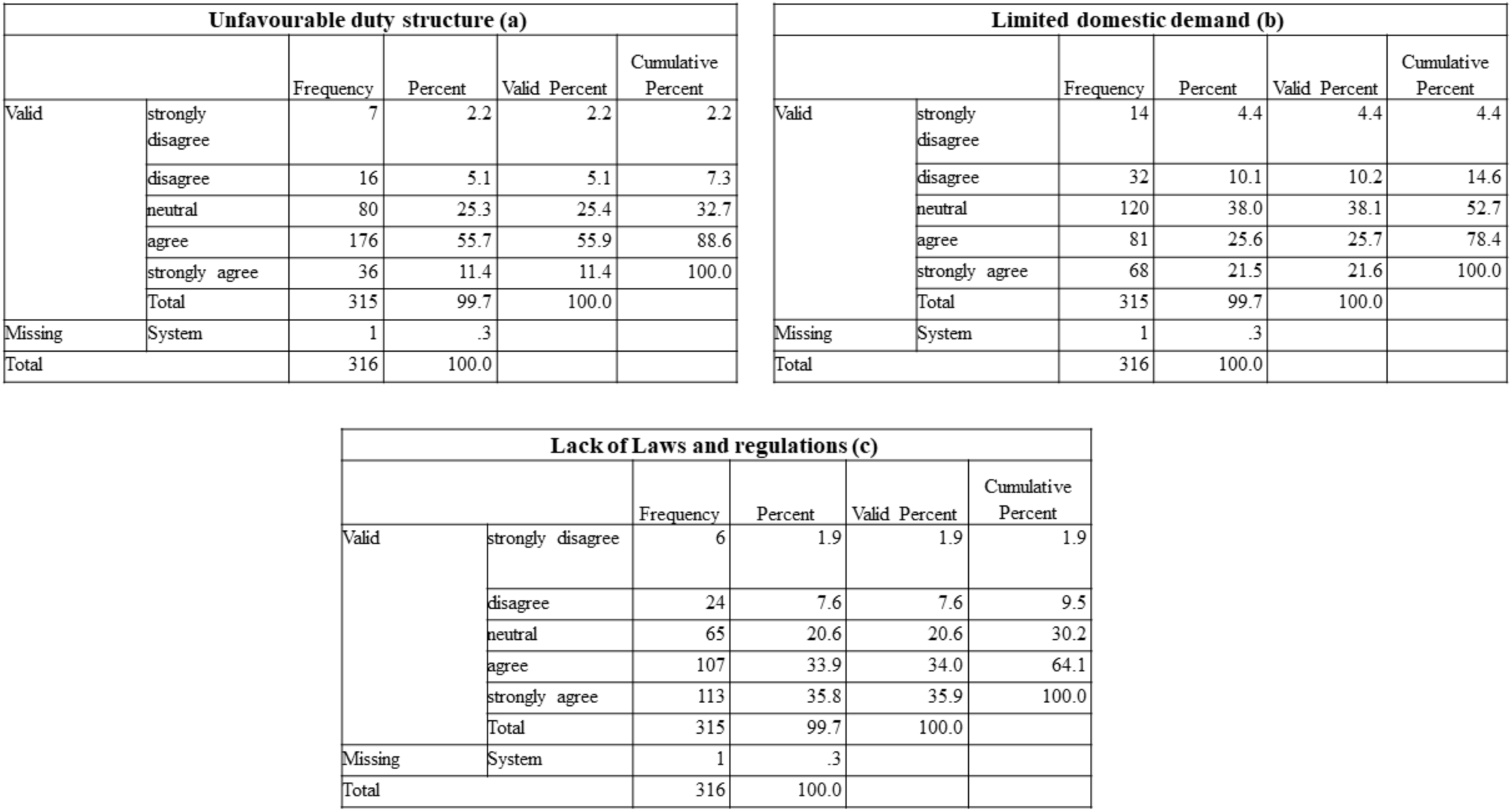
The frequency characteristics of a) Unfavourable duty structure b) Limited domestic demand c) Lack of laws and regulations estimated from the collected data from respondents.

**Fig 9.**
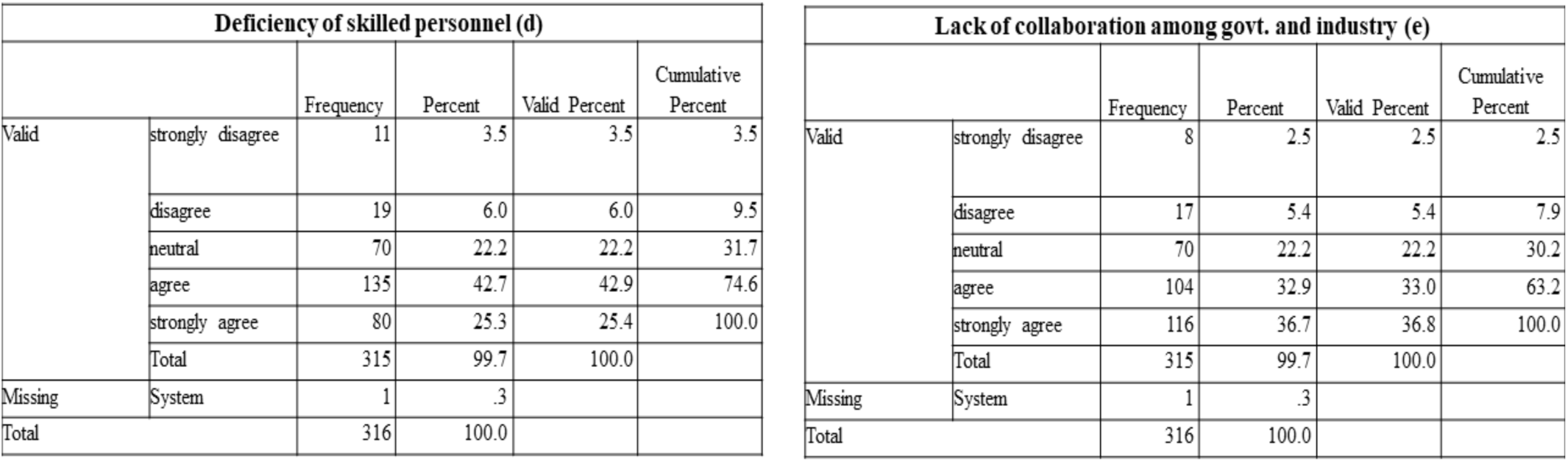
The frequency characteristics of d) Deficiency of skilled personnel e) Lack of collaboration among government and industry calculated from the respondents’ data.

From **Chart 11**, we can observe most people agreed or totally agreed to the possible reasons for the limitations of the biomedical device sector.

**Chart 11.**
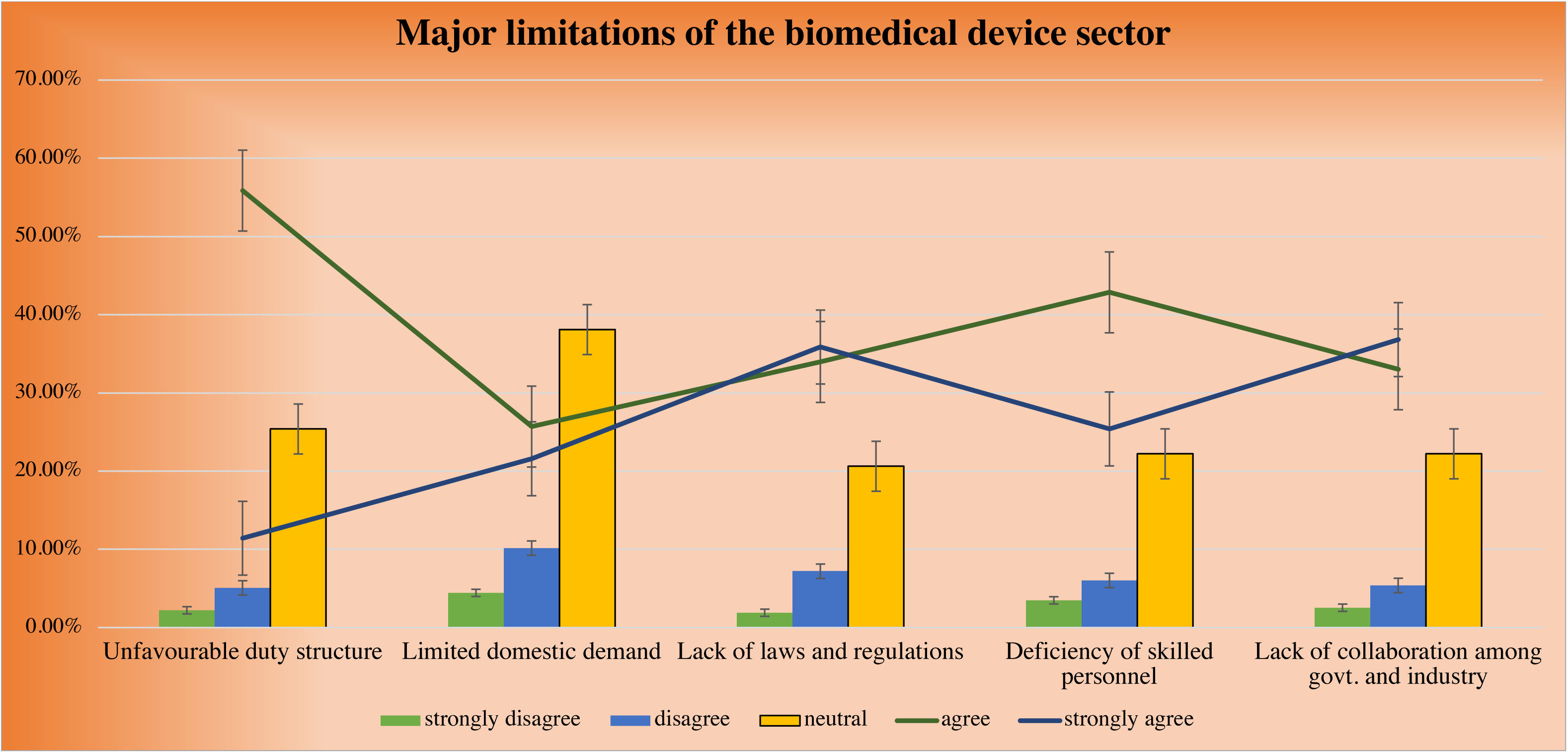
The percentage of responses recorded for each possible cause of limitations of the Indian medical devices industry.

Currently, imported raw substance consumers in an extent of manufacturing industries segment are in a place due to inverted customs duty structure that leads them non-competitive against inexpensive finished product imports and deters domestic value addition. Under the ‘inverted duty structure,’ import duty on accomplished products is lesser than on parts or elements used in producing the components [23]. And, 55.7% of respondents agreed to the unfavorable duty structure to be the primary cause for the limitations faced by the Indian medical devices industry.

To manufacture a safe, effective, economical device, the recruitment of skilled personnel is obligatory. In most cases, due to the deficiency of qualified personnel, the quality of finished products is below average compared to foreign products. 42.7% of respondents agreed with the same for the significant cause of hindrance of advancement in the Indian medical devices industry.

To diminish input costs of manufacturing goods, there is a necessity for the Government to offer infrastructure, logistic assistance that will empower manufacturers to diminish the cost of inputs and also provide economical products.

Through its headlight, the Govt. of India ‘Make in India’ inaugural counted heavily on the Indian manufacturers to encounter the uprising requirement of essential healthcare equipment for India, pressing the Indian medical devices industry to turn into self-reliant or Atmanirbhar Bharat. At this time, when India rushes to become Atmanirbhar, the medical device industry seeks Government endorsement in terms of various policy initiatives to establish an enabling ecological system for the native medical device industry to understand its capability. And, 36.7% of respondents strongly agreed that the government has not focused on the upliftment of the Indian biomedical sector.

The government should also bring some changes in their old policies, laws, and regulations to promote the manufacture of useful indigenous medical devices. Due to the lack of standard laws and regulations, medical devices of sub-standard quality are produced, decreasing their demand. 35.8% of respondents strongly agree on the same.

Due to the production of sub-standard products, the domestic demand also decreases, and the export of foreign products increases. Thus, to enhance the domestic demand, there is a need to build trust in Indians and outsiders that the products manufactured are safe, effective, and cheap. 38% of respondents majorly stayed neutral/ in confusion regarding the same as a cause for the limitations of the Indian medical device industry, and 25.6% of respondents agreed to it as the least cause for the same.

#### 3.1.7. Preventive approaches to overcome the limitations of Indian medical devices industry

As we had already discussed the causes of limitations of the Indian medical devices industry, we then tried to understand the perception of the respondents regarding the approaches/measures to be taken to overcome those limitations, as proposed by us. The question intends to find out how the limitations that are being faced by the industry can be improved.

To determine the reliability of data, we performed the Cronbach Alpha test, which resulted in a coefficient of alpha of 0.795, providing us the way for further data analysis (alpha value-0.6-1 is reliable).

**Table 9** depicts the values of central tendency estimated from the respondents’ data. The respondents have agreed to all the probable solutions to advance the Indian medical devices sector as interpreted from the mean values.

**Table 9.** The measures of central tendency.

| Statistics |  |  |  |  |  |  |
| --- | --- | --- | --- | --- | --- | --- |
|  |  | Reducing tax rate imposed on domestic manufacturers | Increasing skilled workforce | Implementation of strong laws and regulations to improve quality standards | By gaining trust of investors | Collaboration among government and industry |
| N | Valid | 315 | 315 | 315 | 315 | 315 |
|  | Missing | 1 | 1 | 1 | 1 | 1 |
| Mean |  | 3.7460 | 3.9238 | 4.0984 | 3.9524 | 4.1302 |
| Median |  | 4.0000 | 4.0000 | 4.0000 | 4.0000 | 4.0000 |
| Std. Deviation |  | .88094 | .93783 | .95429 | .92778 | .88811 |

**Fig 10** and **Fig 11** illustrate the frequency characteristics of all the possible approaches that can be taken to overcome the limitations of Indian medical devices industry.

**Fig 10.**
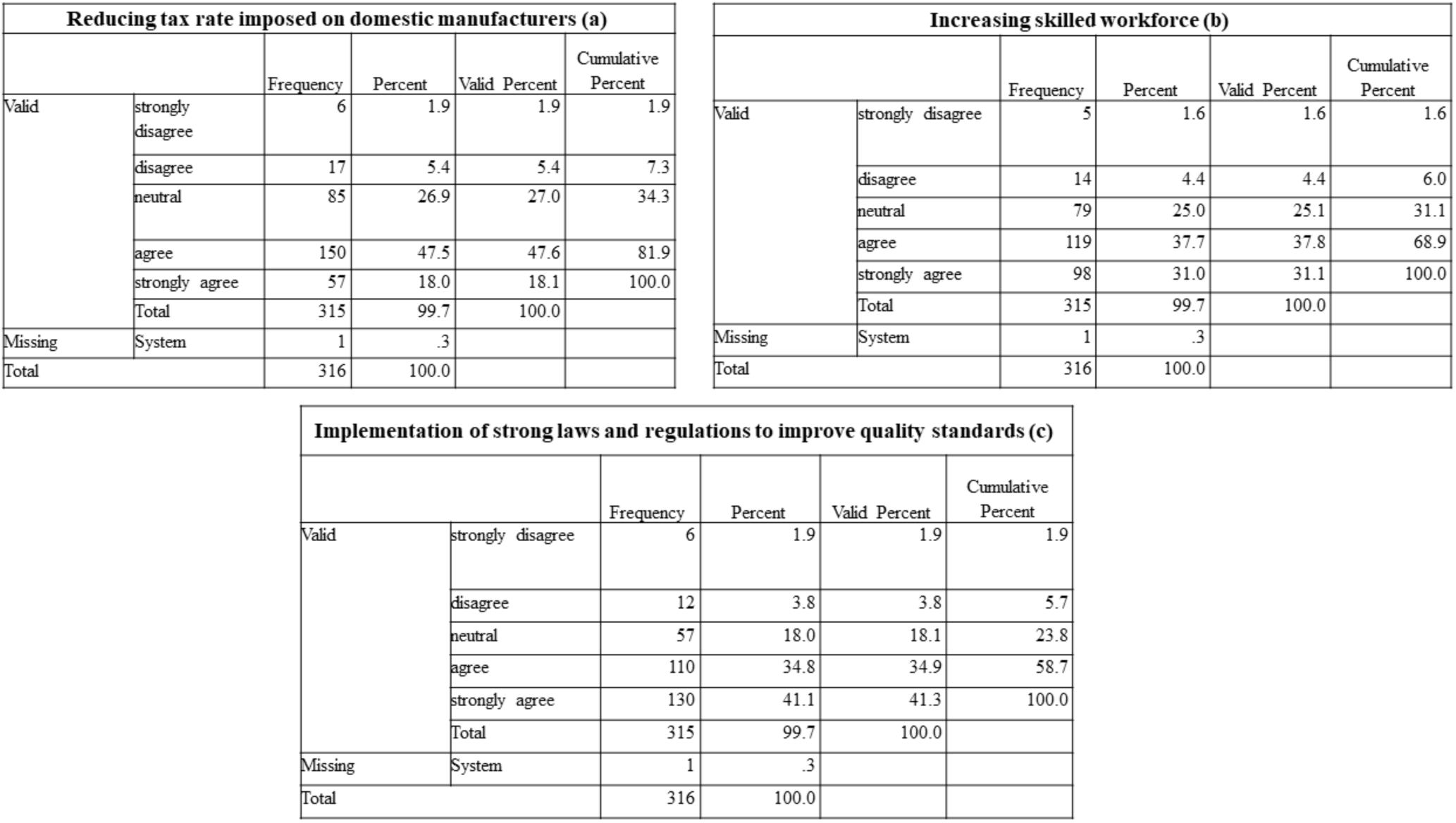
The frequency characteristics of a) Reducing tax rate imposed on domestic manufacturers b) Increasing skilled workforce c) Implementation of strong laws and regulations to improve quality standards estimated from the respondents’ data.

**Fig 11.**
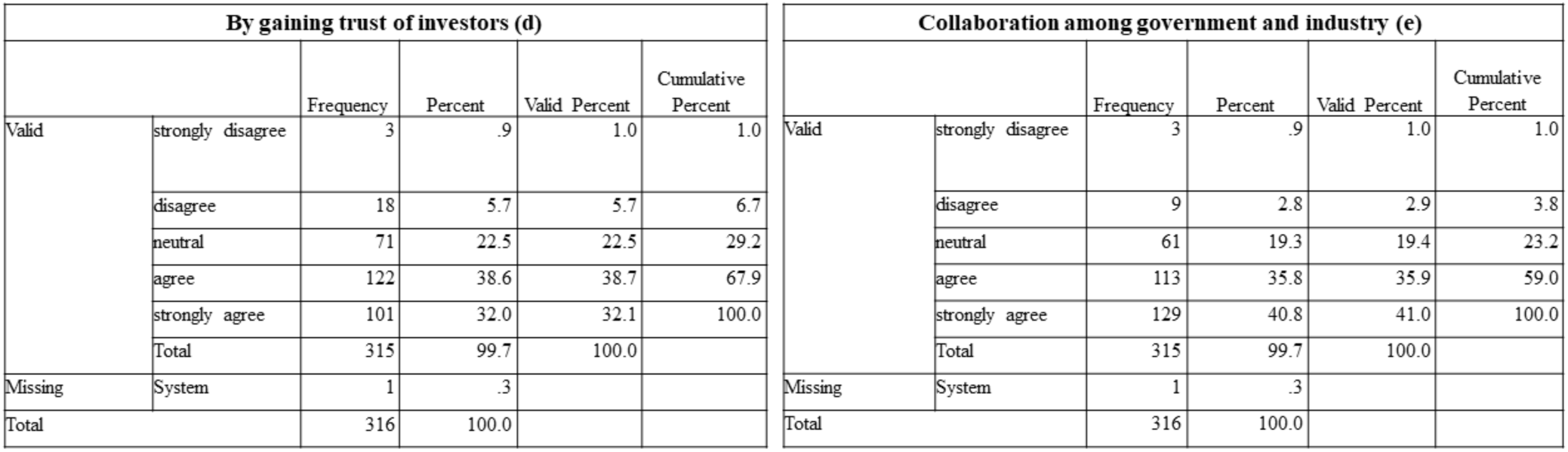
The frequency characteristics of d) By gaining trust of investors e) Collaboration among government and industry calculated from the respondents’ data.

As discussed earlier, inverted duty structures are majorly responsible for the cause of limitations of the Indian medical device sector [23]. And, to overcome the same limitation, there is a need to reduce the imposed tax rate on domestic manufacturers. 47.5% of respondents agreed with the same as the superior probable solution to advance the Indian medical devices sector.

41.1% of respondents have strongly agreed with implementing strong laws and regulations to improve the quality standard of the medical devices manufactured in India. And, 40.8% of respondents strongly agreed that the government should uplift the Indian medical devices industry for the betterment in manufacturing standard medical devices. A significant interference of the government in both matters can change the outlook of the Indian medical device sector.

37.7% of respondents agreed that the recruitment of skilled personnel is necessary to promote the manufacture of safe, standard medical devices, leading to the advancement of the Indian medical devices industry.

Investors also play a significant role in the betterment of the Indian medical devices industry. Due to the lack of quality products from the same, the investors hesitate to invest money to produce medical devices, as they fear going into a loss. Thus, gaining the trust of investors is essential to improve the medical devices industry. 38.6% of respondents resonated and agreed with the same to overcome the limitation of the Indian medical devices sector.

The results proved that, yes, there is a lot of scope for improvement in the Indian medical devices industry. The respondents also agreed on the possible ways which can be taken to improve the conditions.

**Chart 12** discusses the percentage of responses calculated for each possible approach to curb the limitations of the Indian medical devices industry.

**Chart 12.**
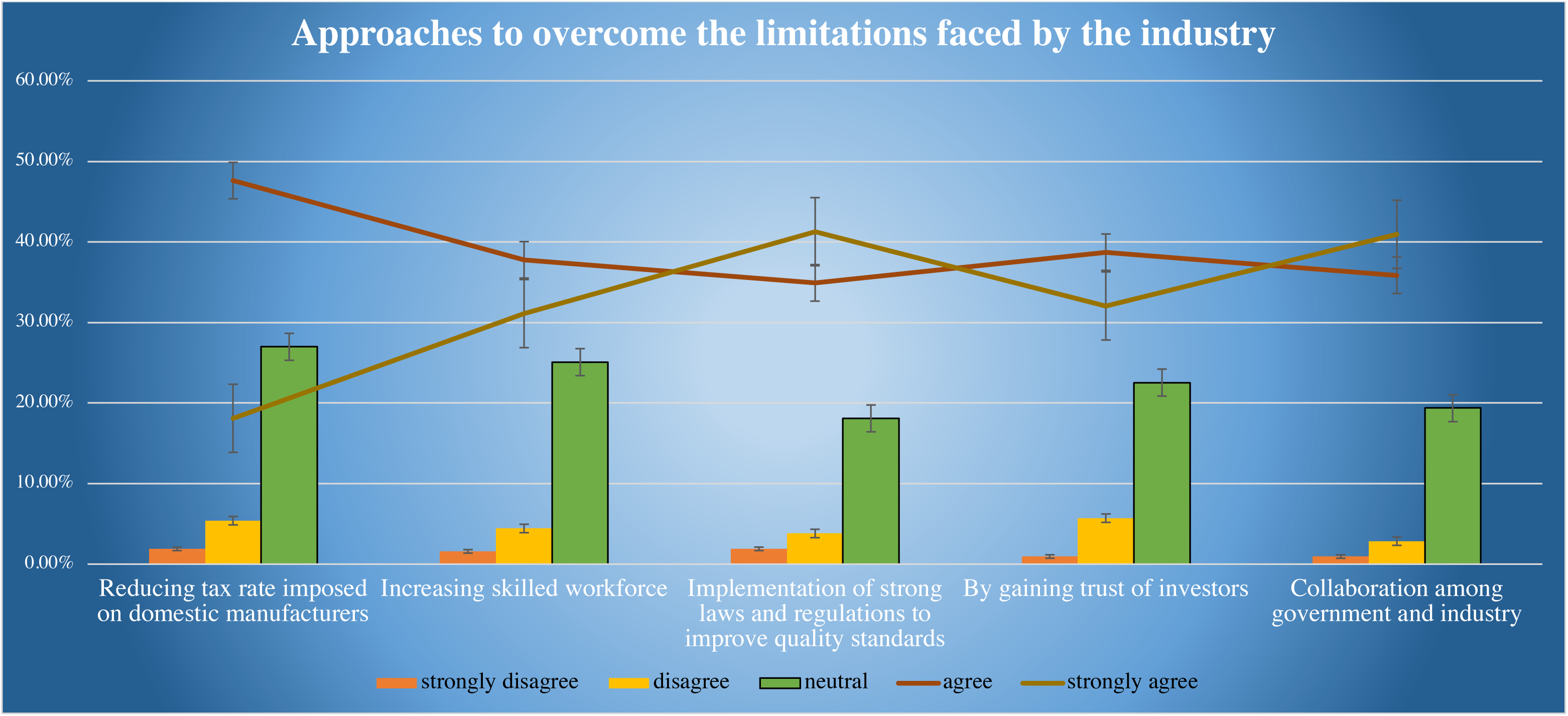
The percentage of responses calculated for each possible measure to overcome the limitations of Indian medical devices industry

#### 3.1.8. Is there any gap between Industry and Academia?

Industry-Academia gap! Yes, there is. It’s the gap between the engineering education system and the industry’s expectations from the entry-level students. And, something is lacking from both sides. We had a practical overview of the same debate, which made our motive to prepare the question to understand the knowledge and the perception of the respondents concerning the Industry-Academia gap.

According to **Chart 13**, 98.41% of respondents echoed a gap between industry and academia, causing a hold in the advancement of medical devices in India. **Table 10** demonstrates the frequency characteristics estimated from the respondents’ data.

**Chart 13.**
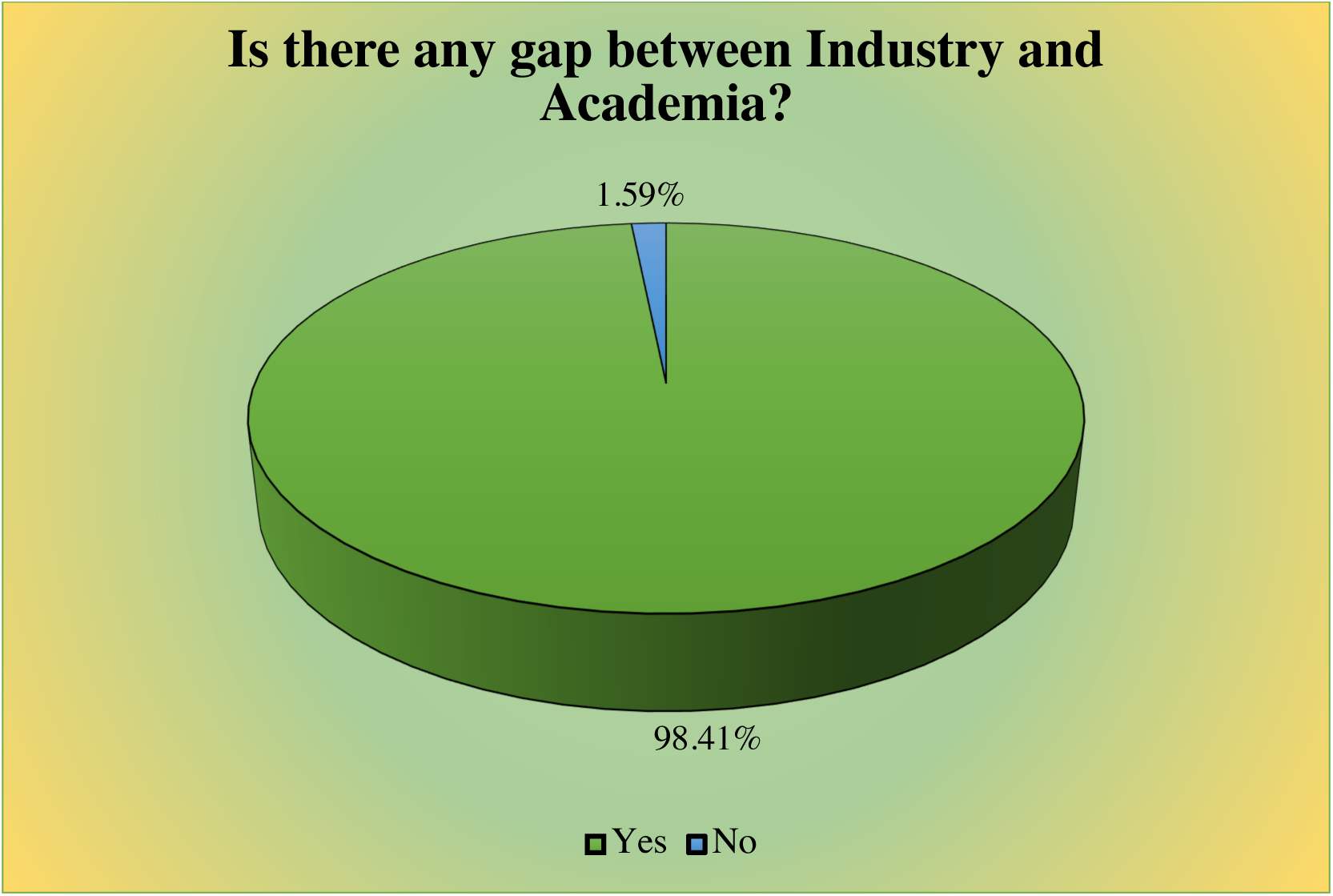
The percentage of respondents agreed and disagreed on the gap present between industry and academia.

**Table 10.** The frequency characteristics estimated from the data of the respondents.

| Is there any gap between Industry and Academia? |  |  |  |  |  |
| --- | --- | --- | --- | --- | --- |
|  |  | Frequency | Percent | Valid Percent | Cumulative Percent |
| Valid | Yes | 310 | 98.1 | 98.4 | 98.4 |
|  | No | 5 | 1.6 | 1.6 | 100.0 |
|  | Total | 315 | 99.7 | 100.0 |  |
| Missing | System | 1 | .3 |  |  |
| Total |  | 316 | 100.0 |  |  |

The results proved that there is a need to do some work to close the gap between industry and academia.

But the question still remains, how to bridge the gap between academia and industry. Thus, we proposed the best solutions for the same and tried to understand the perception of the respondents. The question intended to find out the possible solutions to improve the relationship between the industry and academia. **Table 11** describes the measures of central tendency of each possible solution for bridging the gap between academia and industry. The respondents have strongly agreed to ‘Academics should focus more on practical knowledge, in developing soft skills, rather than following the entire theoretical course.’ The respondents agreed to other solutions as interpreted from the mean values.

**Table 11.** The measures of central tendency.

| Statistics |  |  |  |  |  |  |
| --- | --- | --- | --- | --- | --- | --- |
|  |  | Industrial Visit/Internships | Diffusion of Knowledge through interaction of peers | By proposing idea of prototype to product to the industry | Collaborative funded start-up | Academics should focus more on practical knowledge, in developing soft skills rather than following the entire theoretical course |
| N | Valid | 313 | 314 | 315 | 313 | 312 |
|  | Missing | 3 | 2 | 1 | 3 | 4 |
| Mean |  | 4.0543 | 3.8599 | 4.1206 | 4.0607 | 4.2340 |
| Median |  | 4.0000 | 4.0000 | 4.0000 | 4.0000 | 4.0000 |
| Std. Deviation |  | .88084 | 1.00769 | .86587 | .94368 | .93506 |

To determine the reliability of data, we performed the Cronbach Alpha test, which resulted in a coefficient of alpha of 0.816, providing us the way for further data analysis (alpha value-0.6-1 is reliable).

The frequency characteristics of each possible solution to bridge the relationship between industry and academia are discussed in **Fig 12** and **Fig 13**.

**Fig 12.**
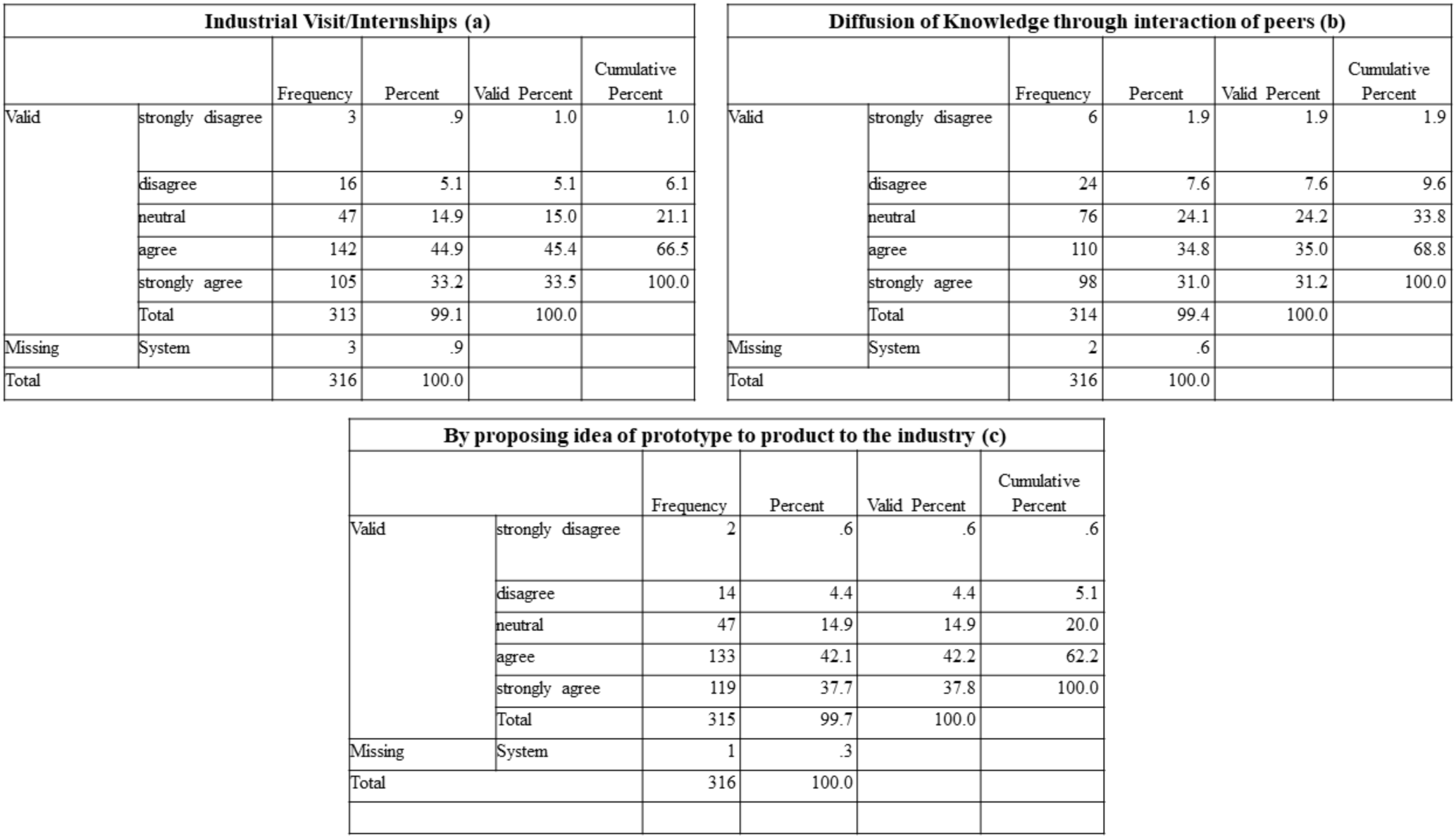
The frequency characteristics of (a) Industrial visit/internships b) Diffusion of knowledge through interaction of peers c) By proposing idea of prototype to product to the industry estimated from the respondents’ data.

**Fig 13.**
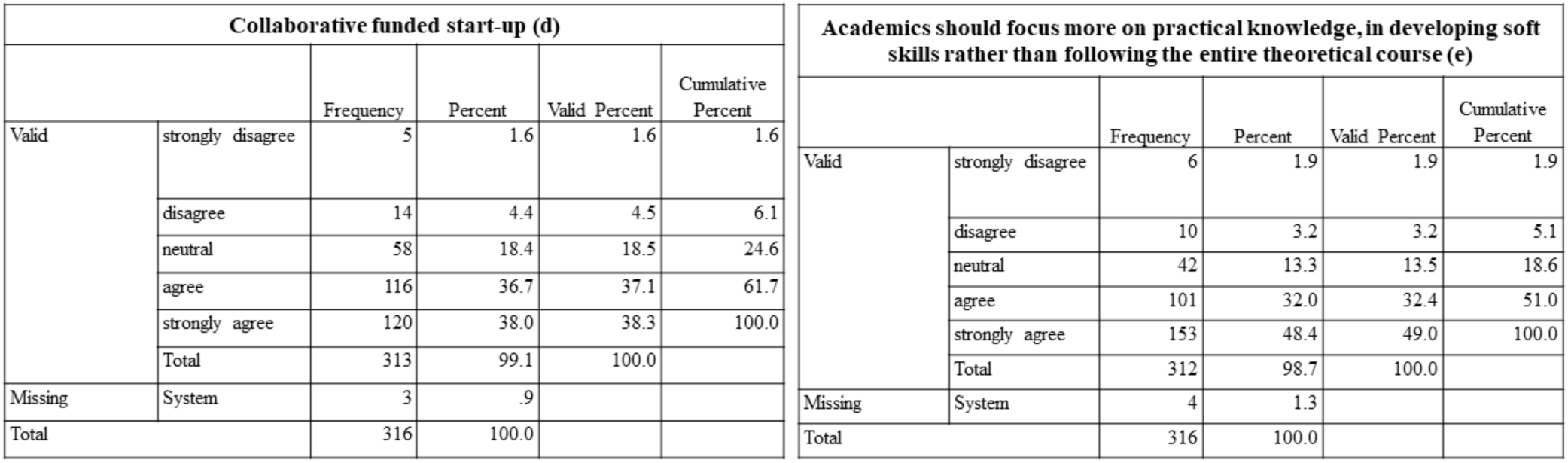
The frequency characteristics of d) Collaborative funded start-up e) Academics should focus more on practical knowledge, in developing soft skill rather than following the entire theoretical course calculated from the data acquired from the respondents.

**Chart 14** describes the percentage of responses for each probable solution to better the relationship between industry and academia.

**Chart 14.**
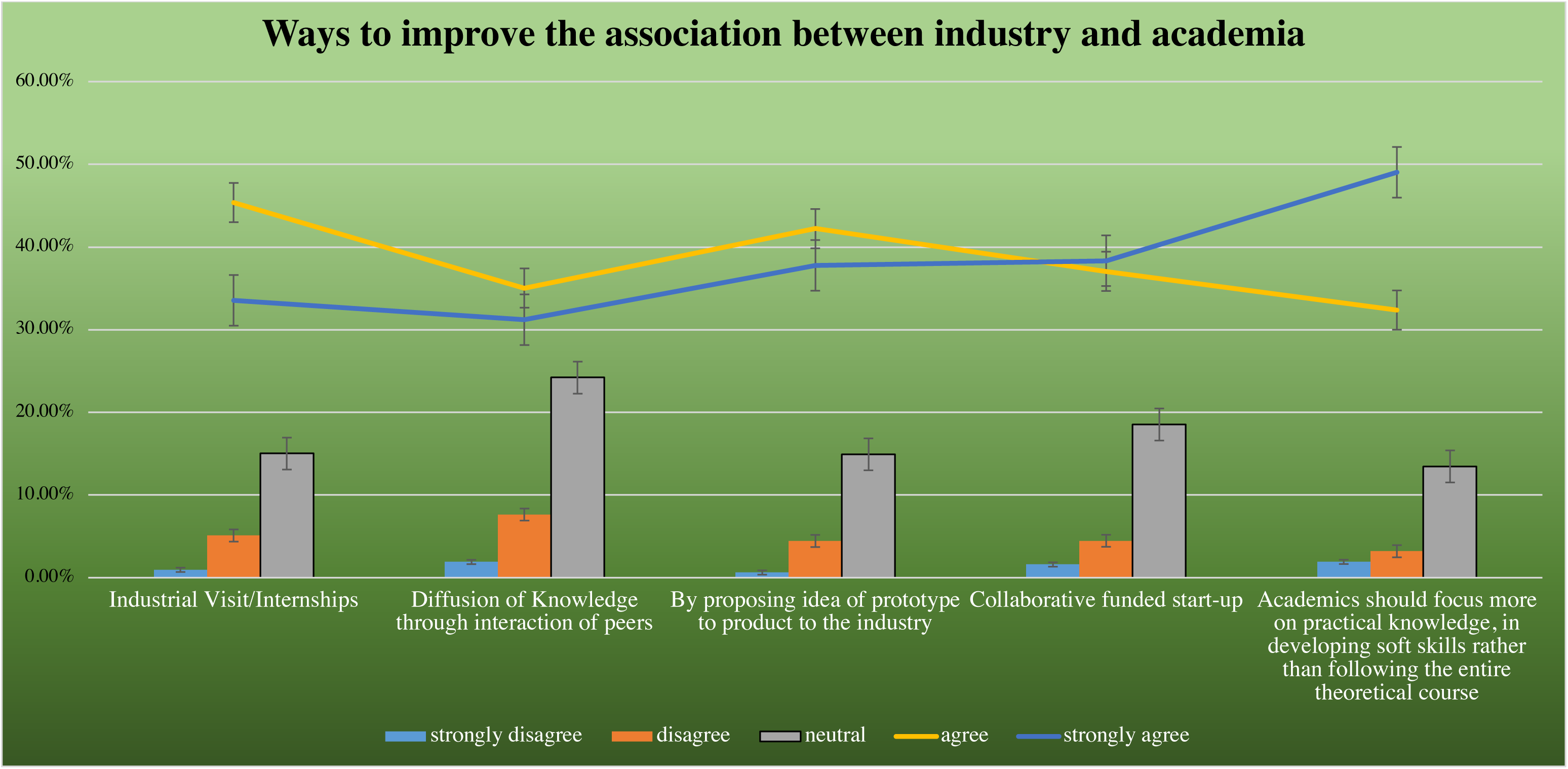
The percentage of responses calculated for each possible solution to bridge the gap between academia and industry

Be it any proficiency, encouraging students play a very significant role. Every student’s uniqueness and strength lie in diverse areas. The pharma and biomedical students should be provided with opportunities to explore/discover their potential. Out-of-the-box work by the students could offer them strong enthusiasm. On the other hand, the budding researchers should not be demotivated in any cases, leading to the destruction of creativity and moral confidence. And, the respondents also resonated with the same vision of performing out-of-the-box work like preparing prototypes. 42.1% and 37.7% of respondents have agreed and strongly agreed with pitching the prototype’s ideas to the industry, respectively.

The syllabus of the pharma/biomedical engineering colleges is now a highly debated topic. According to various dignified persons, the syllabus should be discarded and prepared on the latest trending topics. Thus, a balanced syllabus should be provided, for example, a section for fundamental courses and another section for advanced courses, which would build their knowledge on the trending topics. The students should also be provided with hands-on exercises, assignments, projects for the preparation of devices. This would make the foundation base for the students, and they would be easily recruited by the industry. 48.4% of respondents have strongly agreed to the same.

College students are generally not conscious of real workplace expectations. Hence cognizance is to be created by revealing the real-time workplace to them. Internships/Industrial visits can undoubtedly connect the gap between academia and industry. Through this, the students not only discover in terms of their employment outlook but also find out about other behavioral elements. These behavioral features encompass oral communication with colleagues, seniors, and boss, learning to adapt and complete the given work, providing quality yield and learning on relevant duties. Attending common activities like team-meeting, offsite-workshops can construct a big visualization of how a company works. 44.9% of respondents have agreed to the same.

The diffusion of knowledge is a significant way to improve the relationship between academia and industry. The highly qualified academicians and industrialists should be invited for a guest talk or a lecture in the universities to promote and diffuse advanced knowledge regarding the preparation, regulation of medical devices. Their main intention should lie in building a healthy relationship between academia and industry to advance the Indian medical devices industry. 34.8% of respondents agreed to our proposed solution.

Another significant solution to bridge the gap between Industry-Academia is the collaboration system. Either the industry or the government can offer the fundings to scale up the proposed prototype prepared by the academicians. As the universities majorly lack funds, their innovations do not get converted into products. Thus, the collaboration will play a major factor in scaling up the lab-scale product. 38% of respondents agreed to our proposed solution.

### 3.2. Indian Ventilators

#### 3.2.1. An insight into the perception of Indians for Ventilator manufacturing Indian companies

Every person knows that they are put on mechanical ventilators whenever a patient goes under difficult breathing conditions. But, we suspected that there might be significantly low awareness among the Indian people about the Indian companies that manufacture ventilators.

The main motive was to determine the percentage of awareness about the Indian companies who manufacture ventilators from the respondents. Some options like a company providing reagents or instrumentation to the biomedical industry and belonging to the bio-informatics category and the name of the hospital were also included to check the understanding.

The frequency characteristics calculated from the respondents’ data for each standard medical devices company are discussed in **Fig 14**.

**Fig 14.**
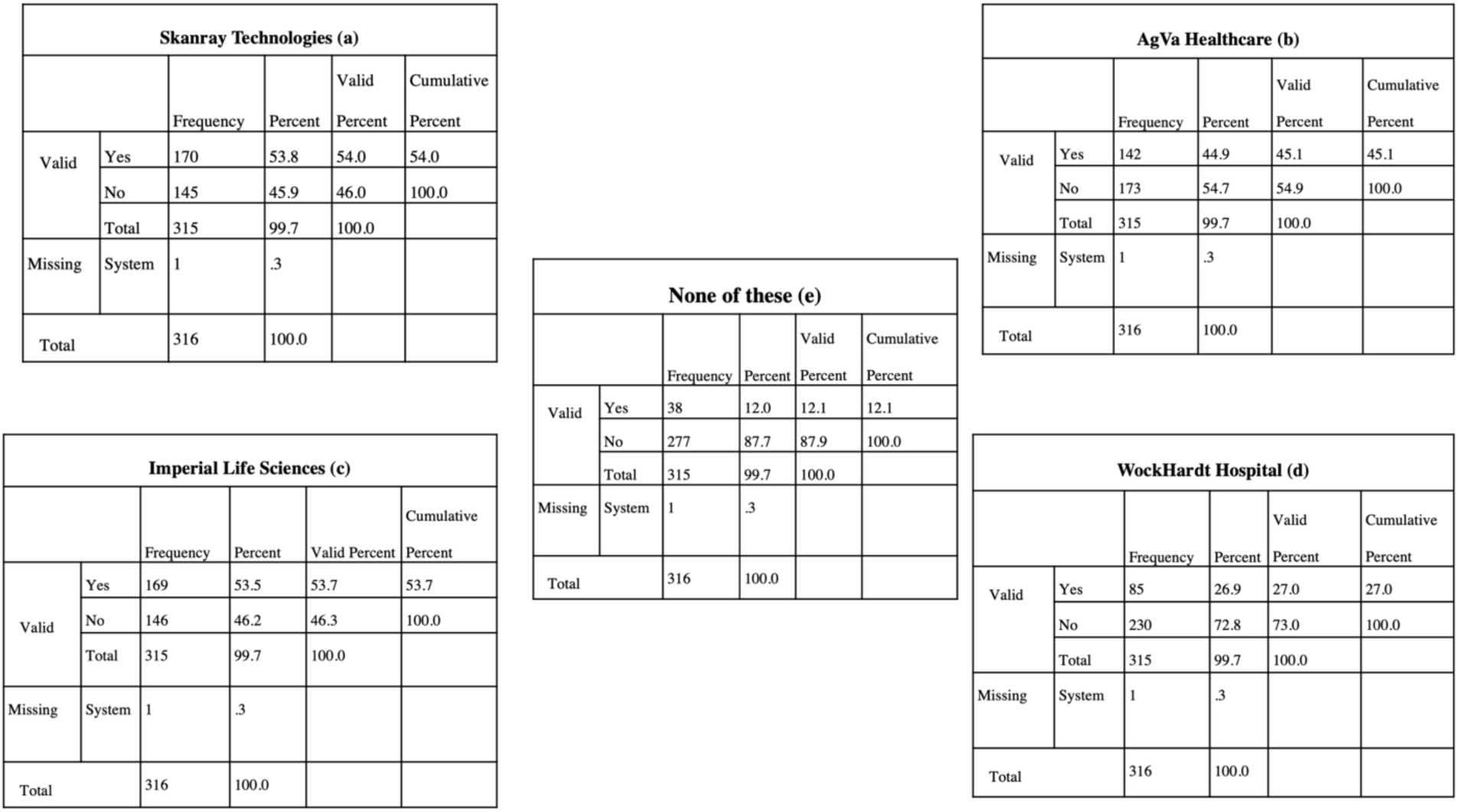
The frequency tables of (a) Skanray Technologies (b) AgVa Healthcare (c) Imperial Life Science (d) WockHard Hospital (e) None of these acquired through SPSS software according to the responses of the targeted community.

Observing **Chart 15**, we conclude that, on average, 53.87% of respondents accurately identified Indian ventilator manufacturing companies (Skanray and AgVa Healthcare). About 53.54% of respondents incorrectly determined bioinformatics and reagent supplier company, Imperial Life Sciences as a ventilator manufacturing company. For the other option, which is a hospital, Wockhardt, people were more aware, and 73% of respondents identified it as a non-manufacturing ventilator company. At the same time, approximately 12% of respondents found the options doubtful and haven’t voted for any.

**Chart 15.**
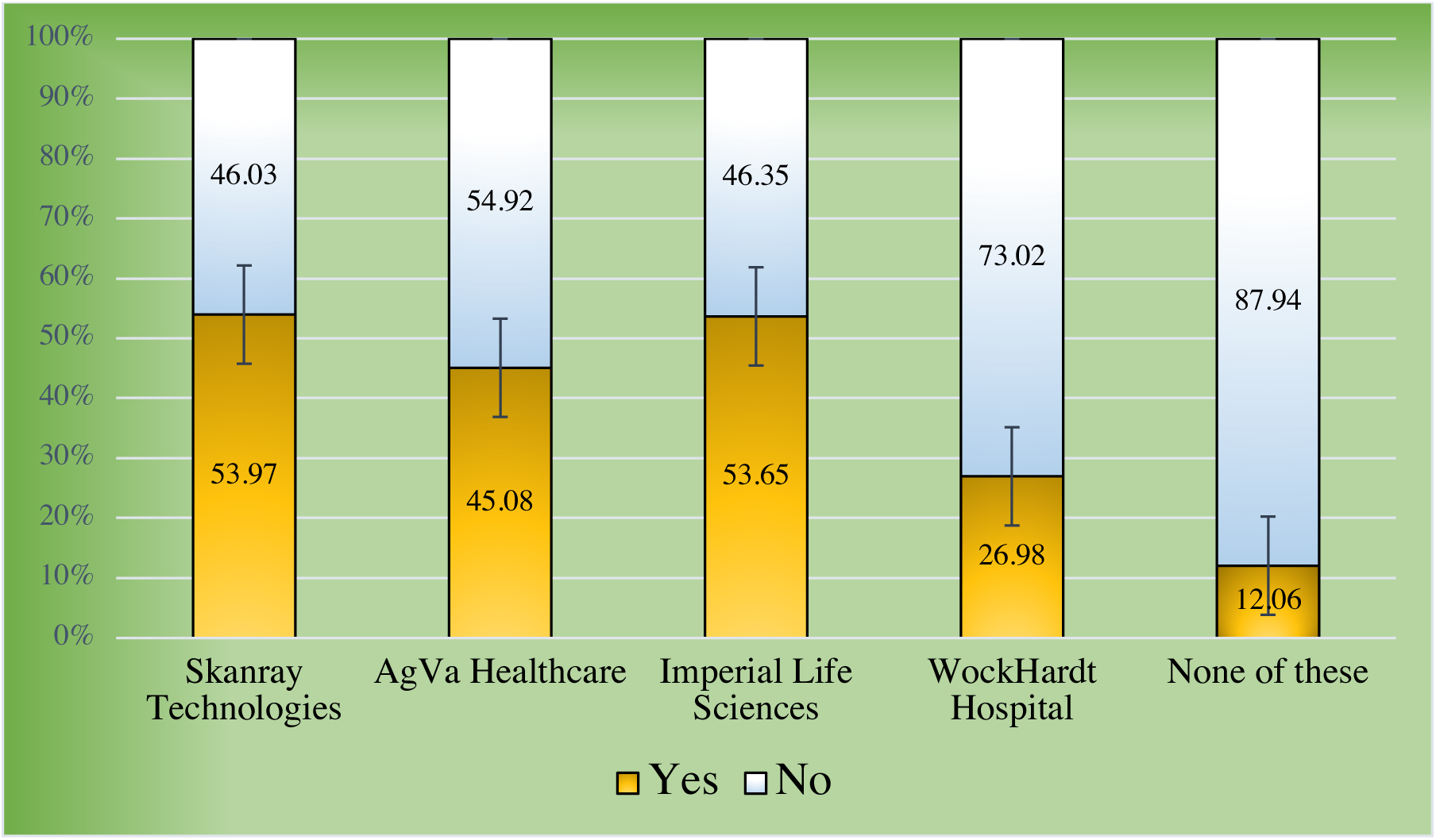
The percentage of responses each company received from the respondents regarding their knowledge on Indian ventilator manufacturing companies.

From our inspection, we actually determined right about the less awareness about the same. The overall result of the survey showed around half of the respondents correctly identified ventilator manufacturing companies. Still, a majority of respondents have mistaken a bioinformatics company for a ventilator manufacturing company. Respondents are aware that the hospital doesn’t serve as a ventilator manufacturing unit, which is good. While there is only a minor percentage of people, who didn’t find any options right and haven’t selected any.

#### 3.2.2. Understanding the general awareness of Indians about the production of faulty ventilators due to mass production during the SARS-CoV-2 pandemic

The SARS-CoV-2 pandemic has changed the entire health scenario of the world [24]. The major device that came into the limelight is ventilators, required to provide oxygen to the COVID-19 affected patients. As the number of patients is increasing at an unparalleled pace, there seems to be a scarcity of providing ventilators to each patient. India is completely dependant on foreign countries regarding the ventilators. Under these circumstances, the Indian companies started mass production of ventilators, which came into the market without proper regulatory inspection and safety standards. Thus, it’s a major issue that should be known to the fellow Indians. For instance, as per the news on June 30, 2020, St. George hospital and JJ hospital in Mumbai have returned about 81 ventilators, which were manufactured by the AgVa Healthcare-Maruti consortium because they failed to increase oxygen to the necessary levels for critical COVID-19 patients. Among these, 42 ventilators were donated to J J hospital by the American India foundation while 39 ventilators to St. George hospital by the private charity organization Rotary Club (Both are NGOs). In a letter dated 19^th^ June, doctors highlighted that AgVa ventilators were not supplying 100% oxygen to COVID-19 patients and that oxygen didn’t increase to the desired level [25]. Two employees of AgVa healthcare revealed that the company had manipulated to show that devices were pumping more oxygen into patients’ lungs than they were [26]. A Rajkot-based private company, Jyoti CNC, has developed a ventilator named Dhaman in just 10 days, having a manufacturing cost of less than 1lakh. The ventilators were installed just after testing on one patient, and also, the installed ventilators didn’t have a DCGI license. So far, 900 ventilators were installed in the state, while 230 of them were present in Ahmedabad’s Civil Hospital [27]. A similar case was reported in the case of Andhra Pradesh MedTech Zone(AMTZ). Jyoti CNC automation and AMTZ have received about Rs. 22.5 crore payment from PM cares funds, but failed to produce good quality ventilators and later dropped from the ministry list. The question was intended to check the knowledge of respondents on the inefficient working of ventilator cases that emerged during the pandemic [28].

**Fig 15** discusses the measures of central tendency, frequency statistics, and percentage of responses recorded from the data of the respondents.

**Fig 15.**
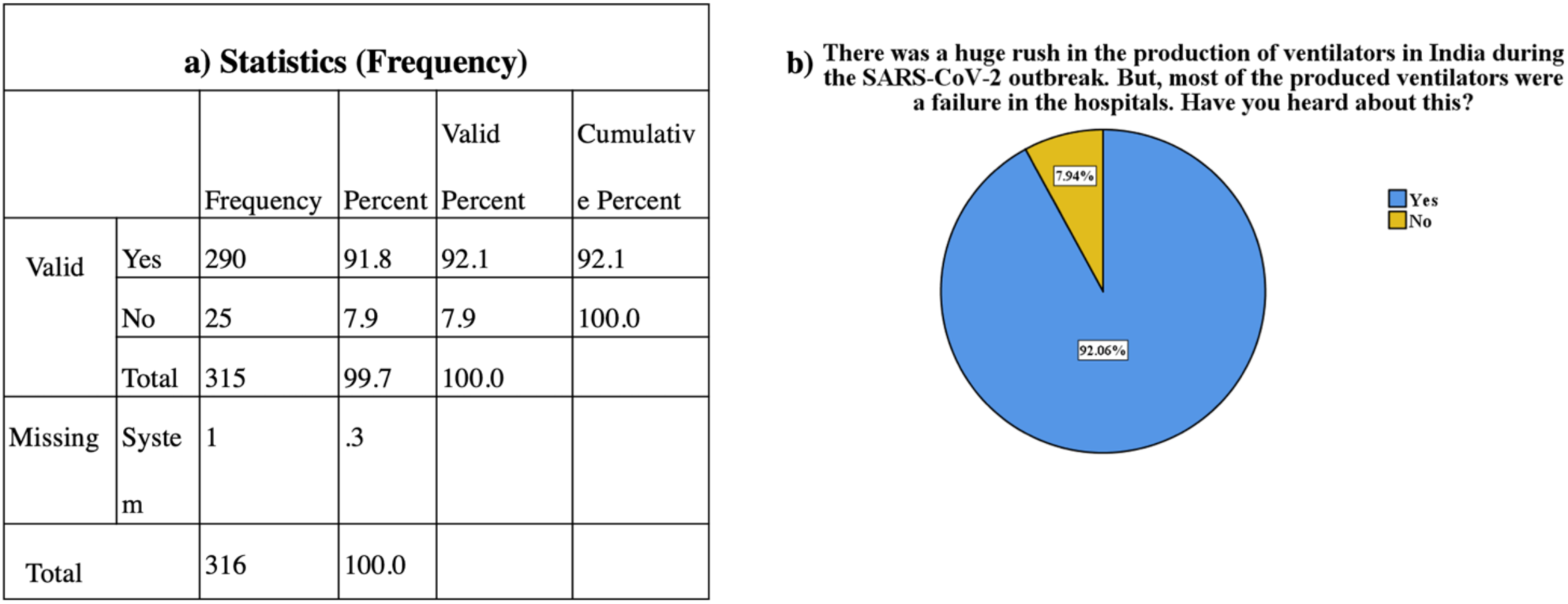
The a) Frequency statistics b) Percentage of responses represented via Pie-chart for the question of awareness on the production of faulty ventilators in India.

It was observed that 92.6% of Indians are aware of the fact that current failure cases of ventilators were reported in India. Only about 7% of people are unaware of this situation, which was not expected. It was expected that people would be significantly aware of the current scenario of the ventilators in India. Respondents are mostly aware of the ineffective working of newly produced ventilators, which correlates with our hypothesis for the particular question. The question doesn’t confirm that the people have exact knowledge about the problem, but it surely confirms they might have an idea about this condition.

#### 3.2.3. Improvement and adaptation of policies to manufacture ventilators easily in India

The SARS-CoV-2 outbreak has pointed out several loopholes in the healthcare and biomedical industry. One of them is the lack of proper regulations and policies for manufacturing safe, economical, and effective biomedical devices like ventilators. The question was intended to correlate our possible solutions for making the manufacturing process of ventilators as smooth as possible. In the COVID-19 era, many countries have used various strategies for making the ventilator manufacture process as easy as possible. A few of them were suggested in this question as follows.

Compulsory Licensing (CL) policy is an intervention mechanism that allows the government to achieve two different objectives at once; one is rewarding patentees for their invention while the other is making the patented products available to the third party in an emergency crisis like COVID-19. This policy is adopted by some countries like Canada for increasing the availability of the patented products and giving other manufacturing companies rights to construct, use, and sell the patented invention whenever necessary [29].

For the COVID-19 outbreak, USFDA took one initiative known as ‘Open sourcing and ventilator software and design.’ The scheme made ventilator designs and software publicly available and allowed other companies to do the modifications up to certain limits for increasing the functionality of ventilators [30]. Initially, there was no specific standard available for ventilators, but after the COVID-19 outbreak, the government issued separate guidelines for it and made consistent changes as required. Because of the constant varying Indian standards, local companies face difficulties while designing parts of the ventilators and getting approval for the final product.

In this crisis, many industry personnel doesn’t have much knowledge about developing new ventilators or don’t have an understanding of the whole manufacturing process. So, the addition of a particular course regarding this topic may play an important role in increasing literacy about the ventilator industry.

To determine the reliability of data, we performed the Cronbach Alpha test, which resulted in a coefficient of alpha of 0.817, providing us the way for further data analysis (alpha value-0.6-1 is reliable).

**Table 12** demonstrates the values of central tendency calculated from the data of the respondents. And, all the respondents agreed to all the solutions as interpreted from mean values.

**Table 12.** The measures of central tendency.

| Statistics |  |  |  |  |  |
| --- | --- | --- | --- | --- | --- |
|  |  | Compulsory Licensing<br>(authorizations<br>permitting a third party<br>to make, use, or sell a<br>patented invention<br>without the patent<br>owner's consent) | Open-source knowledge<br>hub for designing and<br>working of ventilators | Improve regulatory<br>standards | Inclusion of a particular<br>course for all the<br>industry personnel |
| N | Valid | 315 | 315 | 315 | 315 |
|  | Missing | 1 | 1 | 1 | 1 |
| Mean |  | 3.8889 | 3.8286 | 4.2794 | 4.1048 |
| Median |  | 4.0000 | 4.0000 | 4.0000 | 4.0000 |
| Std. Deviation |  | .96617 | 1.00752 | .89836 | .92994 |

The frequency characteristics of each of the policies adopted to manufacture ventilators in India with ease are discussed in **Fig 16** and **Fig 17**.

**Fig 16.**
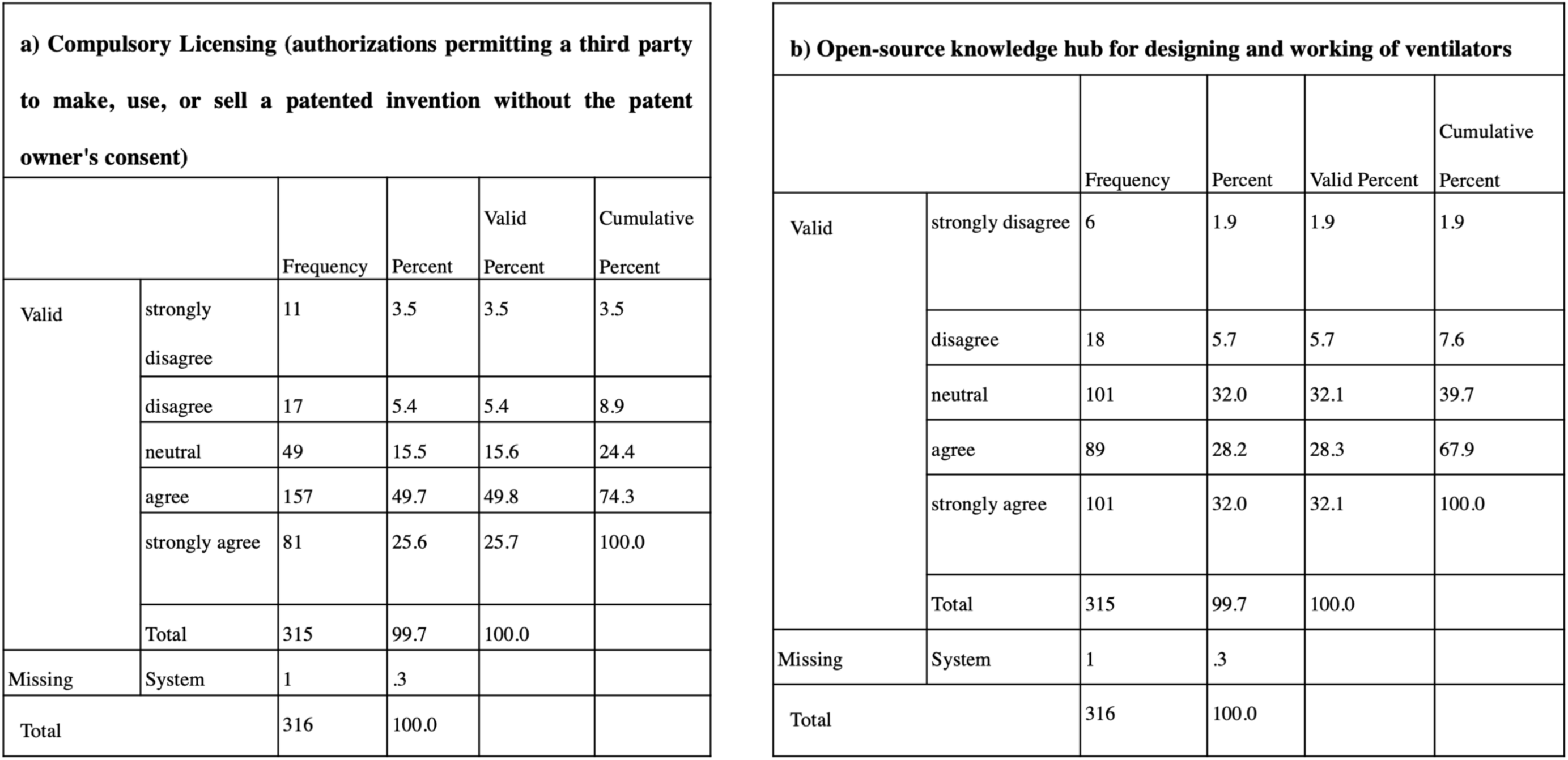
The frequency table of (a) Compulsory licensing and (b) Open-source knowledge hub for designing and working of ventilators calculated according to the perception of respondents.

**Fig 17.**
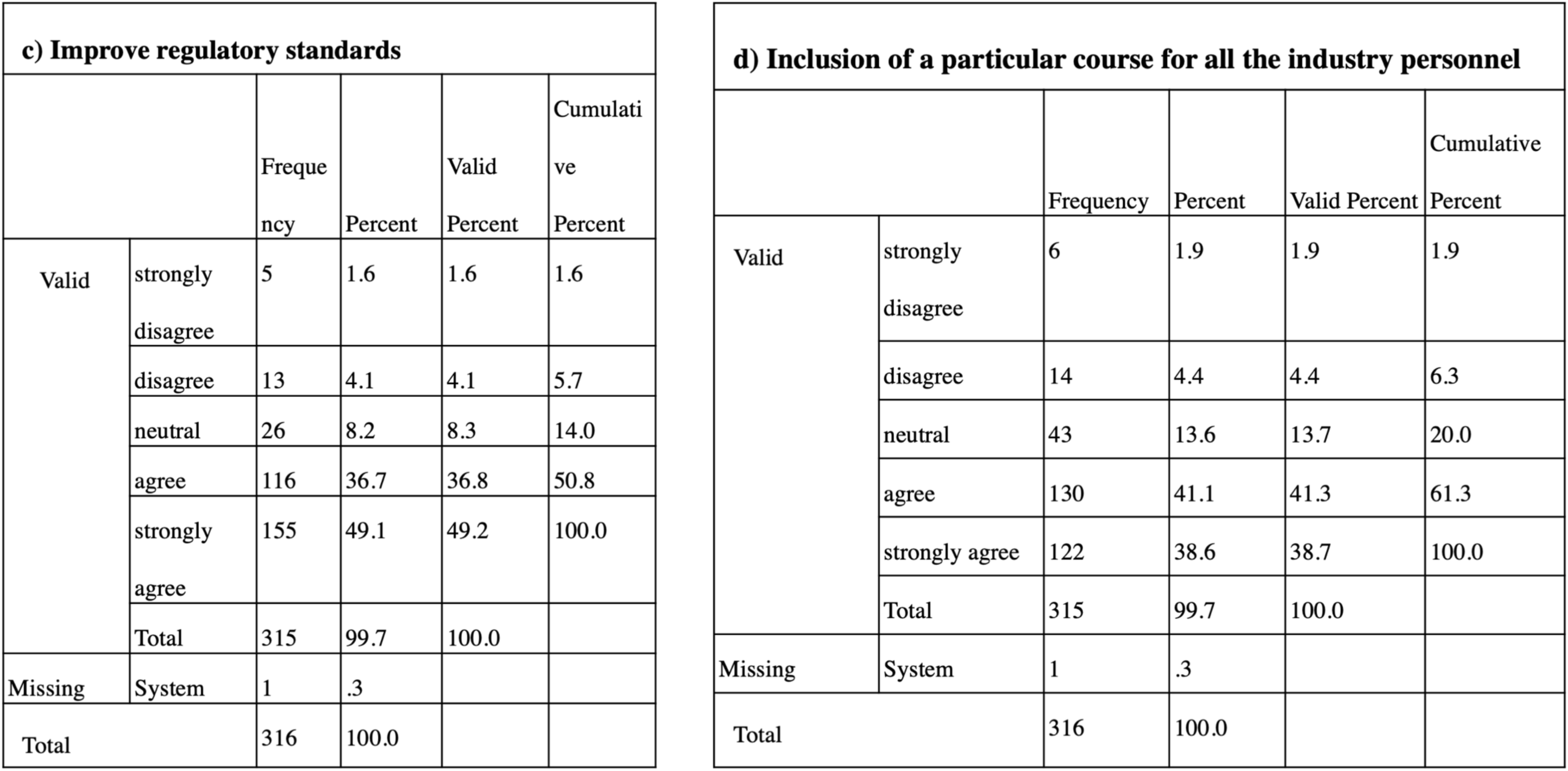
The frequency table of (c) Improve regulatory standards and (d) Inclusion of particular course for all the industry personnel calculated according to the perception of respondents.

In the survey, it is observed (**Chart 16**) that about 49% of respondents strongly agreed on ‘improvement in the regulatory standards,’ which could serve as a better option for making manufacturing of ventilator easier. Thus, India should adopt the Compulsory Licensing scheme to increase the availability of patented ventilator-related products. Also, improvement in the standards of the ventilator is necessary for boosting the production process. On average, 49.76 % of respondents agreed that ‘compulsory licensing policy’ and ‘inclusion of a particular course for Industry personnel’ may serve as good measures for the same reason. On the other hand, about 32% stayed neutral about the addition of an ‘Open-source knowledge hub for designing and working of ventilators.’ In India, this system may serve as a boon for companies in underdeveloped areas as they can access qualitative ventilator designs from the database and make them on their own.

**Chart 16.**
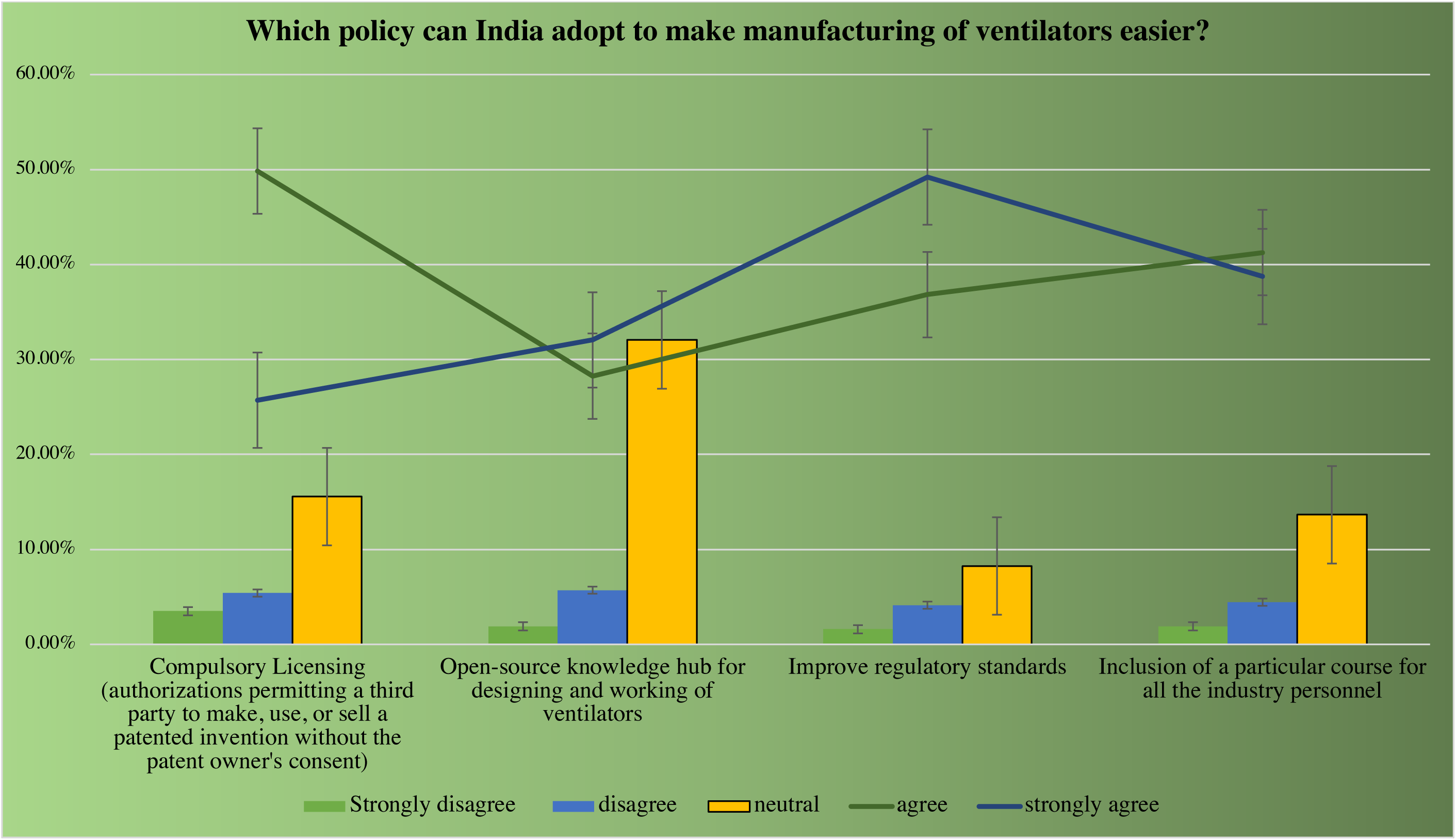
The percentages of responses recorded for every possible solution for improving the manufacturing of ventilators in India.

The survey’s overall result conveyed that the respondents strongly recommend improving regulatory standards as the most suitable solution. Respondents also agreed on Compulsory licensing and the Inclusion of a particular course for industry personnel, which could also serve as solutions secondary to the previously mentioned. Respondents prefer to stay neutral about the option of adding an Open-source knowledge hub for designing and working of ventilators, which was unexpected. This might be because respondents may have thought about the possibility of spreading misinformation by using such an open database or developing inefficient working ventilator devices, which in the end, slows down the manufacturing process.

#### 3.2.4. Probable reasons for less production of ventilators in India

Despairing hospitals state that they can’t find anywhere to purchase the ventilators, which assist patients in breathing and those at death’s door due to COVID-19. The number of imported ventilators and manufactured ventilators didn’t cope up as the COVID-19 cases started mounting. India, which was primarily dependent on foreign ventilators, was trapped in a hole. The export system got closed, and very few Indian medical companies manufacture ventilators at an economical and complying safety standard. The background of our question stands on the same. The motive behind our question was intended to investigate the reason behind the less production of ventilators in India.

To check the reliability of the data, we performed a Cronbach alpha test that gave a coefficient of alpha .800 (alpha value-.6-1 reliable), flooring our way for further analysis of the collected survey data. **Table 13** depicts the values of central tendency estimated from the respondents’ data. And, all the respondents agreed to all the reasons for less production of ventilators as interpreted from mean values. **Fig 18** and **Fig 19** discuss the frequency characteristics of each of the possible reasons for the less production of ventilators in India calculated from the respondents’ data.

**Fig 18.**
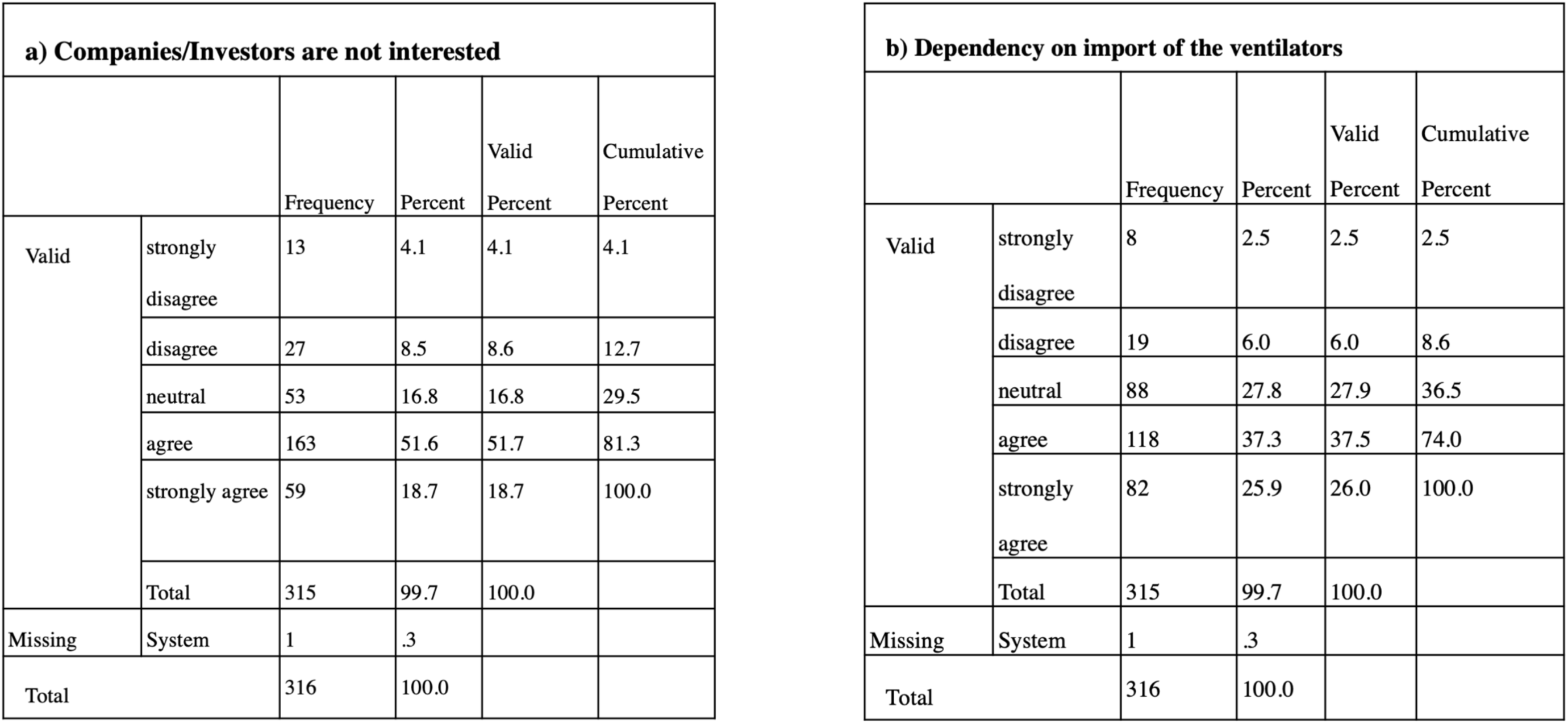
The frequency statistics of (a) Companies/investors are not interested (b) Dependency on import of the ventilators acquired from the responses collected through the survey.

**Fig 19.**
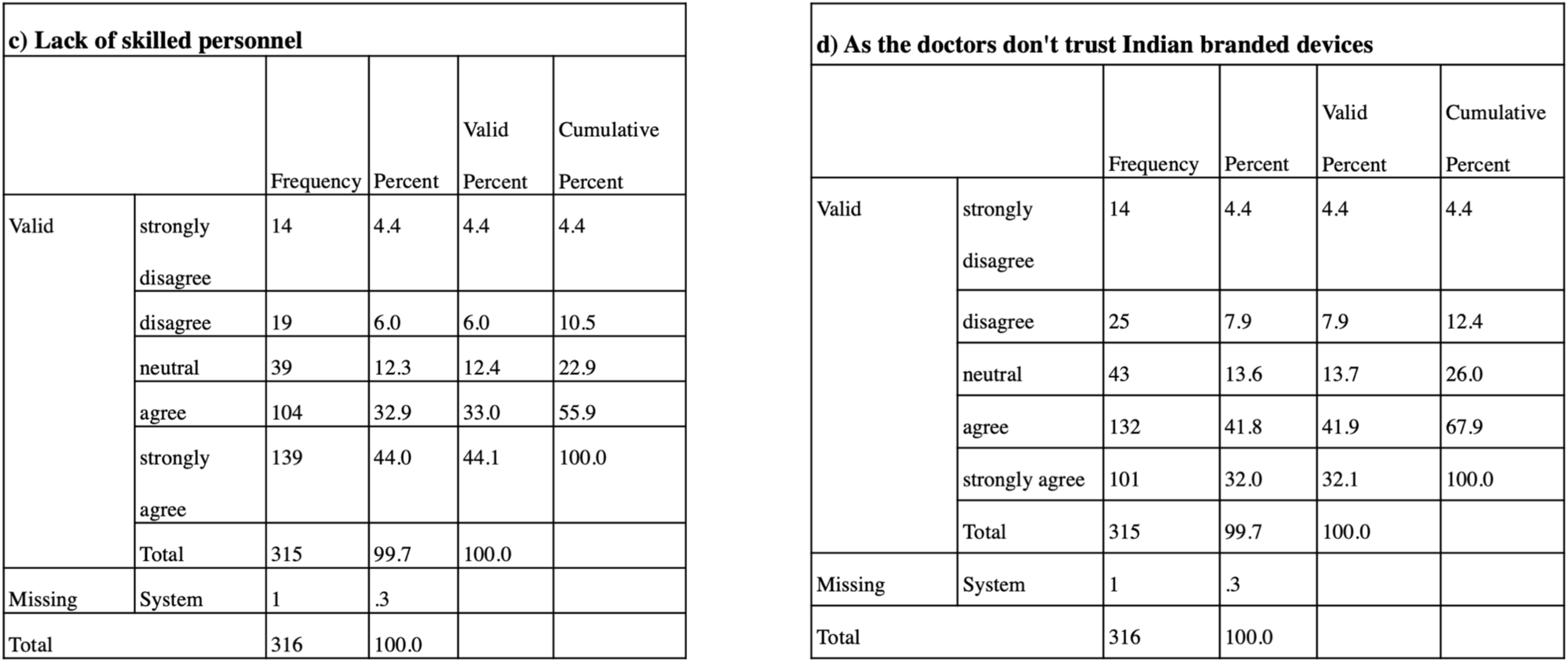
The frequency statistics of (c) Lack of skilled personnel (d) As the doctors don’t trust Indian branded devices acquired from the survey responses.

**Table 13.** The measures of central tendency.

| Statistics |  |  |  |  |  |
| --- | --- | --- | --- | --- | --- |
|  |  | Companies/Inve<br>stors are not<br>interested | Dependency on<br>import of the<br>ventilators | Lack of skilled<br>personnel | As the doctors<br>don't trust Indian<br>branded devices |
| N | Valid | 315 | 315 | 315 | 315 |
|  | Missing | 1 | 1 | 1 | 1 |
| Mean |  | 3.7238 | 3.7841 | 4.0635 | 3.8921 |
| Median |  | 4.0000 | 4.0000 | 4.0000 | 4.0000 |
| Std. Deviation |  | .99836 | .98285 | 1.09825 | 1.08012 |

In the survey, it was observed (**Chart 17**) that about 44% of respondents strongly agreed with the opinion that the lack of skilled personnel might be the primary reason behind less production of ventilators. Approximately 51% of respondents agreed with ‘Companies/Investors are not interested,’ which may be the primary reason, while about 41% of respondents said that the secondary reason is ‘As the doctors don’t trust Indian branded devices.’ About 37 % of respondents agreed with dependency on importing the ventilators as a reason for the mentioned issue, but this option may have the least effect compared to the other three.

**Chart 17.**
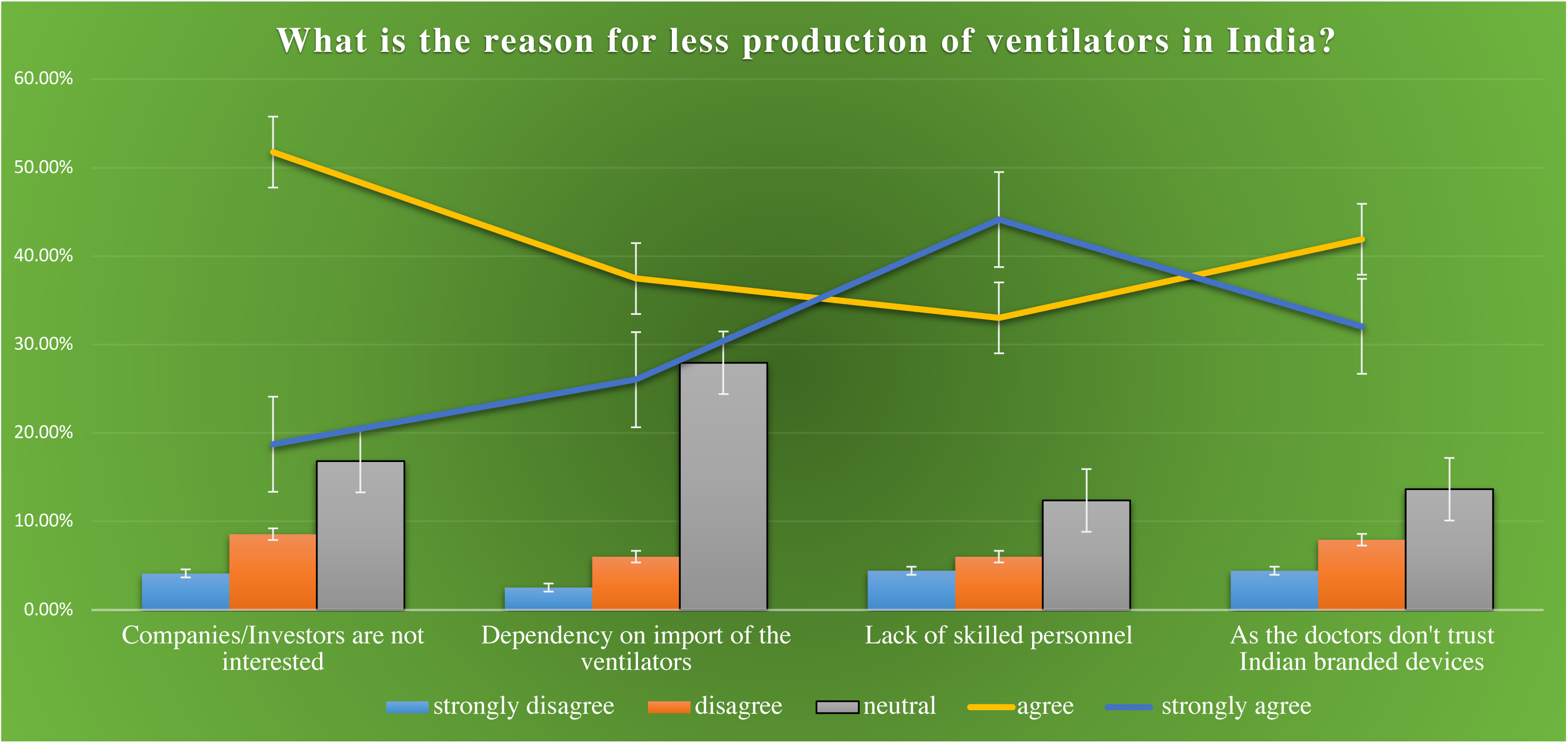
The percentage of responses recorded for each possible reason for less production of ventilators in India.

Till COVID-19 emerged, the ventilator industry hasn’t got much serious attention from investors. Also, India was initially dependent on countries like China, the USA, etc., for the import of raw materials of ventilators or its complete assembly. The number of companies who manufacture ventilators was also very few at the start; hence there was a lack of skilled people to initiate rapid production of ventilators.

The doctors also mainly don’t trust Indian branded devices and prefer the imported ones because of their high performance and other standard quality attributes. The overall outcome of these reasons was nothing but the low production of the ventilator device.

This question was designed to determine the major root cause of less production of ventilators based on respondents’ opinions. The survey results showed that the respondents feel that lack of Industry personnel is the important cause behind this issue. While they also think that least companies’/investors’ interest and doctors’ distrust of the Indian brands are possible reasons for less production. A small portion of people believe dependency on imports is also the cause of the mentioned issue, but it has less significance than other mentioned options. However, it again proved that people are less aware of the fact that ventilators are majorly exported from foreign countries.

#### 3.2.5. Possible solutions for Barotrauma- a Ventilator-associated problem

Pulmonary barotrauma, a complication of mechanical ventilation, is the presence of extra alveolar air in locations where it is not present under normal circumstances. The patients on mechanical ventilation ventilate with positive pressures, in contrast to the natural mechanism of breathing in humans, which depends on negative intrathoracic pressures. This may elevate the trans-alveolar pressure or the difference in pressure between the alveolar pressure and the pressure in the interstitial space, resulting in leakage of air into the extra-alveolar tissue. Barotrauma is most commonly due to alveolar rupture, which leads to an accumulation of air in extra alveolar locations. Excess alveolar air could then result in complications such as pneumothorax, pneumomediastinum, and subcutaneous emphysema. The incidence of barotrauma in patients receiving invasive mechanical ventilation is much higher than patients receiving non-invasive mechanical ventilation. Patients at increased risk of developing barotrauma from mechanical ventilation include individuals with predisposing lung pathology such as chronic obstructive pulmonary disease (COPD), asthma, interstitial lung disease (ILD), and acute respiratory distress syndrome (ARDS). However, certain ventilator settings, as well as specific disease processes, may increase the risk of barotrauma significantly. When managing a ventilator, physicians and other health care professionals must be aware of these risks to avoid barotrauma [13].

The intention for preparing this question was to determine the respondents’ perception of the possible soultions of Barotrauma proposed as in the options.

The coefficient of alpha value .719 denoted high reliability of data (from Cronbach’s alpha test), making it suitable for further analysis.

**Table 14** demonstrates the values of central tendency calculated from the respondents’ data, where the respondents agreed to all options determined from mean values.

**Table 14.**
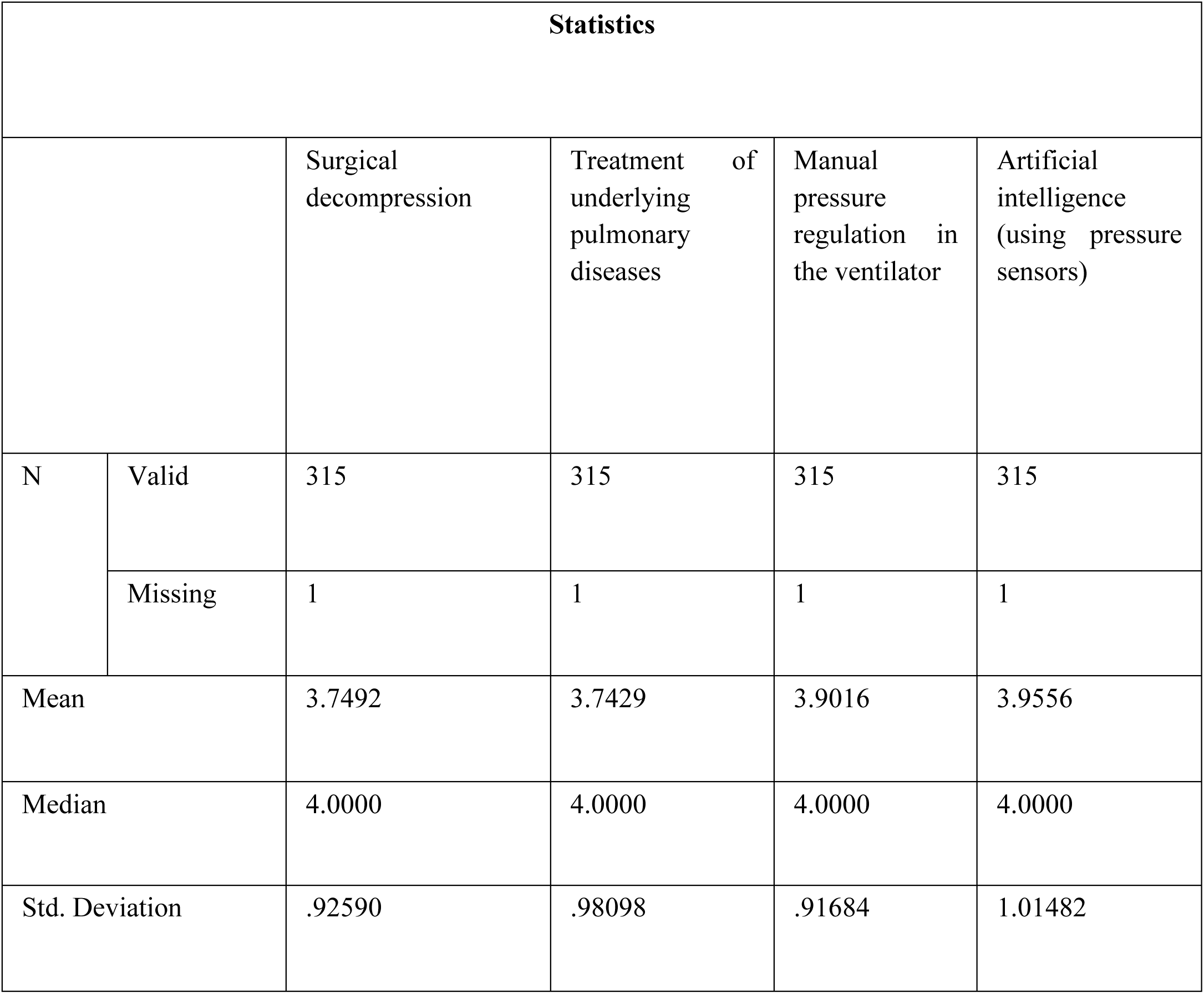
The measures of central tendency.

The frequency characteristics for each of the possible preventive measures of Barotrauma estimated from the respondents’ data are discussed in **Fig 20** and **Fig 21**.

**Fig 20.**
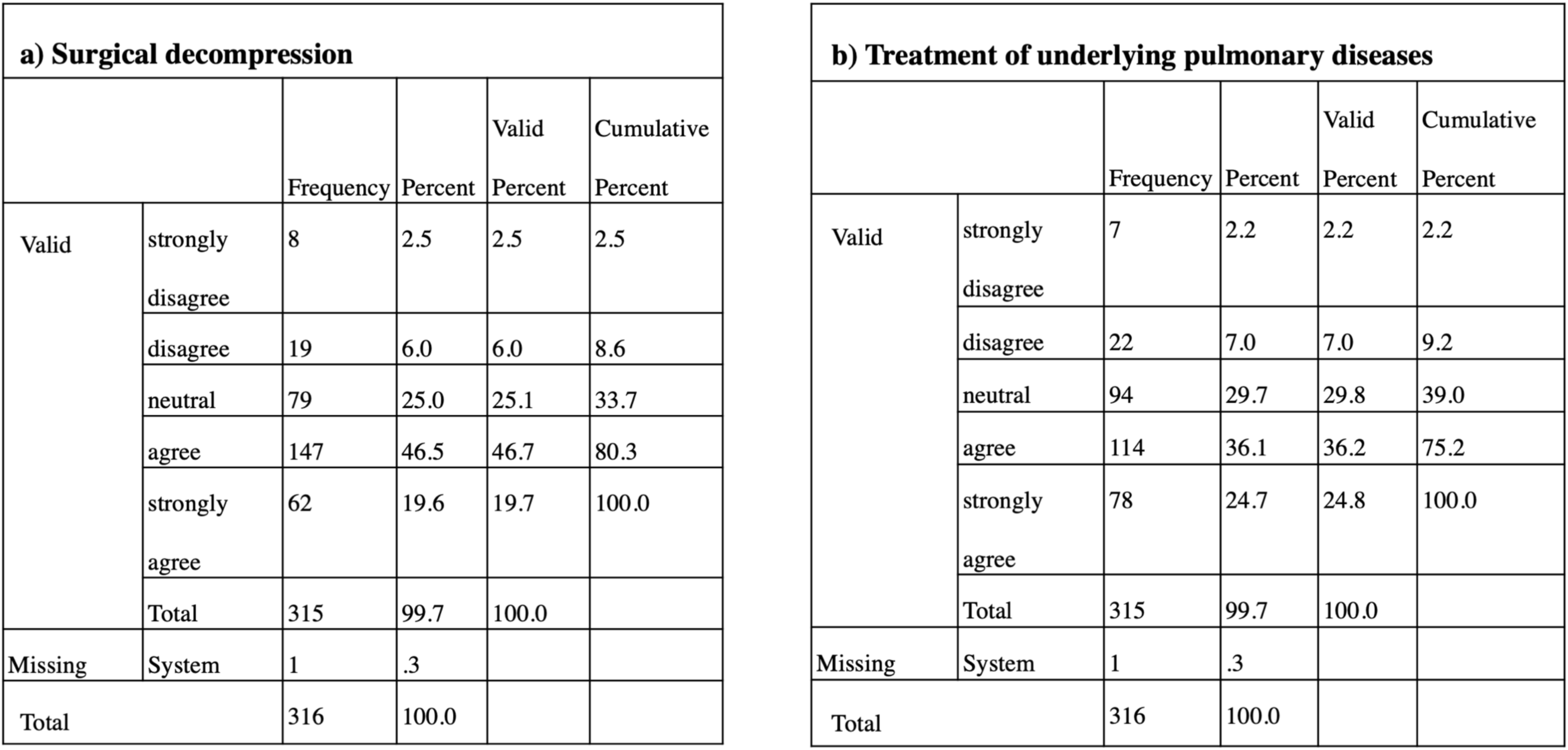
The frequency statistics of (a) Surgical decompression (b) Treatment of underlying pulmonary diseases are calculated from the data of respondents.

**Fig 21.**
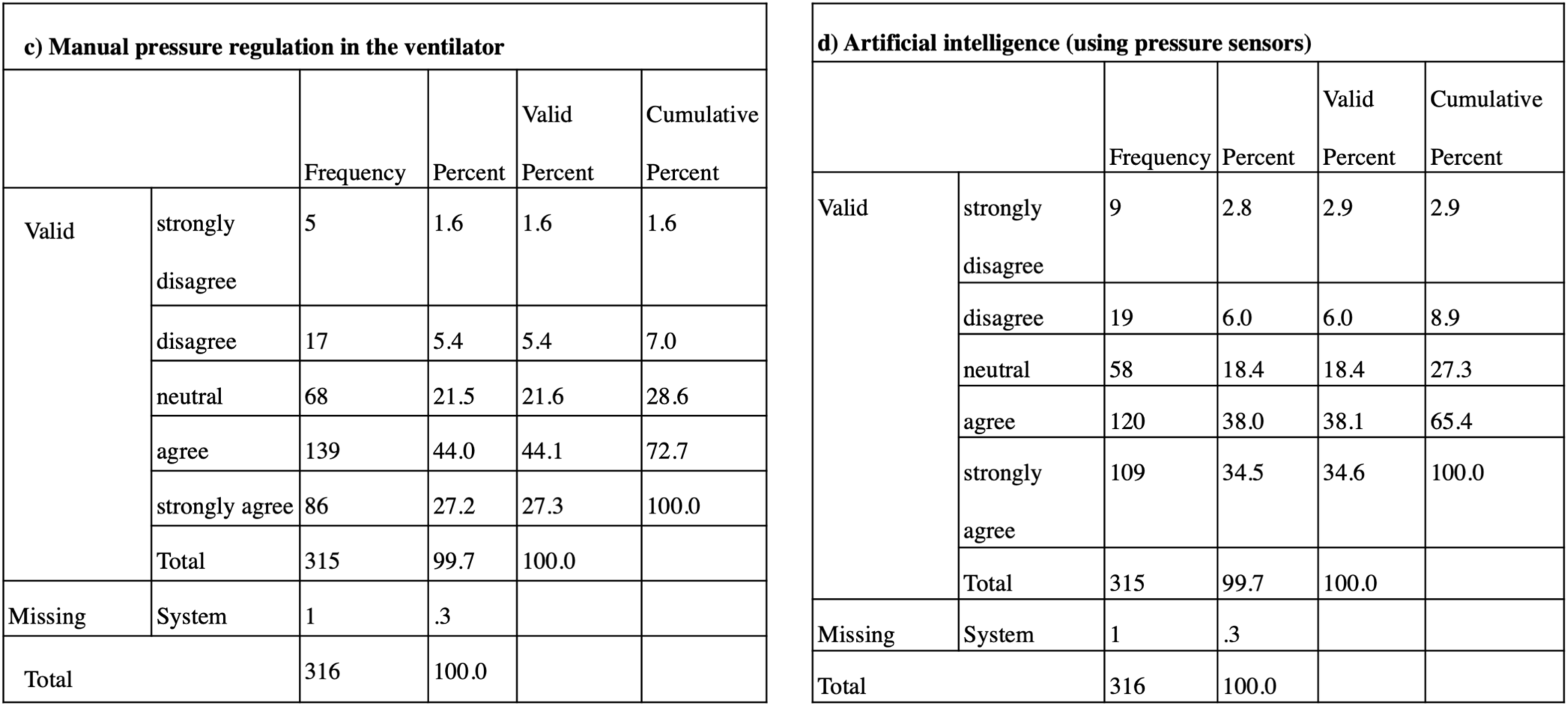
The frequency statistics of (c) Manual pressure regulation in the ventilator (d) Artificial intelligence (using pressure sensors) acquired from the respondents’ data.

Positive mechanical ventilation leads to the development of massive pressure inside the lungs, causing barotrauma. Thus, this pressure requires removal via surgical means, often referred to as surgical decompression [31]. According to the collected data (**Chart 18**), 46.67% of respondents have agreed with surgical decompression, making it the most suitable way of treating Barotrauma from all other probable solutions.

**Chart 18.**
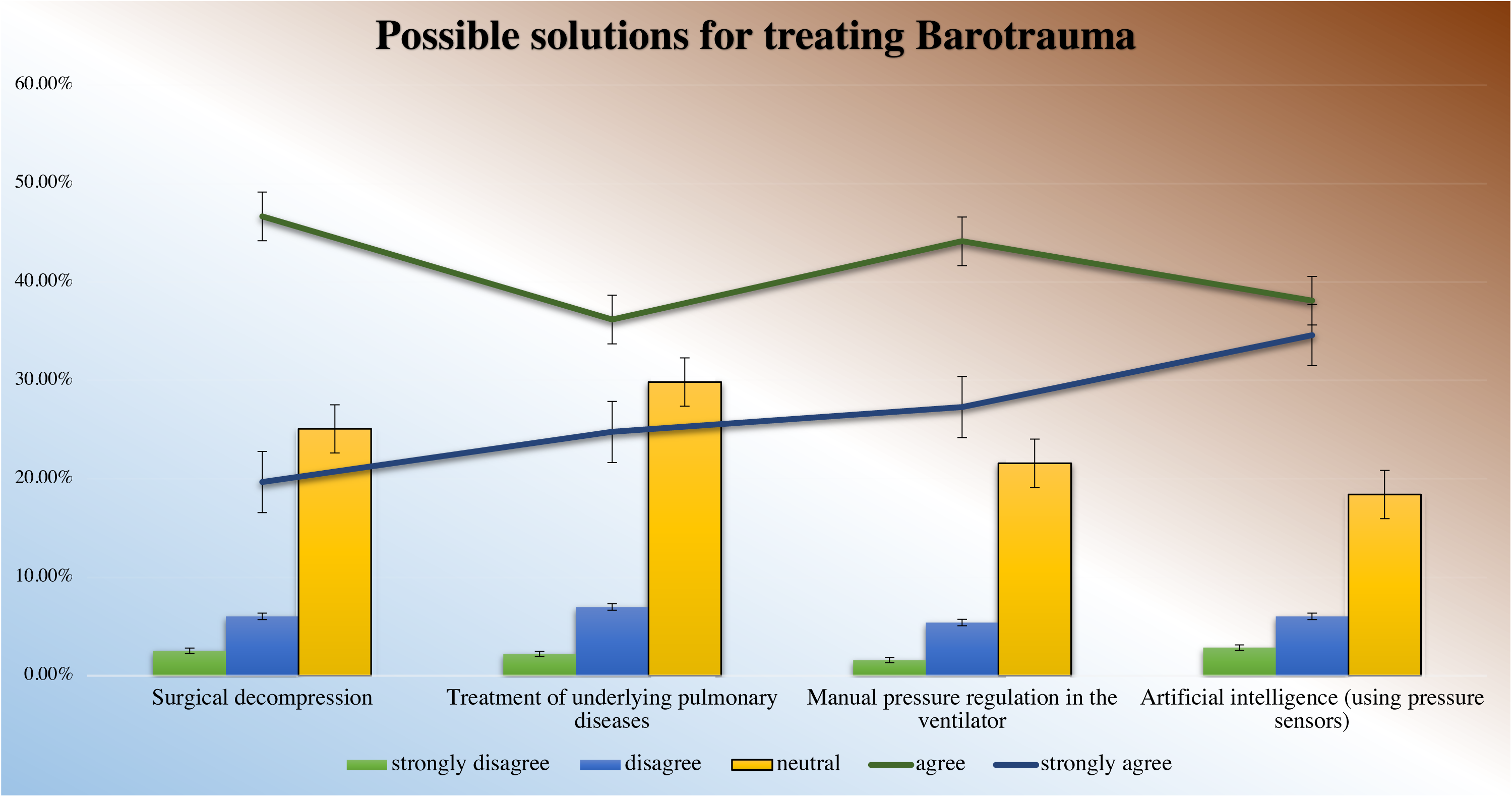
The percentage of responses received for each possible solution for treating barotrauma in the survey.

Also, 44% of respondents agreed to control the pressure manually in the ventilator. The survey participants also echoed that adjustment of proper pressure regulation according to the patient’s requirement is necessary to avoid the development of extra pressure leading to any complications.

Artificial intelligence (AI) has currently grasped the healthcare market with its exceptional capabilities to treat diseases. Thus, applying the AI (pressure sensors) could sense the patient’s pressure, and the ventilator could be adjusted according to it. This diminishes the chances of developing any extra pressure, which might be detrimental to the patient’s health. Though the AI system has not still disseminated in a broad population, an average of 34.56% (agreed and strongly agreed) respondents considered it as one of the possible solutions for the same.

Patients with predisposing lung pathology such as COPD, asthma, ILD, and ARDS are at high risk of developing barotrauma. Treatment of these ailments can reduce the chances of developing barotrauma, thus preventing alveolar rupture [13]. 36.1% of respondents confirmed this as one of the possible solutions for the same.

Thus, we observed that many respondents have agreed with all four solutions (with a very less significant difference), and very few people have strongly disagreed with all four possibilities (on an average 6.3%).

#### 3.2.6. Possible solutions for Ventilator-associated Pneumonia (VAP)

Ventilator-associated pneumonia (VAP) remains a feared complication in ICU and high postoperative surgical patients. VAP is associated with substantial excess morbidity. On average, 10-20% of ICU patients ventilated for less than 2 days experience VAP. VAP prevention targets the primary pathogenic mechanism, which is bacterial translocation from the stomach and oropharynx to the lower respiratory tract. Within hours following endotracheal intubation, pathogenic microorganisms colonize the oropharyngeal mucosal surfaces, dental plaque, sinuses, and stomach. Accumulation of oropharyngeal secretions colonized with these pathogens occurs above the endotracheal tube (ETT) cuff. Generally, VAP prevention focuses on reducing the exposure time, maintaining oral hygiene by antiseptic rinsing, and avoiding microaspiration [13].

The main motive behind the question was to understand the perception of the respondents on the proposed solutions for treating VAP.

The Cronbach alpha test resulted in a coefficient of alpha .849, which considered the data highly reliable. Further analysis was carried out with the collected data to find out the observation of the respondents.

**Table 15** discusses the values of central tendency calculated from the data of the respondents, where all respondents agreed to all preventive measures, as interpreted from the mean values.

**Table 15.** The measures of central tendency.

| Statistics |  |  |  |  |  |  |  |  |
| --- | --- | --- | --- | --- | --- | --- | --- | --- |
|  |  | Nebulized and broad-spectrum antibiotics | Subglottic secretion drainage (SSD). | Antibacterial coating in the ET tube | Introducing non-invasive positive pressure ventilation | Daily weaning trials and sedation holidays | Early tracheostomy | Elevation of head of the bed to reduce aspiration of gastric content |
| N | Valid | 315 | 315 | 315 | 315 | 315 | 315 | 315 |
|  | Missing | 1 | 1 | 1 | 1 | 1 | 1 | 1 |
| Mean |  | 3.8635 | 3.7302 | 3.9079 | 3.8349 | 3.7873 | 3.8762 | 3.8984 |
| Median |  | 4.0000 | 4.0000 | 4.0000 | 4.0000 | 4.0000 | 4.0000 | 4.0000 |
| Std. Deviation |  | .90492 | .94104 | .93470 | .95340 | .98192 | .94796 | 1.00119 |

The frequency characteristics of each of the probable preventive approaches for treating VAP calculated from the respondents’ data are discussed in **Fig 22** and **Fig 23**.

**Fig 22.**
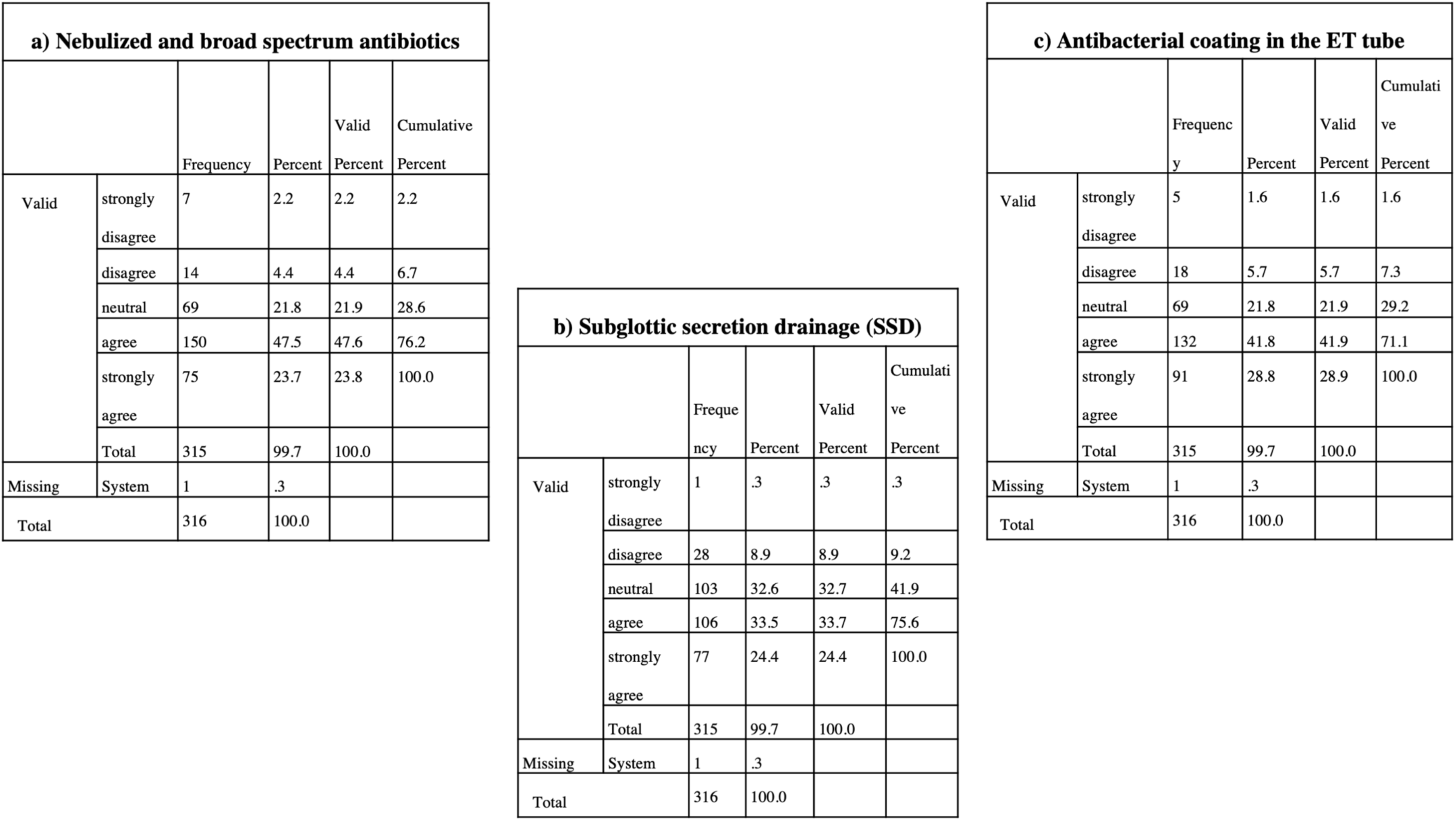
The frequency statistics of (a) Nebulized and broad-spectrum antibiotics (b) Subglottic section damage (c) Anti-bacterial coating in the ET tube calculated from the data acquired from the respondents.

**Fig 23.**
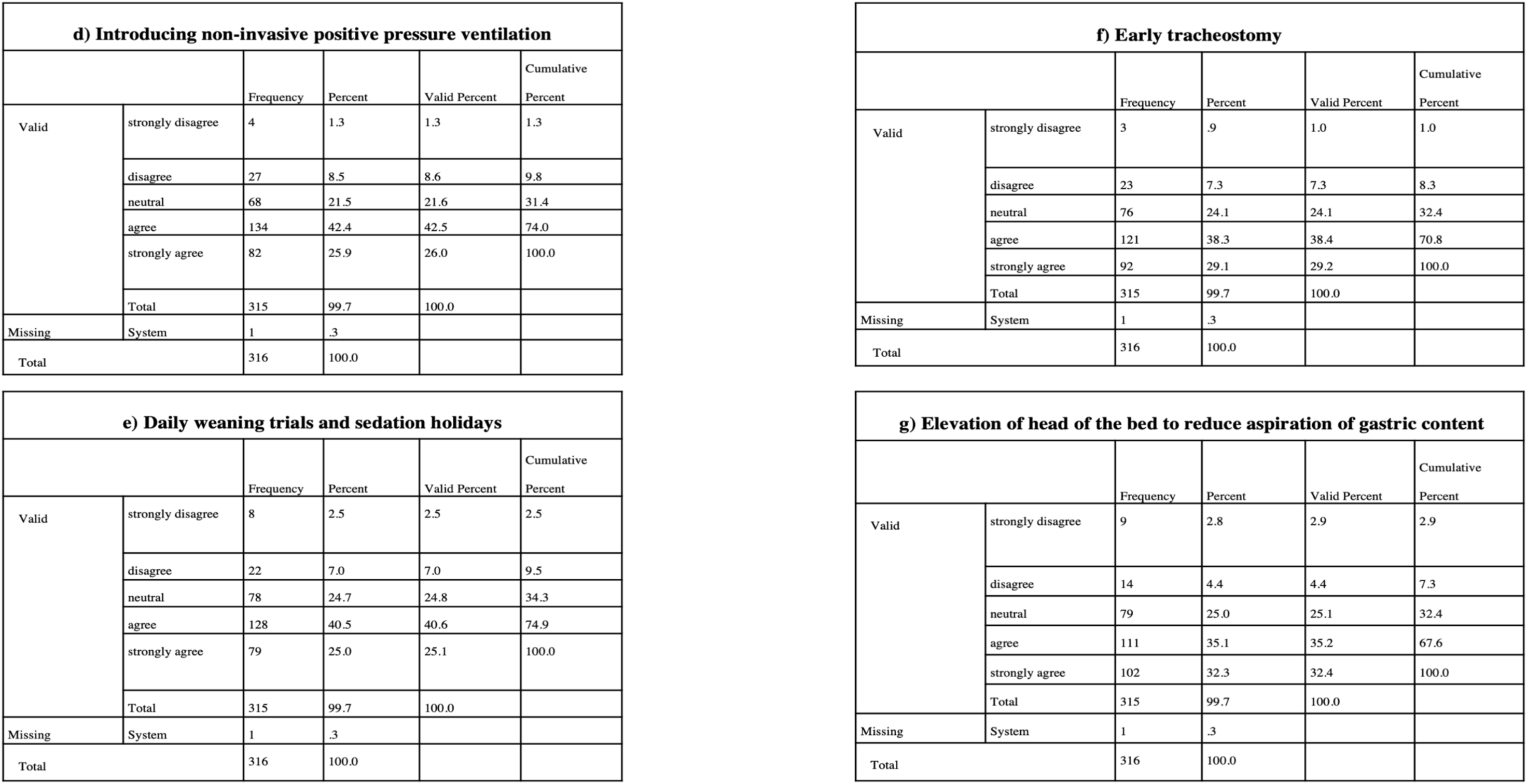
The frequency statistics of (d) Introducing non-invasive positive pressure ventilation (e) Daily weaning trials and sedation holidays (f) Early tracheostomy (g) Elevation of the head of the bed to reduce aspiration of gastric content measured according to the responses received from the respondents.

Broad-spectrum intravenous antibiotics and nebulized antibiotics could represent an attractive alternative for VAP treatment, mainly for cases caused by MDR Gram-negative bacteria and other classes. As MDR pathogens are only gullible to traditional antibiotics associated with adverse effects, nebulized therapy is specifically symbolized as a fascinating approach for VAP treatment [32], [33]. According to the collected data (**Chart 19**), 47.5 % of the respondents agreed with the same and made it the superior solution for the treatment.

**Chart 19.**
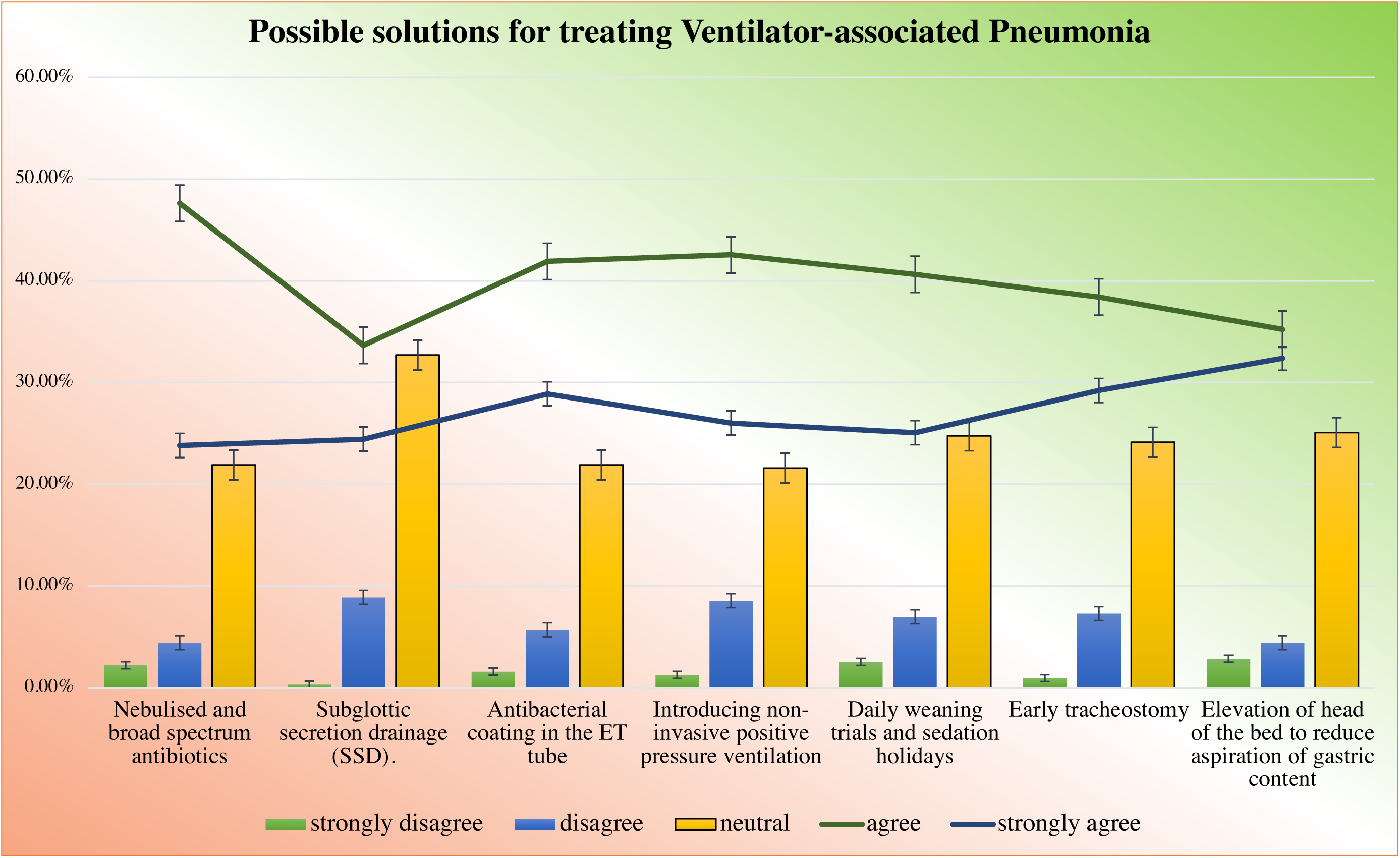
The percentage of responses received from the respondents for each possible solution for Ventilator-associated pneumonia.

Antibiotics for treating lower respiratory tract infections are not always effective and can result in increased antibiotic resistance. Prophylactic approaches by altering the surface of ETTs to prevent the growth and establishment of bacteria have exhibited promising results. Besides, surface modifications with bacteriostatic properties or active coatings that possess a bactericidal effect have been proven effective [34]. VAP can occur through impairment of cough and mucociliary clearance, accumulation above and leaking around the cuff of contaminated secretions, or increased bacterial binding to the surface of the bronchial epithelium. With non-invasive positive pressure ventilation, the risk of aspiration of colonized or infected oropharyngeal secretions is likely to be reduced, which is experimentally proven [35]. Of the above two proposed solutions, on average, 41.86% of respondents agreed to them as the probable solution for the treatment of VAP.

The primary routes of bacterial entry into the lower respiratory tract are the aspiration of oropharyngeal pathogens and leakage of subglottic secretions containing bacteria around the endotracheal tube cuff. Subglottic secretion drainage (SSD), using a specially designed endotracheal tube with a separate dorsal lumen that opens immediately above the endotracheal cuff, has been developed to prevent the occurrence of VAP [36]. 33.5% and 24.4% of respondents agreed and strongly agreed to subglottic secretion drainage as the possible solution for the same, respectively.

Daily weaning trials and sedation holidays are validated as strategies that limit the time of mechanical ventilation. As the risk of VAP is related to the duration of mechanical ventilation, limiting this duration makes physiologic sense with regards to reduction in VAP rates [37]. 40.5% of respondents have agreed this strategy to be beneficial for the same.

Long-term ventilation leads to an increased risk of mechanical complications, such as tube disconnection, mucus impaction, laryngeal and tracheal ulcers, and increased rates of VAP [38]. This can be prevented by early tracheostomy, and 38.3% of respondents also agreed to the same.

It has been suggested that placing critically ill ventilated patients in a semi-recumbent position reduces the incidence of developing VAP. Elevating the head of the bed to 45° solves the problem [39]. On average, 35.1% of respondents resonated the same for the treatment of VAP.

Thus, we observed that many respondents have agreed with all the solutions, and very few people have strongly disagreed with all the four possibilities (8.90% of respondents strongly disagreed).

#### 3.2.7. Possible solutions for Microaspiration

Microaspiration of subglottic secretions through channels formed by folds in high-volume low-pressure cuffs of endotracheal tubes is considered an important pathogenic mechanism of VAP. A series of preventive measures target the avoidance of microaspiration. Accumulation of oropharyngeal secretions colonized with the pathogens occurs above the endotracheal tube cuff. Microaspiration of these subglottic secretions might occur through an underinflated tracheal cuff or through longitudinal folds in high volume-low pressure cuffs. Furthermore, a nasogastric tube may facilitate gastroesophageal reflux. Therefore, gastric juice may be aspirated into the lungs, provoking local inflammation. An analogous conclusion can be made regarding taper-shaped cuffs compared with classic barrel-shaped cuffs. The clinical usefulness of endotracheal tubes developed for subglottic secretions drainage is established in multiple studies and confirmed by meta-analysis. Any change in cuff design will fail to prevent microaspiration if the cuff is insufficiently inflated. Gel lubrication of the cuff before intubation temporarily hampers microaspiration through sludging the channels formed by folds in high volume-low pressure cuffs [12].

The question’s primary motion was to check if the respondents resonated the same for the proposed solution to treat microaspiration.

The coefficient of alpha .848 calculated from the Cronbach alpha test proved the collected data to be highly reliable, paving the way for further data analysis.

**Table 16** discusses the measures of central tendency estimated from the respondents’ data. All the respondents agreed with all the preventive measures, as interpreted from mean values.

**Table 16.**
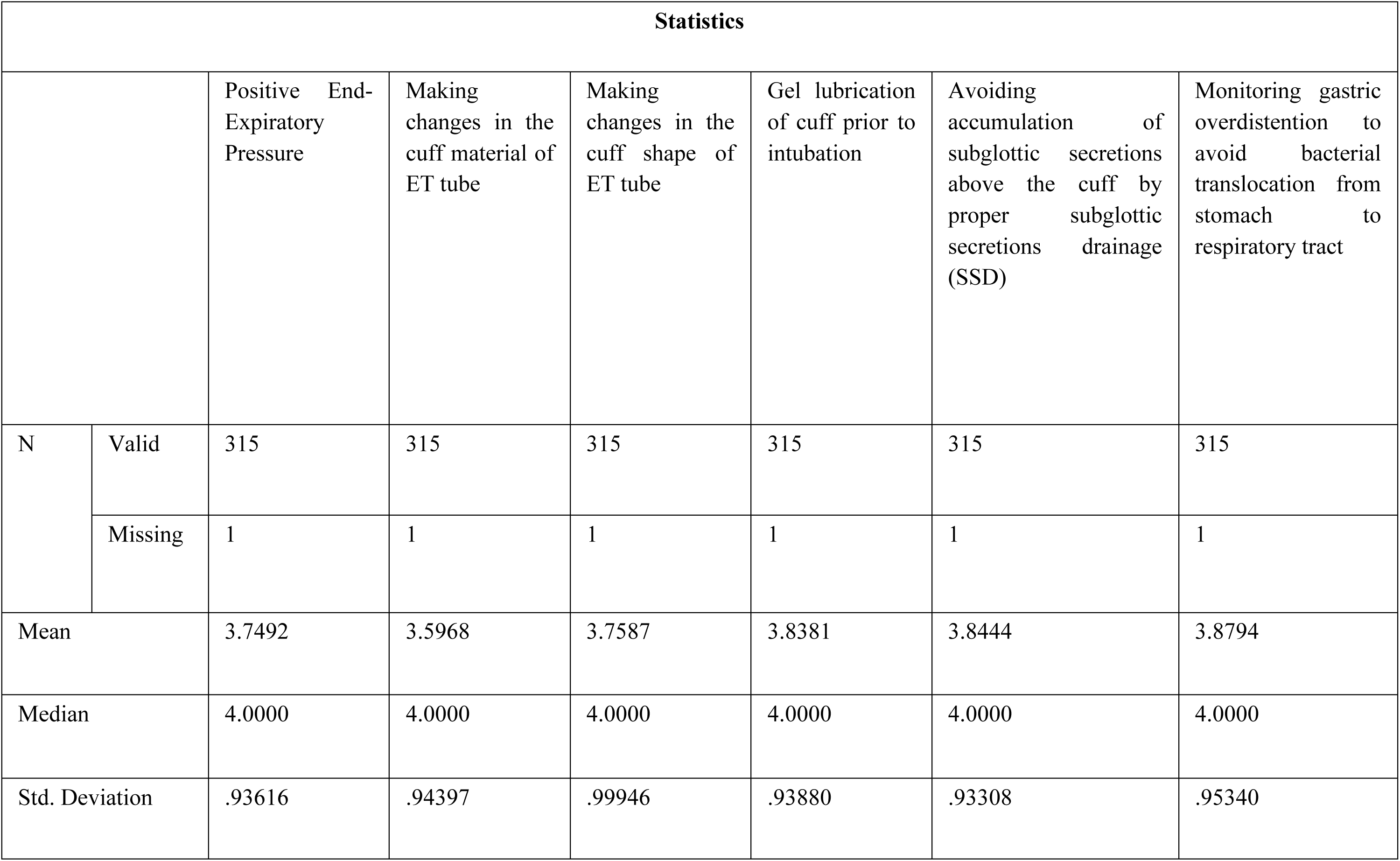
The measures of central tendency.

The frequency characteristics for each of the preventive measures to treat microaspiration calculated from the respondents’ data are discussed in **Fig 24** and **Fig 25**.

**Fig 24.**
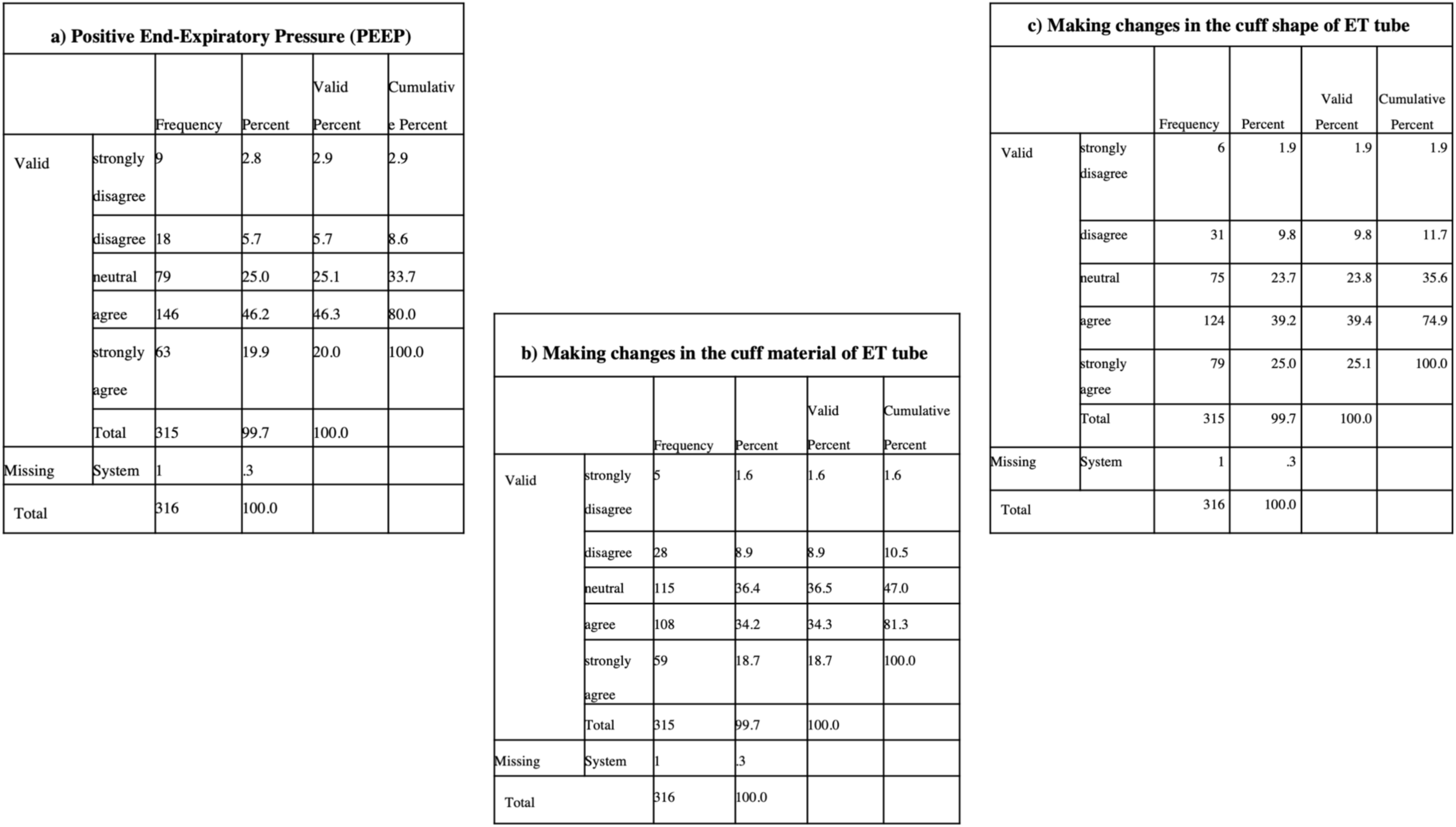
The frequency statistics of (a) Positive end-expiratory pressure (PEEP) (b) Making changes in the cuff material of ET tube (c) Making changes in the cuff shape of ET tube estimated from the responses recorded from the survey.

**Fig 25.**
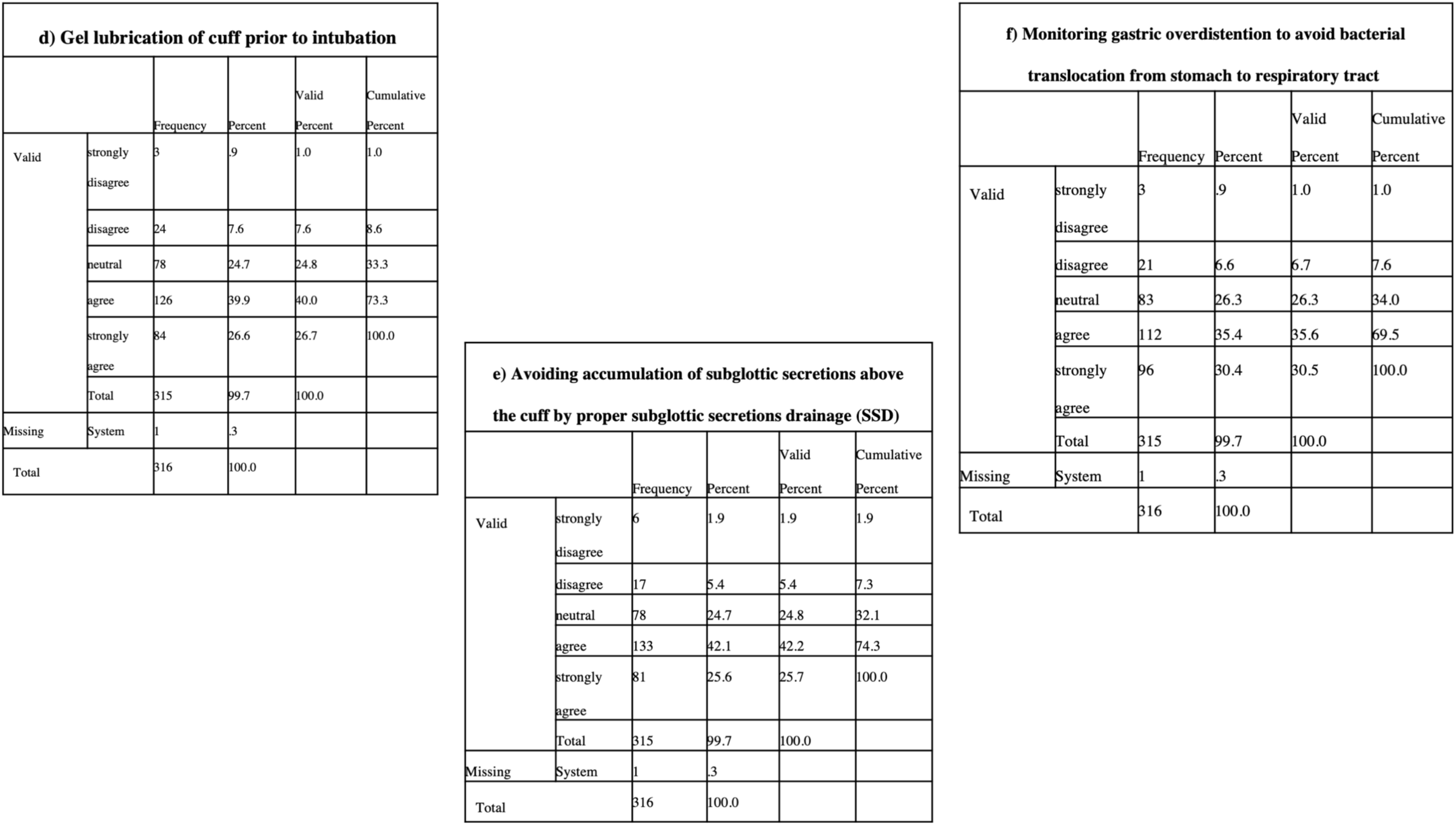
The frequency statistics of (d) Gel lubrication of cuff prior to intubation (e) Avoiding accumulation of subglottic secretions above the cuff by proper SSD (f) Monitoring gastric overdistention to avoid bacterial translocation from stomach to respiratory tract acquired from the data collected through the survey.

The gas contained within the cuff is redistributed from the distal to the proximal cuff end as the air pressure rises, resulting in a cone-shaped cuff in which the intra-cuff pressure is temporarily (during the inspiratory phase) higher than the cuff pressure during the expiratory phase. Also, positive pressure ventilation creates a ‘self-sealing’ effect by which tracheal occlusion is maintained despite airway pressure exceeding intra-cuff pressures. Therefore, it was hypothesized that PEEP could result in a better sealing capacity throughout the ventilation cycle and reduce micro-aspiration [40]. And, from the collected data (**Chart 20**), 46.2% of respondents agreed to it as the superior solution for treating the same.

**Chart 20.**
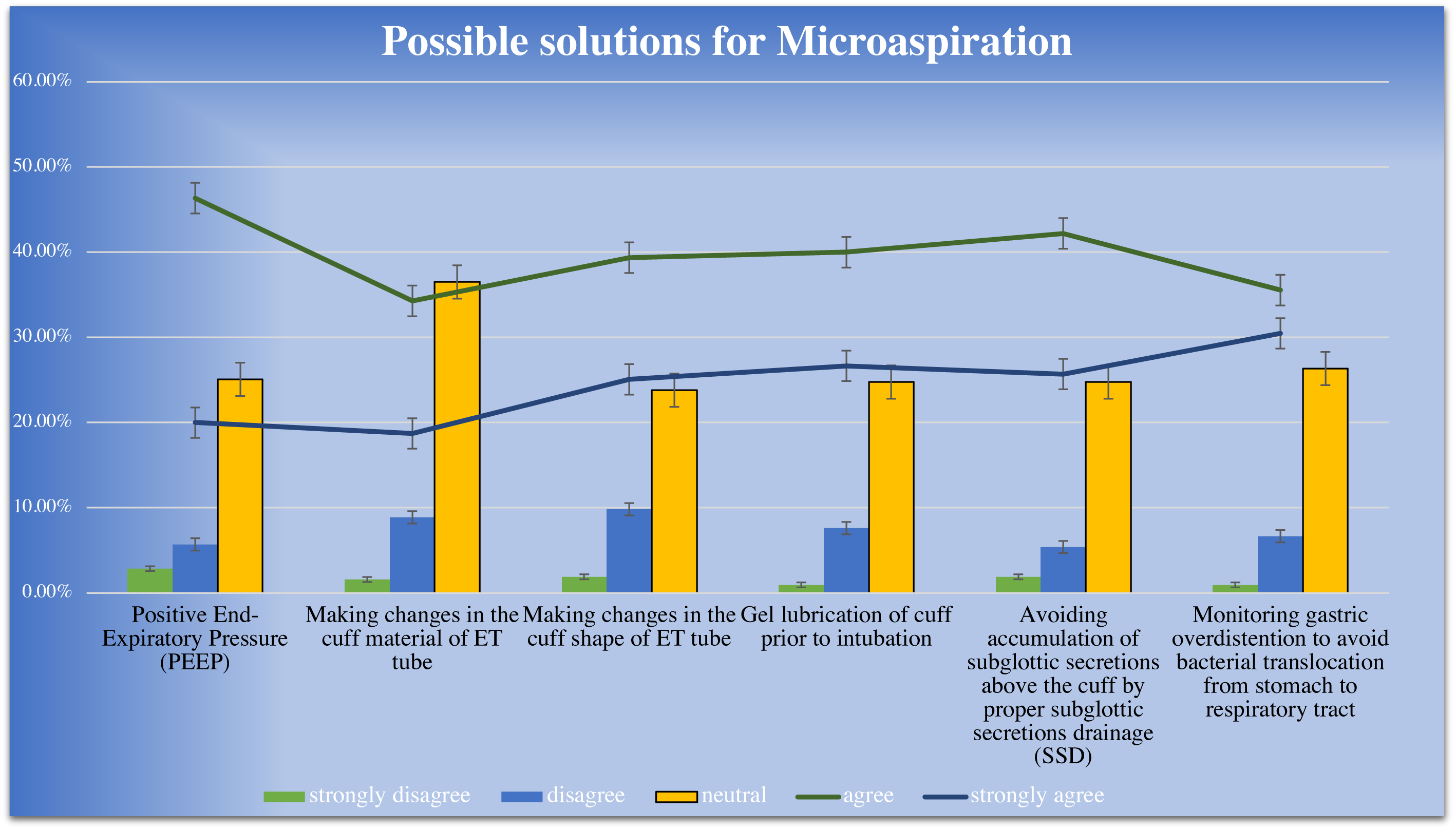
The percentage of responses recorded for each of the possible solutions for treating Microaspiration.

Conventional ETTs have a cylindrical-shaped cuff. An ETT with a taper-shaped cuff was developed for the treatment of microaspiration. Because of its tapered shape, this cuff seals the trachea, at least at one point, without fold formation [41]. This also prevents fluid leakage, thus reduces microaspiration. 39.2% of respondents agreed with the same and echoed our proposed solution.

Likely, 39.9% of respondents agreed to gel lubrication of cuff prior to intubation as one of the strategies done to smoothen the procedure. Yet, by doing so, the channels along the cuff wall are plugged, thereby blocking microaspiration of oropharyngeal secretions [42].

ETTs for SSD can drain secretions through a separate dorsal lumen that opens directly above the cuff. The use of SSD was associated with a reduced ICU stay, decreased length of ventilatory dependence, and an increased time to the first episode of VAP [43]. 42.1% of respondents agreed to the same for the possible treatment of the microaspiration.

Gastric overdistention has been historically considered a risk factor for VAP as it is assumed to facilitate bacterial translocation from the stomach to the respiratory tract. Monitoring gastro-intolerance to enteral feeding by checking residual volumes is a frequent practice. The most frequent thresholds used to interrupt enteral feeding are residual volumes of 200–250 mL. Yet, cessation of enteral feeding is not recommended unless residual volumes exceed 500 mL [44]. This solution has been least agreed upon when compared with others. 35.4% of respondents agreed to it as one of the possible solutions for the same.

34.2% of respondents agreed to the material change of ETT, the option which has the least percentage of agreement. Recently, ultrathin polyurethane cuffs have been developed to minimize the channel size within folds of an inflated cuff. Polyurethane ETTs prevented fluid leakage more efficiently than PVC cuffs (p < 0.001).

Thus, we observed that many respondents have agreed with all four solutions, and very few people have strongly disagreed with all the solutions (on an average of 9.3%).

#### 3.2.8. Possible solutions to control Acute Respiratory Distress Syndrome (ARDS)

Acute respiratory distress syndrome (ARDS) is characterized by severe inflammatory response and hypoxemia. Worsening of this inflammatory response, called “ventilator-induced lung injury” (VILI), can be caused by the use of mechanical ventilation (MV) for correction of gas exchange. The process of withdrawing mechanical ventilation, referred to as weaning from MV, may cause worsening of lung injury by spontaneous ventilation. The initial evaluation and treatment of the patient with ARDS begin with the correction of the inflammatory mechanism that triggered the process and the decision to start ventilatory support, which can be provided as supplemental oxygen, high-flow nasal oxygen therapy, and non-invasive MV and in most cases invasive MV. The choice of ventilatory mode (controlled versus spontaneous) and parameter settings to adopt the “lung-protective ventilation” strategy are other essential aspects. This protective ventilation consists of the adjustment of low tidal volume based on the weight and elevated levels of Positive end-expiratory pressure (PEEP) with Respiratory regulation (RR). In the most severe cases, the use of a Neuro-muscular blockade (NMB), prone position, and Extra-corporeal membrane oxygenation (ECMO) will be evaluated when there is refractory hypoxemia. These initial strategies are the key to successfully treating ARDS and therefore weaning success and reducing MV time [14].

The significance of the question is to understand the perception of the respondents on the possible solutions presented for the treatment of ARDS.

The coefficient alpha .760 estimated from the Cronbach alpha test ensured that the data obtained is reliable, opening the window for further data analysis.

**Table 17** depicts the values of central tendency estimated from the respondents’ data. And, the respondents agreed to all the possible solutions, as interpreted from the mean values.

**Table 17.** The measures of central tendency.

| Statistics |  |  |  |  |  |
| --- | --- | --- | --- | --- | --- |
|  |  | Adopting lung protective ventilation strategies (Low tidal volume and low inspiratory pressure) | Using neuromuscular blocking agents (NMBA) at early stages | Monitoring asynchrony | Prone positioning of patient for more than 12 hours |
| N | Valid | 315 | 315 | 315 | 315 |
|  | Missing | 1 | 1 | 1 | 1 |
| Mean |  | 3.7746 | 3.6286 | 3.9111 | 3.5873 |
| Median |  | 4.0000 | 4.0000 | 4.0000 | 4.0000 |
| Std. Deviation |  | .86877 | .95361 | .92988 | .96491 |

The frequency characteristics of each of the probable measures to treat ARDS calculated from the respondents’ data are discussed in **Figures 26** and **27**.

**Fig 26.**
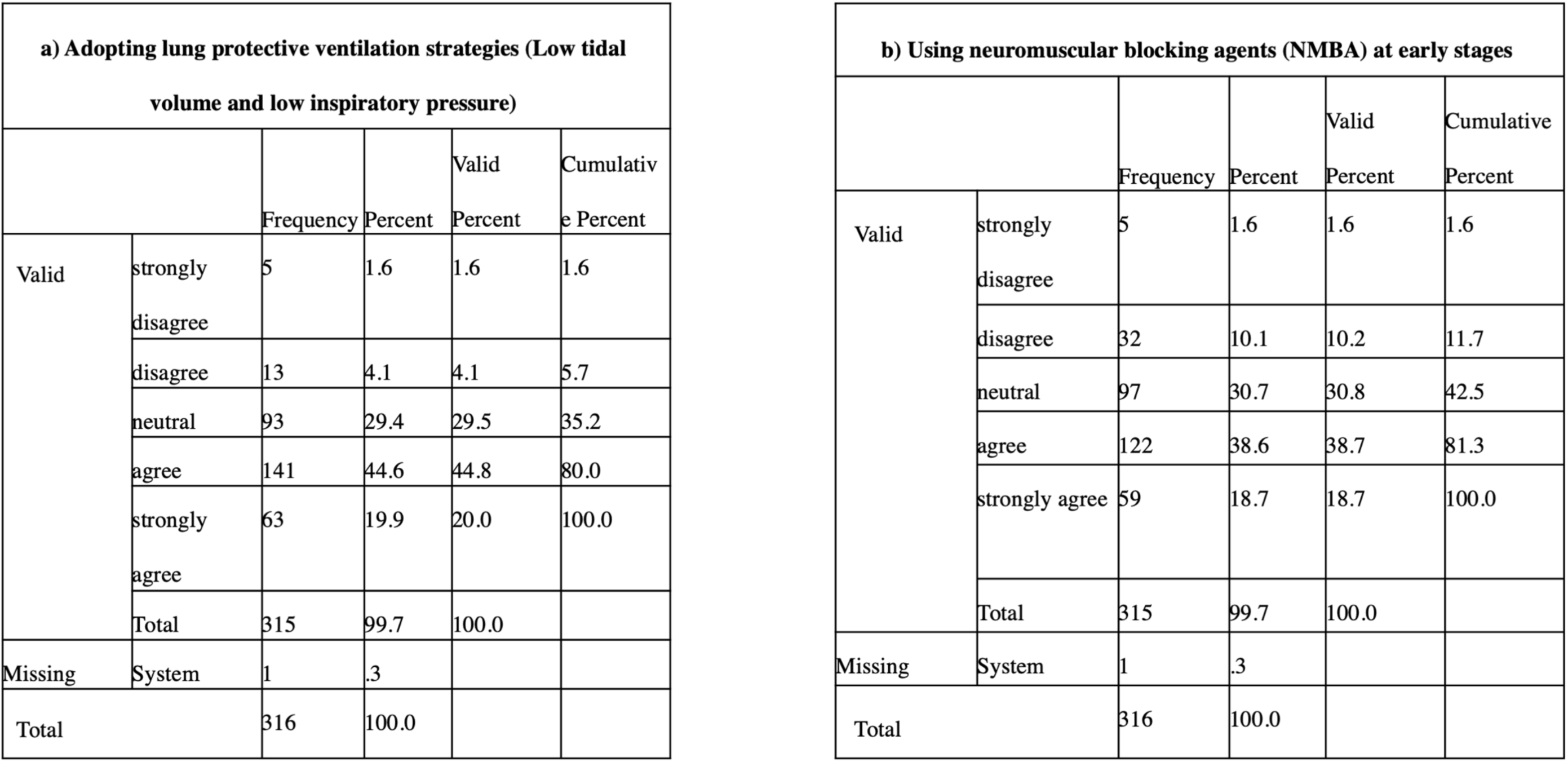
The frequency characteristics of (a) Adopting lung protective ventilation strategies (b) Using NMBA at early stages calculated from the responses received from the survey.

**Fig 27.**
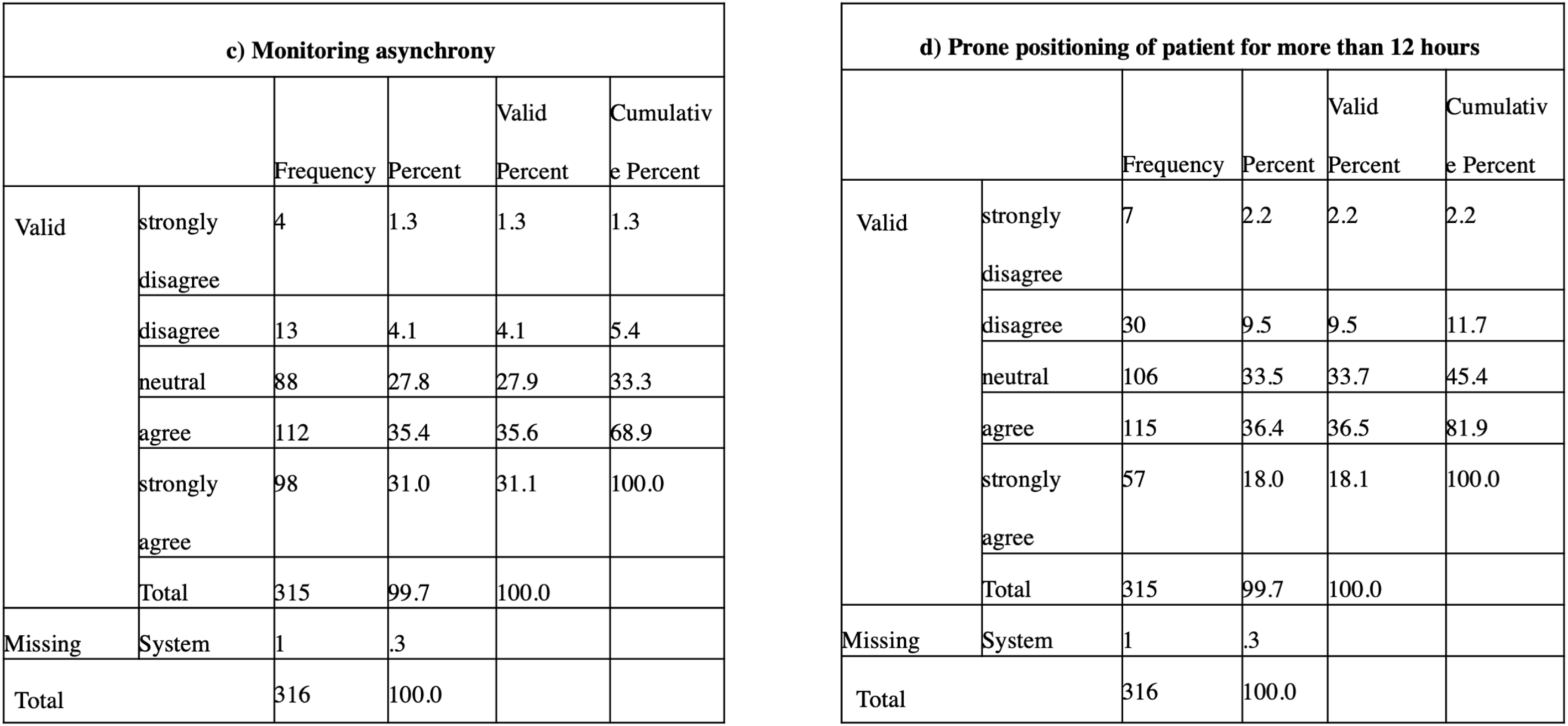
The frequency characteristics of (c) Monitoring asynchrony (d) Prone positioning of patient for more than 12 hours estimated from the data collected via survey.

In ARDS, there is an inflamed lung with increased levels of proinflammatory mediators, and ventilation can also enhance the amount of inflammatory mediators produced by the lungs. Once the barrier function of the alveolar-capillary membrane is lost, there is leakage of mediators to the circulation (decompartmentalization), leading to multi-organ failure. The use of lung protective ventilation reduces the cyclic collapse of the lung by minimizing the inflammatory response and preserving the barrier function of the lungs [45]. From the collected data (**Chart 21**), 44.6% of respondents agreed with this solution, mostly for the treatment of ARDS.

**Chart 21.**
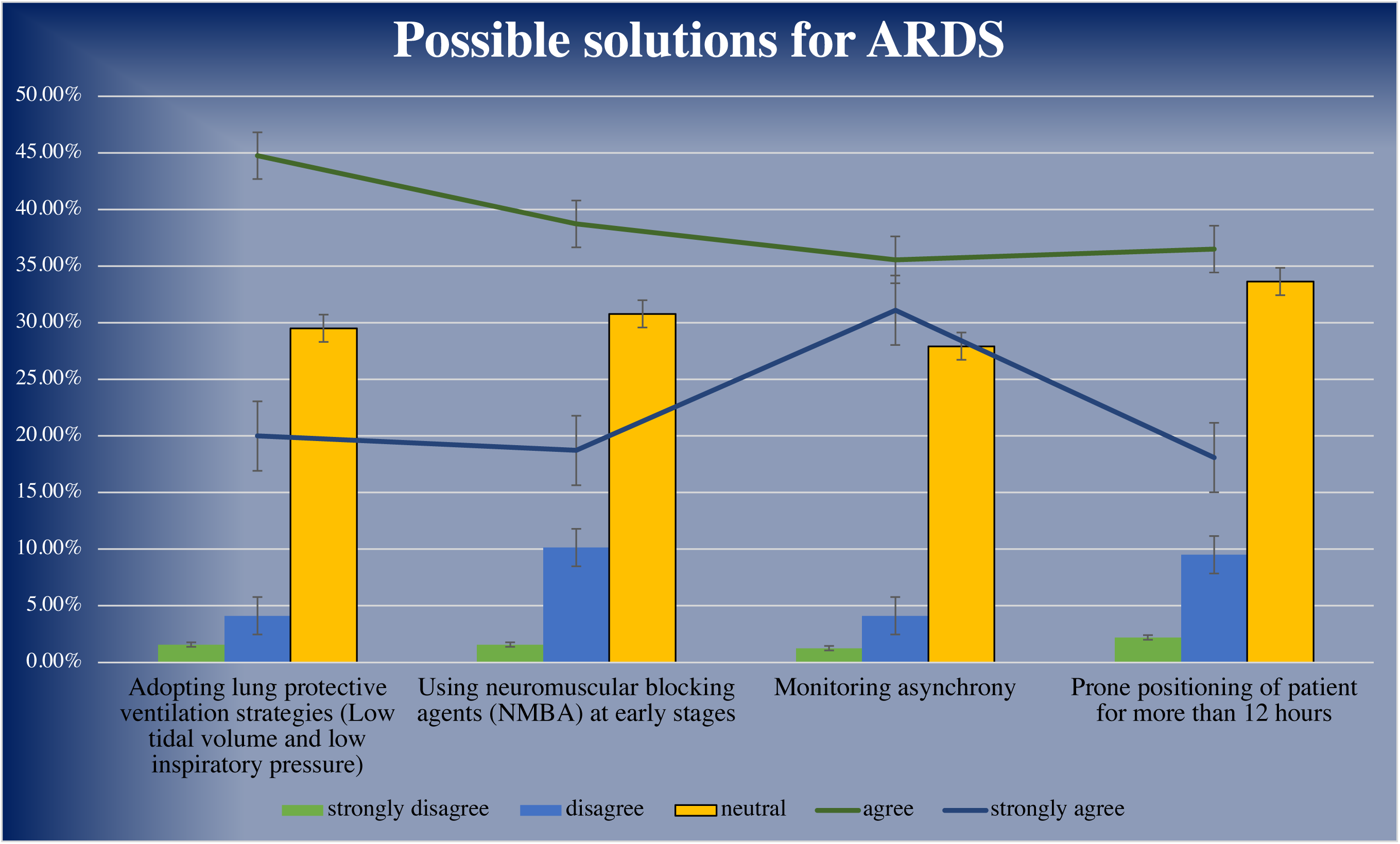
The percentages of responses documented from the respondents’ data for each probable solution for treating ARDS.

NMB agents are considered beneficial as one of the preventive measures for ventilator-induced lung injury. By resting the respiratory muscles, NMBA can avoid high regional transpulmonary pressures and induce less regional volutrauma or biotrauma. The use of NMBA can also avoid dangerous respiratory entrainment (“reverse-triggered” breaths) and pendelluft phenomena [46]. The same solution has been agreed by 38.6% of respondents for preventing ARDS.

Prone-positioning (PP) maintains a safe gaseous exchange. In PP, the transpulmonary pressure and ventilation are more homogeneously distributed throughout the lungs. Overall, lung stress and overdistension are minimized, lung volume increases, and biotrauma and ventilator-induced lung injury are decreased by PP [46]. 36.4% of respondents agreed with this technique as a possible treatment for ARDS.

Ventilator asynchrony is associated with increased ICU stay and mortality. Episodes of asynchrony may occur, worsening hypoxemia and increased respiratory muscle work. Asynchrony may cause large transpulmonary pressure swings that may be especially harmful in critically ill patients receiving lung-protective ventilation. The presence of asynchrony during the weaning of ARDS patients from MV could be better quantified and distinguished. The presence of asynchrony, both quantity, and type, could be considered a predictor of MV weaning failure in patients with ARDS [14]. 35.4% of respondents agreed to monitor asynchrony for the treatment of ARDS, making it the least agreed from all the other possible solutions.

Thus, we observed that the maximum number of respondents have agreed with all the solutions, and very few people have strongly disagreed with all of them (on an average of 4.5%).

## 4. Conclusion

This research-based survey was done to grasp the current scenario, the strength, and shortcomings of the Indian medical device industry. The survey was also conducted to uncover the people’s perception of the Indian medical device industry as well as on the problems related to ventilators. Respondents were also given details of the problems related to the Indian biomedical industry/ventilators and asked to rate the best possible solutions that we came up with for the problems.

The survey questionnaire was prepared according to the best of our ability to understand the same aforementioned purpose. And it definitely gave an indication of many problems and supported our proposed solutions. 315 respondents took part in this survey ranging from graduates to Ph. D students. The first section of the survey consisted of the problems and solutions related to the biomedical industry of India. Many people agreed with the reasons due to which this industry is facing obstacles and also agreed with our proposed solutions that can be taken into account to make this industry better. The second section was based on one of the biomedical devices, i.e., ventilators, as it was one of the top requirements in this era of COVID-19. The frequent use of ventilators resulted in various pharmacological issues, which we tried to address through this survey. We have also tried to explore the main reasons for less production of ventilators and proposed groundbreaking solutions for the conditions associated with frequent use of ventilators. The results complied with our hypothesis as the majority of the respondents have agreed with almost all the probable solutions in both the sections given by us as options.

## 5. The way forward

India is a ground full of potential for parties in the medical devices industry. The country has also turned into one of the principal addresses for high-end diagnostic facilities with enormous capital investment for progressive diagnostic equipment, thus providing an increased proportion of the population. Moreover, Indian medical service customers have become more mindful of their healthcare maintenance.

The Indian healthcare sector is much multifarious and full of possibilities in every fragment, including suppliers, payers, and medical technology. With the rise in rivalry, businesses are sounding to investigate the latest dynamics and trends that will have an optimistic strike on their business. The whole research was primarily aimed at finding out about the Indian biomedical device industry’s current scenario. Although the Indian biomedical industry is one of the steadily growing markets with many potentials, there are still some loopholes present, like the absence of strong laws and authority. Not only this, but many underlying problems are associated with it. With this research-based survey, we aimed to bring about the matter to the hands of the people, mostly from the pharmacy/biomedical field, to understand the same. We tried our best to distribute this paper to a mass of people to be considered a problem and could be worked upon. Moreover, we had worked upon ventilators in India and the possible reasons for ventilators being scarce at the most important time, i.e., Coronavirus. We came up with possible changes that can be done to remove the current problems associated with the ventilators, like barotrauma, Ventilator-Associated Pneumonia (VAP), microaspiration, and ARDS. We desired to bring light upon this topic to bring it out to the masses and the probable solutions for it so that ventilators can be made better.

This research survey provides the in-detailed condition of the Indian biomedical industry as well as the ventilators, consisting of the perception of Indian respondents on our proposed solutions.

## Data Availability

Data available within the article or its supplementary materials

## Acknowledgment

The authors would like to thank Dr. Ramesh Parameswaran and Dr. Vignesh Muthuvijayan for their valuable support in conducting the survey. Author SP would like to thank the Indian Institute of Technology, Madras, for providing financial assistance and SCTIMST, Trivandrum, for providing workspace. The authors would also like to thank the IPGA student forum and Ms. Zeba Khan (President, IPGA-SF), Advisors of Erevna 3.0 (Dr. Atul Kumar, Mr. Atul Nasa, Dr. Vijay Bhalla, and Dr. Arun Garg), Project Dopamine team members (Project Directors: Mr. Dharmanshu Chaudhury, Mr. Rajesh Sharma; Project Head: Tanushree Jain; Project team: Mr. Abhinav Gupta, Ms. Shelly Kashyap, Ms. Puja Samanta, Ms. Gunjan Kumari).

## Credit Authorship Statement

Sheersha Pramanik: Conceptualisation, Resouces, Methodology, Investigation, Supervision, Writing: Original draft, Writing: Review and Editing; Sucheta Karmakar: Data analysis and curation, Writing-Original Draft; Shreyas Mukherjee: Data analysis and curation, Writing: Original draft; Indraneel Dhavale: Data analysis and curation, Writing: Original Draft; Rohan Shrestha: Writing-Original Draft, Resources.

## Abbreviations

MDI: Medical devices industry
SARS-CoV-2: Severe acute respiratory syndrome coronavirus 2 FDA-Food and Drug Administration
EU: European Union
CE: Conformité Européenne
COVID-19: Coronavirus disease 2019
ISO: International Organization for Standardization IT-Information Technology
IIT: Indian Institutes of Technology
ARDS: Acute respiratory distress syndrome CT-Computed tomography
MDR: Medical Device Reporting
EMA: European Medicines Agency US-United States
QMS: Quality management system Govt. – Government
DCGI: Drugs Controller General of India CL-Compulsory Licensing
USFDA: United States Food and Drug Administration COPD-Chronic Obstructive Pulmonary Disease
ILD: Interstitial Lung Disease
ARDS: Acute Respiratory Distress Syndrome AI-Artificial Intelligence
VAP: Ventilator-Associated Pneumonia ET-Endo-Tracheal Tube
ICU: Intensive Care Unit
SSD: Subglottic Secretion Drainage MDR-Multi-Drug Resistant
PEEP: Positive End-Expiratory Pressure PVC-Poly Vinyl Chloride
MV: Mechanical Ventilation
VLI: Ventilator-Induced Lung Injury
NMBA: Neuro-Muscular Blocking Agents RR-Respiratory Regulation
ECMO: Extra-Corporeal Membrane Oxygenation PP-Prone Positioning

