## Appendix 1 for "Unmasking the Current Scenario of Indian Biomedical Devices Industry: Ventilators being the Heart of the Discussion"

The Indian biomedical industry has been growing exponentially and has made a headway remarkably in recent years. Nevertheless, several hurdles need to be conveyed in providing access to quality, budget-friendly medical devices in India. But the Indian biomedical device industries continue to be affected by the aspects of availability, affordability, and quality of Indian devices. Thus, the ultimatum for Indian companies is to manufacture medical devices that are both economical and efficient to enhance penetration and use. Taking all into account, the "Make in India" trump-card becomes prominent for the medical devices industry.
The survey discusses the problems and solutions of the Indian Biomedical Industry followed by Ventilators in the second section. All of your responses will be kept confidential. We would like to thank you for spending your valuable time for the questionnaire. For any queries you can contact

*Required

1. Name *

2. Age *

*Mark only one oval.*

- 10-20
- 21-30
- 31-40
- 41-50
- >50

3. Educational Qualification *

*Tick all that apply.*

- [ ] Under-Graduate
- [ ] Graduate
- [ ] Post-Graduate
- [ ] Ph. D.

4. Have you ever heard of Biomedical devices? *

*Mark only one oval.*

- Yes
- No
- Maybe

5. Choose medical devices from below. (You can choose multiple answers) *

*Tick all that apply.*

- [ ] Hot Air Ovens
- [ ] CT Scanner
- [ ] Infusion pumps
- [ ] Weighing Machine
- [ ] X-Ray Scanner
- [ ] None of these

6. Do you know any Indian companies who make medical devices? *

*Mark only one oval.*

- Yes
- No
- Maybe

7. Which of the following are Indian biomedical companies? *

*Tick all that apply.*

- [ ] Phillips Healthcare
- [ ] Centenial Surgical Suture Ltd.
- [ ] CDR Medical Industries Ltd.
- [ ] Medtronic
- [ ] Johnson & Johnson
- [ ] None of these

8. Have you heard of any of the following initiatives taken by the government to enhance Indian biomedical sector? *

*Tick all that apply.*

- [ ] The Medical Devices Amendment Rules of 2020 bring all medical devices in India under regulation as drugs
- [ ] A Productions Linked Incentives Scheme for Medical Devices, 2020. Incentive at the rate of 5% of incremental sales over the base year 2019-20
- [ ] Funding for Medical Devices Parks in the country, 2020
- [ ] None of the above
- [ ] All of the above

9. Do you feel that unlike the America (FDA) or the Europe (EU), India lacks a high standard licensing authority for regulation? *

*Mark only one oval.*

- ( ) Yes
- ( ) No

10. What do you think are the major limitations of the biomedical device sector? (Rate- 1=strongly disagree, 2=disagree, 3=neutral, 4=agree, 5=strongly agree) *

*Mark only one oval per row.*

| | 1 | 2 | 3 | 4 | 5 |
|---|---|---|---|---|---|
| Unfavourable duty structure | ◯ | ◯ | ◯ | ◯ | ◯ |
| Limited domestic demand | ◯ | ◯ | ◯ | ◯ | ◯ |
| Lack of Laws and regulations | ◯ | ◯ | ◯ | ◯ | ◯ |
| Deficiency of skilled personnel | ◯ | ◯ | ◯ | ◯ | ◯ |
| Lack of collaboration among govt. and industry | ◯ | ◯ | ◯ | ◯ | ◯ |

11. According to you, what can be done to improve the conditions or lower the limitations faced by the industry? (Rate- 1=strongly disagree, 2=disagree, 3=neutral, 4=agree, 5=strongly agree) *

*Mark only one oval per row.*

| | 1 | 2 | 3 | 4 | 5 |
|---|---|---|---|---|---|
| Reducing tax rate imposed on domestic manufacturers | ◯ | ◯ | ◯ | ◯ | ◯ |
| Increasing skilled workforce | ◯ | ◯ | ◯ | ◯ | ◯ |
| Implementation of strong laws and regulations to improve quality standards | ◯ | ◯ | ◯ | ◯ | ◯ |
| By gaining trust of investors | ◯ | ◯ | ◯ | ◯ | ◯ |
| Collaboration among government and industry | ◯ | ◯ | ◯ | ◯ | ◯ |

### ISO Certification

ISO certification is a seal of approval from a third party body that a company runs to one of the international standards developed and published by the International Organization for Standardization (ISO). The main ISOs are Quality management (ISO 13485), Endurance testing (ISO 10651) and Risk management (ISO 14971). In India effective implementation of ISO standards are lacking. Answer the following questions based on this.

12. How can we improve the Quality Management (ISO 13485) and the Endurance testing (ISO 10651) implementations of biomedical devices in the Indian biomedical sector? (Rate- 1=strongly disagree, 2=disagree, 3=neutral, 4=agree, 5=strongly agree) *

*Mark only one oval per row.*

| | 1 | 2 | 3 | 4 | 5 |
|---|---|---|---|---|---|
| Stringent laws and implementation | ⬭ | ⬭ | ⬭ | ⬭ | ⬭ |
| Formation of strong certification authority | ⬭ | ⬭ | ⬭ | ⬭ | ⬭ |
| Having a knowledgeable and skilled workforce | ⬭ | ⬭ | ⬭ | ⬭ | ⬭ |
| Proper funding from government along with adequate testing facilities | ⬭ | ⬭ | ⬭ | ⬭ | ⬭ |

13. How can we improve the Risk Management (ISO 14971) implementation on biomedical devices in the Indian biomedical sector? (Rate- 1=totally disagree, 2=Slightly disagree, 3=slightly agree, 4=moderately agree, 5=totally agree) *

*Mark only one oval per row.*

| | 1 | 2 | 3 | 4 | 5 |
|---|---|---|---|---|---|
| Safety testing and strict regulations | ◯ | ◯ | ◯ | ◯ | ◯ |
| Development of an effective risk management plan for each devices | ◯ | ◯ | ◯ | ◯ | ◯ |
| Post production analysis of marketed devices | ◯ | ◯ | ◯ | ◯ | ◯ |
| Recruitment of expertees in design of clinical studies for validation | ◯ | ◯ | ◯ | ◯ | ◯ |

14. Is there any gap between Industry and Academia? *

*Mark only one oval.*

◯ Yes

◯ No

15. If yes, how can we improve the association between industry and academia? (Rate-1=strongly disagree, 2=disagree, 3=neutral, 4=agree, 5=strongly agree) *

*Mark only one oval per row.*

| | 1 | 2 | 3 | 4 | 5 |
|---|---|---|---|---|---|
| Industrial Visit/Internships | ⬭ | ⬭ | ⬭ | ⬭ | ⬭ |
| Diffusion of Knowledge through interaction of peers | ⬭ | ⬭ | ⬭ | ⬭ | ⬭ |
| By proposing idea of prototype to product to the industry | ⬭ | ⬭ | ⬭ | ⬭ | ⬭ |
| Collaborative funded start-up | ⬭ | ⬭ | ⬭ | ⬭ | ⬭ |
| Academics should focus more on practical knowledge, in developing soft skills rather than following the entire theoretical course | ⬭ | ⬭ | ⬭ | ⬭ | ⬭ |

### Ventilators

We have endeavored to provide a brief discussion on one of the significant devices, "Ventilator", currently having a mass production due to SARS-CoV-2 outbreak. A ventilator is a machine that "breathes for you" or "helps you breathe". It is also called a breathing machine or respirator. Our key target is to uncover the causes of less production of ventilators and discuss about the complications and problems caused by the mechanical ventilators in a patient.

Ventilator machine

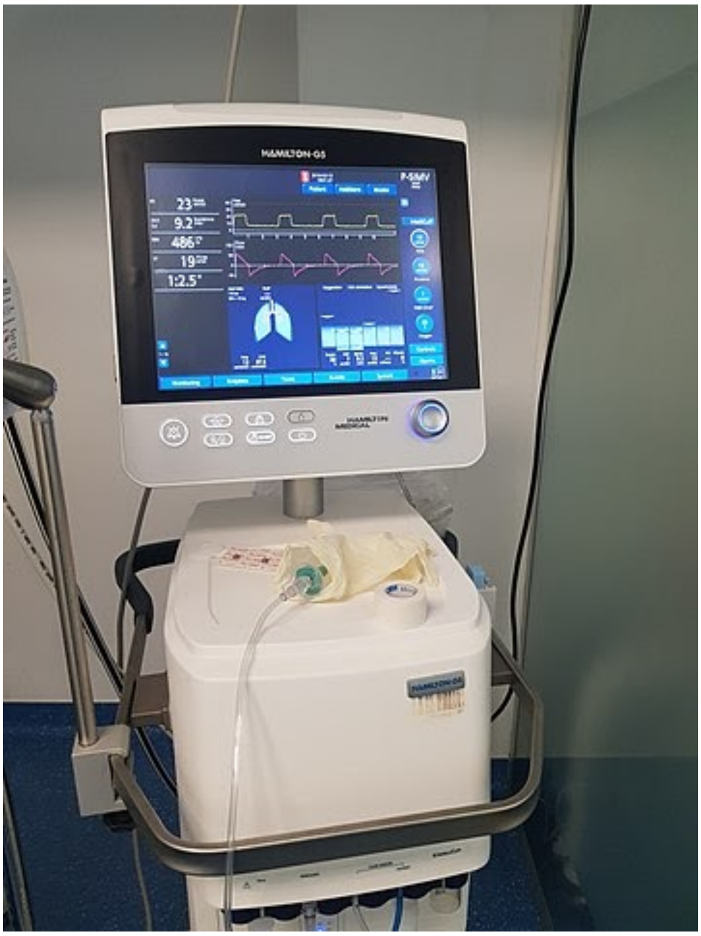


16. Do you know which Indian companies among the following manufacture ventilators? *

*Tick all that apply.*

- [ ] Skanray Technologies
- [ ] AgVa Healthcare
- [ ] Imperial Life Sciences
- [ ] WockHardt Hospital
- [ ] None of these

17. There was a huge rush in the production of ventilators in India during the SARS-CoV-2 outbreak. But, most of the produced ventilators were a failure in the hospitals. Have you heard about this? *

*Mark only one oval.*

- ( ) Yes
- ( ) No

18. Which policy can India adopt to make manufacturing of ventilators easier? (Rate- 1=strongly disagree, 2=disagree, 3=neutral, 4=agree, 5=strongly agree) *

*Mark only one oval per row.*

| | 1 | 2 | 3 | 4 | 5 |
|---|---|---|---|---|---|
| Compulsory Licensing(authorizations permitting a third party to make, use, or sell a patented invention without the patent owner's consent) | | | | | |
| Open source knowledge hub for designing and working of ventilators | | | | | |
| Improve regulatory standards | | | | | |
| Inclusion of a particular course for all the industry personnel | | | | | |

19. What is the reason for less production of ventilators in India? (Rate- 1=strongly disagree, 2=disagree, 3=neutral, 4=agree, 5=strongly agree) *

*Mark only one oval per row.*

| | 1 | 2 | 3 | 4 | 5 |
|---|---|---|---|---|---|
| Companies/Investors are not interested | | | | | |
| Dependency on import of the ventilators | | | | | |
| Lack of skilled personnel | | | | | |
| As the doctors don't trust Indian branded devices | | | | | |

20. A patient named Bhagat was diagnosed with refractory cardiac arrest after being on ventilation. Doctors noted symptoms like hypertension and bradycardia. This might have been caused by tension pneumothorax due to rupture of weakened alveoli or pre-existing blebs. This condition, called barotrauma, occurs due to increased pressure inside the lungs by ventilators. What can be the possible solutions to barotrauma? (Rate- 1=strongly disagree, 2=disagree, 3=neutral, 4=agree, 5=strongly agree) *

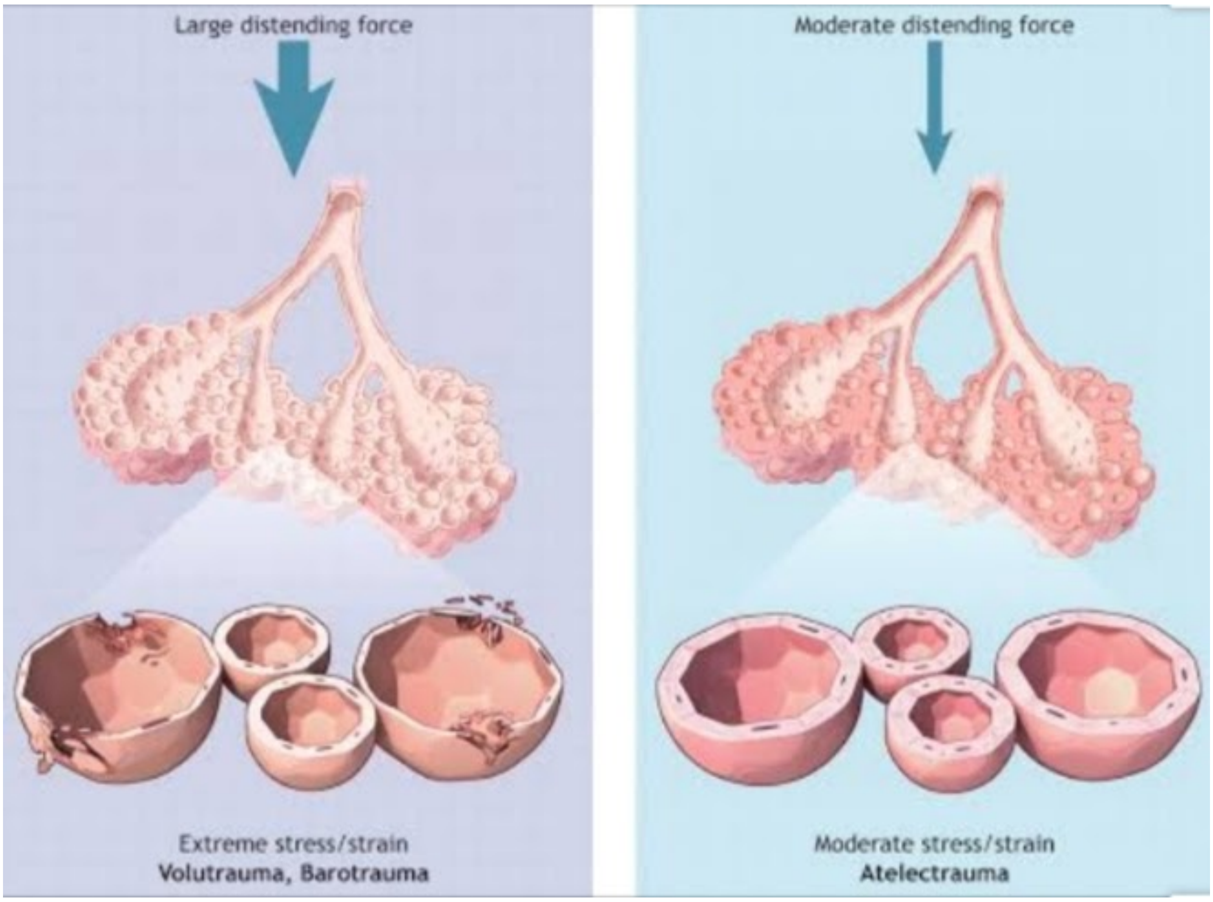


Ruptured alveolis (to the left) due to increased ventilator pressure

*Mark only one oval per row.*

| | 1 | 2 | 3 | 4 | 5 |
|---|---|---|---|---|---|
| Surgical decompression | ⬭ | ⬭ | ⬭ | ⬭ | ⬭ |
| Treatment of underlying pulmonary diseases | ⬭ | ⬭ | ⬭ | ⬭ | ⬭ |
| Manual pressure regulation in the ventilator | ⬭ | ⬭ | ⬭ | ⬭ | ⬭ |
| Artificial intelligence (using | ⬭ | ⬭ | ⬭ | ⬭ | ⬭ |

pressure
sensors)

21. A patient named Rakesh was on ventilation in ICU. He developed symptoms like fever, pus in lung secretions, changes in breathing and low oxygen levels. On clinical testing, it was found that he had a blood leukocyte count of >10,000 cells/ml, purulent tracheal secretions and gas exchange degradation. There was a high suspicion of Ventilator-Associated Pneumonia (VAP). This is caused by the colonization of bacteria like E. coli, S. aureus, etc. in oropharynx and upper airways. What can be the possible treatments for VAP? (Rate- 1=strongly disagree, 2=disagree, 3=neutral, 4=agree, 5=strongly agree) *

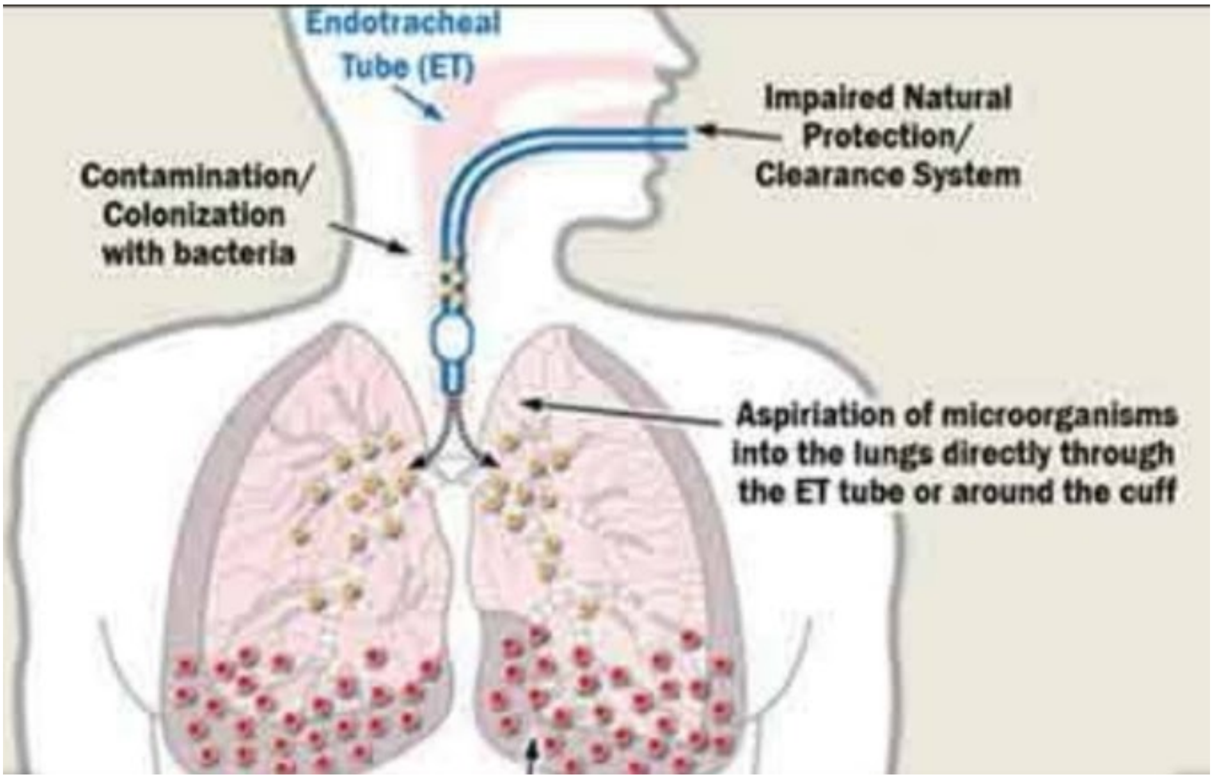


*Mark only one oval per row.*

| | 1 | 2 | 3 | 4 | 5 |
|---|---|---|---|---|---|
| Nebulized and broad sprectrum antibiotics | ⬭ | ⬭ | ⬭ | ⬭ | ⬭ |
| Subglottic secretion drainage (SSD). | ⬭ | ⬭ | ⬭ | ⬭ | ⬭ |
| Antibacterial coating in the ET tube | ⬭ | ⬭ | ⬭ | ⬭ | ⬭ |

| | | | | | |
|---|---|---|---|---|---|
| Introducing non-invasive positive pressure ventilation | ◯ | ◯ | ◯ | ◯ | ◯ |
| Daily weaning trials and sedation holidays | ◯ | ◯ | ◯ | ◯ | ◯ |
| Early tracheostomy | ◯ | ◯ | ◯ | ◯ | ◯ |
| Elevation of head of the bed to reduce aspiration of gastric content | ◯ | ◯ | ◯ | ◯ | ◯ |

22. A person named Urmila is a critically ill intubated patient who has contaminated oropharyngeal and gastric secretions around the tracheal cuff (due to colonization of pathogens). This is called microaspiration. Aspiration of gastric contents is common in patients like Urmila. There were also bacterial growths in the stomach which can enter the lower respiratory tract causing tracheobronchial colonization, that might further progress to VAP. What can be the necessary solutions to reduce microaspiration? (Rate- 1=strongly disagree, 2=disagree, 3=neutral, 4=agree, 5=strongly agree) *

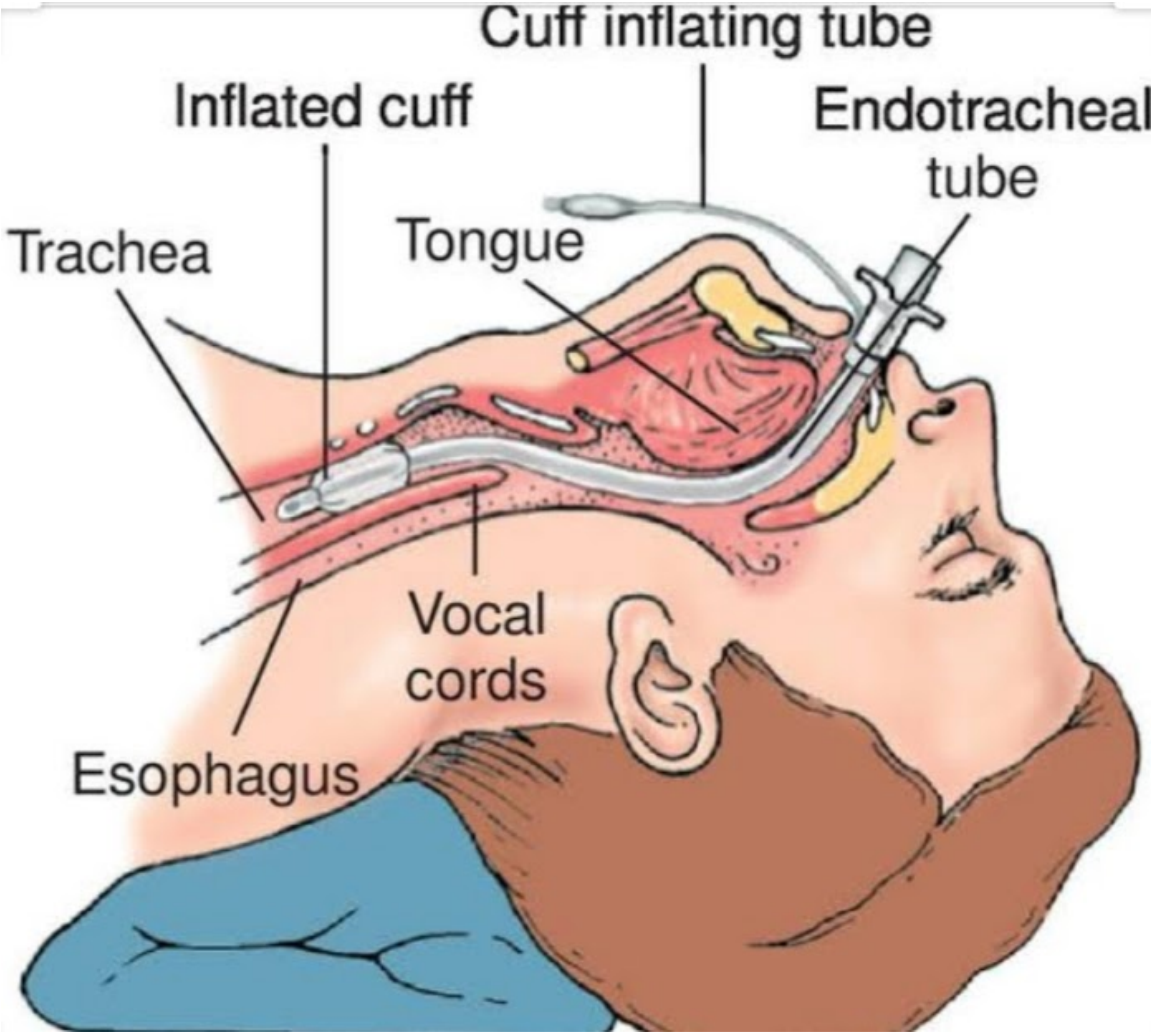


*Mark only one oval per row.*

| | 1 | 2 | 3 | 4 | 5 |
|---|---|---|---|---|---|
| Positive End-Expiratory Pressure | ⬭ | ⬭ | ⬭ | ⬭ | ⬭ |
| Making changes in the cuff material of ET tube | ⬭ | ⬭ | ⬭ | ⬭ | ⬭ |
| Making | ⬭ | ⬭ | ⬭ | ⬭ | ⬭ |

| | | | | | |
|---|---|---|---|---|---|
| Making changes in the cuff shape of ET tube | | | | | |
| Gel lubrication of cuff prior to intubation | ◯ | ◯ | ◯ | ◯ | ◯ |
| Avoiding accumulation of subglottic secretions above the cuff by proper subglottic secretions drainage (SSD) | ◯ | ◯ | ◯ | ◯ | ◯ |
| Monitoring gastric overdistention to avoid bacterial translocation from stomach to respiratory tract | ◯ | ◯ | ◯ | ◯ | ◯ |

23. There were several complications detected in a patient named Latika after she was weaned from mechanical ventilation. A condition, called Acute Respiratory Distress Syndrome (ARDS) has developed resulting in fluid collection in the alveoli of lungs, depriving organs from oxygen. There was an increase dyspnea leading to weaning failure leading to excessive respiratory drive and high ventilatory demands, which has further led to vigorous inspiratory effort resulting in excessive global or regional pulmonary distension due to a nonhomogeneous distribution of stress and strain. What can be the possible ways to reduce ARDS as a side effect of Mechanical Ventilation weaning? (Rate- 1=strongly disagree, 2=disagree, 3=neutral, 4=agree, 5=strongly agree) *

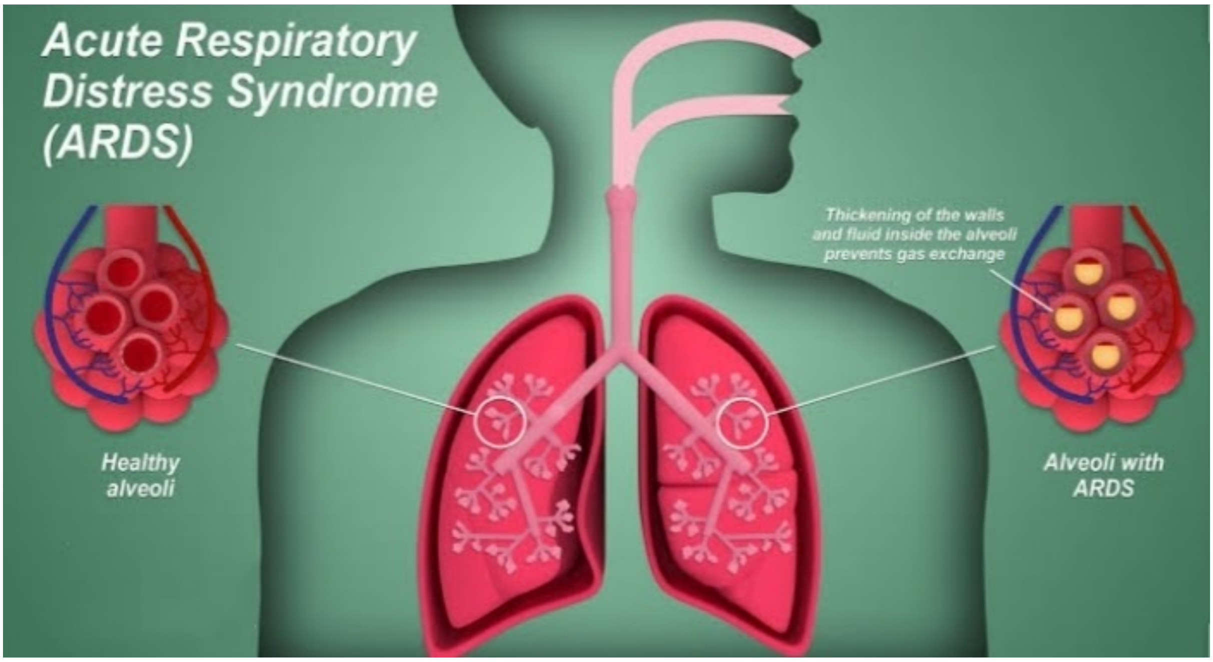


*Mark only one oval per row.*

| | 1 | 2 | 3 | 4 | 5 |
|---|---|---|---|---|---|
| Adopting lung protective ventilation strategies (Low tidal volume and low inspiratory pressure) | ◯ | ◯ | ◯ | ◯ | ◯ |
| Using neuromuscular blocking agents (NMBA) at early stages | ◯ | ◯ | ◯ | ◯ | ◯ |
| Monitoring asynchrony | ◯ | ◯ | ◯ | ◯ | ◯ |
| Prone positioning of | ◯ | ◯ | ◯ | ◯ | ◯ |

Prone positioning of patient for more than 12

This content is neither created nor endorsed by Google.

Google Forms
